# Aging-induced isoDGR-modified fibronectin activates monocytic and endothelial cells to promote atherosclerosis

**DOI:** 10.1101/2021.03.02.21252419

**Authors:** Jung Eun Park, Gnanasekaran JebaMercy, Kalailingam Pazhanchamy, Xue Guo, SoFong Cam Ngan, Ken Cheng Kang Liou, Soe EinSi Lynn, Ser Sue Ng, Wei Meng, Su Chi Lim, Melvin Khee-Shing Leow, A Mark Richards, Daniel J Pennington, Dominique P.V. de Kleijn, Vitaly Sorokin, Hee Hwa Ho, Neil E. McCarthy, Siu Kwan Sze

**Affiliations:** School of Biological Sciences, Nanyang Technological University, 60 Nanyang Drive, Singapore 637551; Diabetes Center, Khoo Teck Puat Hospital, Singapore; Saw Swee Hock School of Public Health, National University of Singapore, Singapore; Cardiovascular and Metabolic Disorders Program, Duke-NUS Medical School, Singapore; Lee Kong Chian School of Medicine, NTU, Singapore; Department of Endocrinology, Tan Tock Seng Hospital, Singapore; Cardiovascular Research Institute, National University of Singapore, Singapore 119228; Christchurch Heart Institute, Department of Medicine, University of Otago, Christchurch 8140, New Zealand; Centre for Immunobiology, The Blizard Institute, Bart’s and The London School of Medicine and Dentistry, Queen Mary University of London, United Kingdom; Department of Vascular Surgery, UMC Utrecht, Utrecht University, Utrecht, the Netherlands; Netherlands Heart Institute, Utrecht, the Netherlands; Department of Cardiac, Thoracic and Vascular Surgery, National University Heart Centre, National University Health System, Singapore 119228; Department of Cardiology, Tan Tock Seng Hospital, 11 Jalan Tan Tock Seng, Singapore 308433

**Keywords:** Atherosclerosis, vascular inflammation, fibronectin, isoDGR motif, integrin, deamidation

## Abstract

**Background and Aims:** Aging is the primary risk factor for cardiovascular disease (CVD), but the mechanisms underlying age-linked atherosclerosis remain unclear. We previously observed that long-lived vascular matrix proteins can acquire ‘gain-of-function’ isoDGR motifs that might play a role in atherosclerotic pathology.

**Methods:** IsoDGR-specific mAb were generated and used for ELISA-based measurement of motif levels in plasma samples from patients with coronary artery diseases (CAD) and non-CAD controls. Functional consequences of isoDGR accumulation in age-damaged fibronectin were determined by bioassay for capacity to activate monocytes, macrophages, and endothelial cells (signalling activity, pro-inflammatory cytokine expression, and recruitment/adhesion potential). Mice deficient in the isoDGR repair enzyme PCMT1 were used to assess motif distribution and macrophage localisation *in vivo*.

**Results:** IsoDGR-modified fibronectin and fibrinogen levels in patient plasma were significantly enhanced in CAD and further associated with smoking status. Functional assays demonstrated that isoDGR-modified fibronectin activated both monocytes and macrophages via integrin receptor ‘outside in’ signalling, triggering an ERK:AP-1 cascade and expression of pro-inflammatory cytokines MCP-1 and TNFα to drive additional recruitment of circulating leukocytes. IsoDGR-modified fibronectin also induced endothelial cell expression of integrin β1 to further enhance cellular adhesion and matrix deposition. Analysis of murine aortic tissues confirmed accumulation of isoDGR-modified proteins co-localised with CD68+ macrophages *in vivo*.

**Conclusions:** Age-damaged fibronectin features isoDGR motifs that increase binding to integrins on the surface of monocytes, macrophages, and endothelial cells. Subsequent activation of ‘outside-in’ signalling elicits a range of potent cytokines and chemokines that drive additional leukocyte recruitment to the developing atherosclerotic matrix.

**Graphical Abstract:** 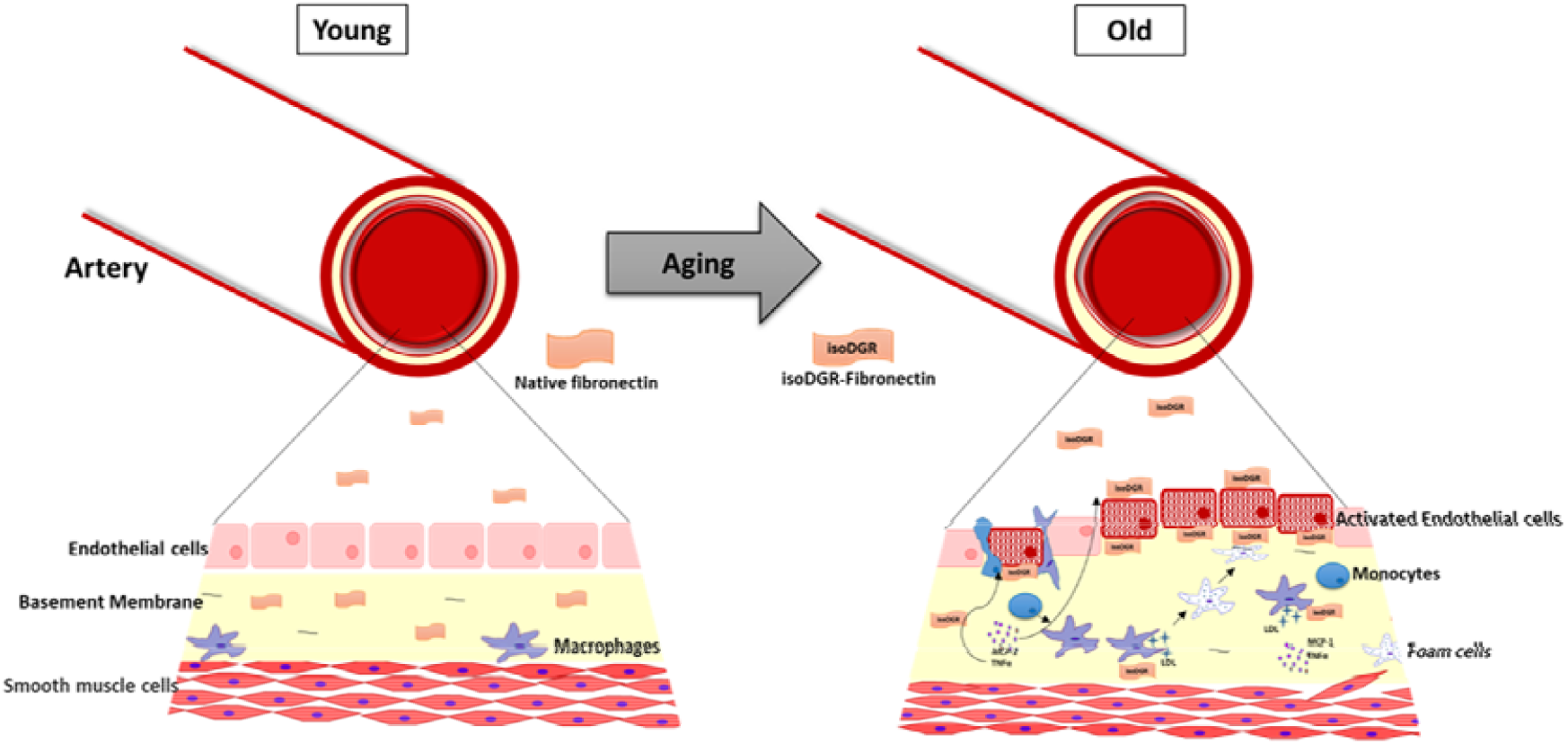

**Highlights:**

- IsoDGR-modified plasma proteins are associated with CAD.
- IsoDGR binding to integrin receptors on monocytes, macrophage, and endothelial cells promotes ‘outside-in’ signalling, monocyte infiltration, and endothelial binding.
- IsoDGR-modified fibronectin may initiate vascular inflammation in atherosclerotic CVD.

## Introduction

Despite recent advances in therapeutic and preventive strategies, cardiovascular disease (CVD) remains a devastating disorder associated with extremely high morbidity and mortality across the world^1^. Previous research has now clearly demonstrated that aging is the primary risk factor for this complex, multifactorial disorder. Early events in CVD pathogenesis include endothelial dysfunction, vascular inflammation, and atherosclerotic plaque formation, ultimately leading to luminal narrowing and thrombotic occlusion of blood vessels^2,3^. While these key processes have now been investigated in detail, it remains unclear how tissue aging initiates the events required for later progression to overt pathology.

We recently reported that extensive deamidation of the amino acid sequence NGR (Asn-Gly-Arg) in extracellular matrix (ECM) proteins results in ‘gain-of-function’ conformational switching to isoDGR (isoAsp-Gly-Arg) motifs^4-7^ that can mimic integrin-binding RGD ligands^8-10^. Atypical isoDGR modifications have previously been detected in fibronectin, laminin, tenascin C, and several other ECM proteins derived from human carotid plaque tissues^4-7^, suggesting that these molecules may be capable of enhancing leukocyte binding to the atherosclerotic matrix. Long-lived ECM components are well recognized to accumulate degenerative protein modifications (DPMs) including spontaneous deamidation^11-17^, under the influence of microenvironmental stresses, flanking amino acid sequences, and genetic factors^13,17^. DPMs are typically associated with loss of protein function and have long been regarded as critical events in human aging and degenerative disorders, but recent data have suggested that DPMs may also induce ‘gain of function’ structural changes that could play equally important roles in human pathology^4-10^. While age is the largest risk factor for many human chronic diseases including CVD, the mechanisms by which advancing age promotes atherosclerosis and CVD have remained obscure and the therapeutic opportunities these may provide are currently unknown. Having previously observed that long-lived ECM proteins undergo age-dependent spontaneous deamidation to generate isoDGR motifs, we postulate that age-related structural damage to key vascular proteins may represent the ‘missing link’ in common pathologies affecting the elderly. In particular, we hypothesize that atypical isoDGR:integrin interactions may serve as a key early trigger for endothelial dysfunction, macrophage activation, monocyte recruitment, and vascular inflammation leading to atherosclerotic plaque development.

Fibronectin is among the first matrix proteins deposited at sites of atherosclerosis^18,19^. Human fibronectin features a RGD integrin binding motif, as well as 6 NGR motifs that can be modified by spontaneous asparagine deamidation to isoDGR. Indeed, we have previously detected 4 characteristic isoDGR motifs in fibronectin derived from human carotid atherosclerotic plaques (at residues 263-265, 367-369, 501-503 and 1432-1434)^7^. In the current study, we generated hybridoma cells that produce isoDGR-specific mAb and aimed to determine the effects of age-damaged fibronectin on monocyte and macrophage function in the early stages of CVD pathogenesis. We observed that interaction of isoDGR motifs with integrins on the surface of monocytes and macrophages triggered an ERK:AP-1 signalling cascade that induced expression of a range of potent chemotactic mediators that promoted further leukocyte infiltration of the assembling atherosclerotic matrix. Biochemical and functional assays also demonstrated that isoDGR-modified FN promoted endothelial activation via expression of integrin β1, which enhanced adhesion of blood leukocytes and facilitated protein deposition resembling intimal thickening in atherosclerosis. Crucially, administration of isoDGR-specific mAb was sufficiently to potently inhibit all of the pathological features assessed *in vitro*, suggesting that antibody blockade of this structure may represent a viable therapeutic option. Consistent with *in-vitro* results, we observed *in-vivo* accumulation of isoDGR-modified proteins co-localised with CD68+ macrophages in aortic tissues of mouse model. In addition, we detected significant enrichment of isoDGR motifs in plasma fibronectin from CVD patients analysed *ex vivo*, indicating isoDGR may be a predictive biomarker of CVD.

## Materials and methods

### Patients and clinical samples

Study participants were recruited from patients undergoing coronary artery bypass grafting (CABG) or cardiac computed tomographic angiography (CCTA) at the Department of Cardiac, Thoracic and Vascular Surgery at the National University Heart Centre, National University Health System (NUHS) or the Department of Cardiology at Tan Tock Seng Hospital (TTSH). The study was approved by the institutional review boards of NTU (IRB-2017-01-013), NUHS (IRB-NUH-2009-0073) and TTSH (TTSH-2013-00930). Experimental procedures complied with the tenets of the Declaration of Helsinki. Informed written consent was obtained from each subject prior to the inclusion in the study. For detailed demographic information and clinical history see; Table 1, Supplementary Data 1 (S.Tables 1 and 2), Fig. 1. Patients who were diagnosed with atherosclerotic coronary artery disease (CAD) by angiography and underwent CABG were included (n=25) and age-matched control subjects were selected from patients who were assessed by angiography/CCTA on clinical suspicion but found to be free of CAD (n=25). All blood samples were collected prior to angiography, CCTA or CABG.

**Table 1.** Demographic summary of clinical cohorts.

|  | **Ischemic Heart Disease**<br>**(IHD)**<br>**(N=25)** | **Control**<br>**(Ctrl)**<br>**(N=25)** |
| --- | --- | --- |
| Age [Mean (± SD)] | 63 (± 7.6) | 54 (± 8.7) |
| Gender [N (%)] |  |  |
| Male | 18(72) | 15 (60) |
| Female | 7(28) | 10(40) |
| Chinese [N (%)] | 25 (100) | 25(100) |
| Treatment [N (%)] |  |  |
| CABG | 25(100) | NA |
| Conservative | NA | 25(100) |
| Hypertension [N (%)] |  |  |
| Hypertension | 19 (76) | 5 (20) |
| No Hypertension | 6(24) | 20(80) |
| Hyperlipidemia [N (%)] |  |  |
| Hyperlipidemia | 25(100) | 8 (32) |
| No Hyperlipidemia | NA | 17(68) |
| Diabetes Mellitus [N (%)] | 15 (60) | 14 (70) |
| Lipid medication (statin therapy) [N (%)] | 25(100) | 10(40) |
| Smoking [N (%)] |  |  |
| Smoking | 9(36) | 1(4) |
| No Smoking | 14(56) | 24(96) |
| Missing information | 2(8) | NA |

**Figure 1:**
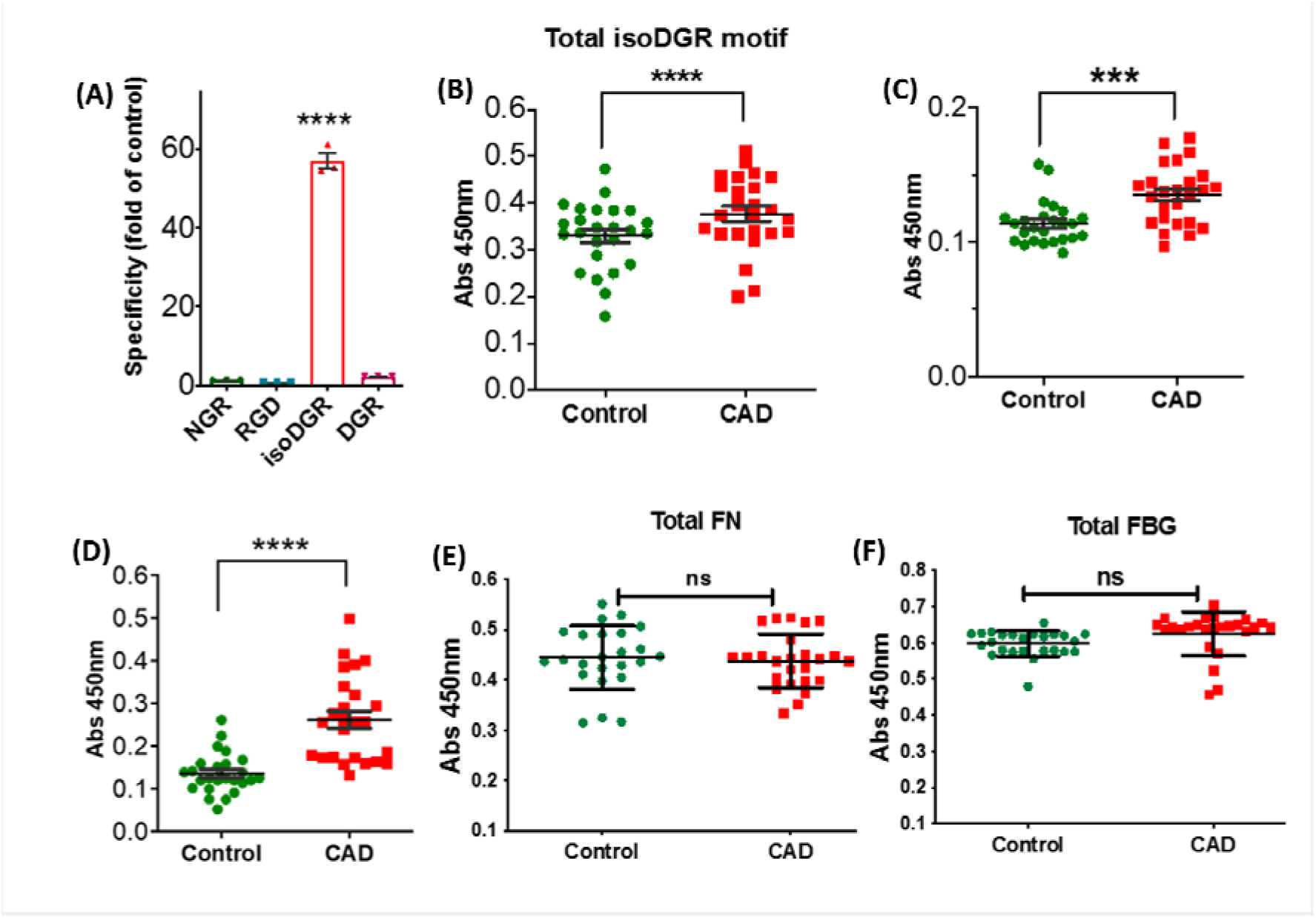
IsoDGR detection in blood plasma from patients with coronary atherosclerosis. **(A)** ELISA results using synthetic peptides indicated that mAb specifically binds peptides that contain an isoDGR motif (but not those containing unmodified NGR, RGD, or DGR sequences). **(B)** Measurement of total isoDGR levels in plasma samples from control subjects (n=25) or CAD patients (n=25) by indirect ELISA with isoDGR-specific mAb. **(C) and (D)** Sandwich ELISA analysis of isoDGR motif level in plasma FN (c) and FBG (d) in the same set of plasma samples. **(E) and (F)** Sandwich ELISA analysis of native/unmodified plasma FN (e) and FBG (f). ELISA assay was performed with three technical replicates (n=3). Statistical differences between groups were determined by unpaired T-test; *\*\*\*p=0*.*003, ****p<0*.*0001*, ns=no significant difference.

### Cell lines and culture methods

Cell culture media and supplements were obtained from Life Technologies Holdings Pte Ltd. (Grand Island, NY, USA) unless otherwise specified. Human monocytic cell lines THP-1 and U937, and murine macrophage cell line RAW264.7 were cultured in RPMI-1640 medium supplemented with 10% fetal bovine serum (FBS) and 1% penicillin/streptomycin (P/S) at 37°C in 5% CO_2_. Human umbilical vein endothelial cells (HUVECs) were cultured in DMEM containing 10% FBS and 1% P/S. All cell lines were tested for mycoplasma contamination by PCR according to published methods^20^. Macrophages were differentiated from U937 and THP-1 cells by incubating the cells with 100ng/ml Phorbol 12-myristate 13-acetate (PMA) (Sigma-Aldrich, St. Louis, MO, USA) for 1h in RPMI-1640 medium containing 1% FBS. The RAW264.7 cells, monocytic U937/THP-1 cells and derivative macrophages were seeded onto fibronectin-coated 24 well plates. To assess the role of isoDGR:integrin receptor interactions in CVD, cell pellets were collected for western blot and RT-PCR analysis to determine ‘outside-in’ signalling activity and cytokine expression. Cell-conditioned medium was also assayed for pro-inflammatory cytokine levels. In some experiments, isoDGR:integrin interactions was blocked by addition of isoDGR-specific mAb (10µg/ml).

### Cytokine multiplex bead assay

The LEGENDplex^Tm^ mouse inflammation panel (Biolegend, San Diego, CA) was used to measure 13 different cytokines (IL-23, IL-1α, IL-1β, IL-6, IL-10, IL-12p70, IL-17A, IL-23, IL-27, MCP-1, IFN-β, IFN-γ, TNF-α, and GM-CSF) in cell culture supernatants after 48h incubation (assessed by LSRII flow cytometer according to the manufacturer’s protocol).

### Secreted alkaline phosphatase (SEAP) reporter assay

U937 cells were reverse-transfected with 50ng AP1-SEAP or NFκB-SEAP vector, then seeded onto native or deamidated fibronectin-coated 96-well plates and incubated for 24h. Culture supernatants were collected, heated at 65°C for 30min, then assayed for alkaline phosphatase activity as follows; 30µl of supernatant was incubated with 120µl assay buffer for 5min, after which 1:20 diluted CSPD substrate was added and the samples read on a TECAN microplate reader (Maennedorf, Switzerland).

### Transwell migration assay

Native or deamidated fibronectin were coated onto 96-well plates (2.5µg/100µl volume per well) via overnight incubation at 4°C. Control wells were prepared with 1% bovine serum albumin (BSA) in PBS and transmigration assays were carried out using n=5 technical replicates. Briefly, U937 cells were primed with PMA (100ng/ml) for 1h at 37°C then re-suspended in 1% FBS-RPMI medium and seeded onto fibronectin-coated plates at a density of 1 × 10^5^ cells/well for 6h. Thereafter, 4 × 10^4^ CSFE-labelled, non-activated U937 cells were added to matrigel-coated inserts and incubated for a further 24h at 37°C. Cell transmigration in response to molecules secreted by fibronectin-bound monocytes was monitored using fluorescence microscopy and quantified by crystal violet staining. IsoDGR-specific mAb was added to the deamidated fibronectin condition to determine motif specificity of these effects.

### Fibronectin fibrillogenesis and DOC solubility assay

Native and deamidated fibronectin were labelled with Alexa488 dye and used for exogenous plasma fibronectin assembly assays. Briefly, HUVECs were seeded onto coverslips and allowed to adhere and grow to 90-100% confluency. Subsequently, cells were treated with either native or deamidated FN (25µg/ml) in 2% FBS-DMEM media containing PMA (100ng/ml) and incubated for the indicated times at 37°C with or without steady flow (100rpm). Fibronectin deposition and/or matrix assembly were monitored using fluorescence microscopy and subsequently processed for sodium deoxycholate (DOC) solubility assays as previously described^21^. Briefly, HUVECs were put on ice and lysed with cold DOC lysis buffer (2% DOC; 20mM Tris–HCl, pH 8.8; 2mM PMSF; 2mM EDTA; 2mM iodoacetic acid; 2mM N-ethylmaleimide). Cells were lysed using a 26G syringe needle and immediately centrifuged at 15,000rpm for 15min at 4°C. The supernatant was collected as DOC-soluble fraction while the pellet was washed with DOC buffer and then solubilized in 25μl SDS buffer (1% SDS; 20mM Tris–HCl, pH 8.8; 2mM PMSF; 2mM EDTA; 2mM iodoacetic acid; 2mM N-ethylmaleimide). DOC-soluble and DOC-insoluble fractions were subsequently resolved by SDS– PAGE using 7.5% polyacrylamide gels, then transferred onto nitrocellulose membranes and immunoblotted with anti-fibronectin mAb.

### Murine model

Mice deficient in the deamidation repair enzyme *PCMT1* (C57BL/6 background) were obtained from The Jackson Laboratory (*PCMT1*^*−/−*^ mice die prematurely, so *PCMT1*^*+/−*^ animals were used in all experiments)^22^. Mouse genotype was confirmed by PCR using the following primers: oIMR1080:CGG CTG CAT ACG CTT GAT C, oIMR1081:CGA CAA GAC CGG CTT CCA T, oIMR1544:CAC GTG GGC TCC AGC ATT, oIMR3580:TCA CCA GTC ATT TCT GCC TTT G. Mice were maintained on normal chow diets and housed with regular light/dark cycles.

### Immunostaining of aorta sections

Aorta from 8-month-old WT and *PCMT1*^*+/−*^ mice were collected and fixed with 4% paraformaldehyde at 4°C for 24h. The aorta tissues were washed with PBS and supplemented with 15% sucrose then 30% sucrose over 1 day. Aortas were embedded in OCT compound with dry ice. A Leica CM3060S Cryostat was used to prepare 10μm cross sections of aorta that were mounted on Fisherbrand Superfrost plus Microscope slides. The slides were kept in warm PBS for 20min to remove OCT before being permeabilized and treated for 1h with blocking buffer (2.5% normal Goat serum, 1% BSA in 0.5%PBST) prior to addition of primary antibodies (isoDGR [1:200] and CD68 [1:200]) overnight at 4 °C. The slides were then incubated with secondary antibodies conjugated to AlexaFluor 488 and 594 for 1h at RT. DAPI staining for 30min was used to visualize cell nuclei. After staining, the slides were washed with 1X PBS and mounted with aqueous mounting media. Images were acquired using a Zeiss LSM710 confocal microscope.

### Statistical analysis

Statistical analyses were performed using GraphPad Prism 5.0 (GraphPad Software, Inc., San Diego, CA). Differences between groups were assessed either by one-way ANOVA or Student’s t-test (p<0.05 was considered significant).

## Results

### IsoDGR motifs are increased in blood plasma from patients with coronary artery disease (CAD)

Deamidation of NGR (Asn-Gly-Arg) amino acid sequences in extracellular matrix (ECM) proteins creates ‘gain-of-function’ isoDGR motifs (isoAsp-Gly-Arg) implicated in atherosclerotic plaque formation, so we developed a specific monoclonal antibody against isoDGR to enable analysis of this structure in human patient samples (Fig. 1). In previous analyses of human carotid plaque tissues, we identified high levels of deamidation in core proteins fibronectin (FN) and fibrinogen (FBG) which are known to play critical roles in CVD ^23,24^. To determine isoDGR levels in blood plasma from CVD patients, we collected samples from n=25 CAD patients immediately prior to CABG as well as n=25 control subjects (confirmed to be free of CAD by angiography or CCTA; Table 1, Supplementary Data 1 (S.Tables 1 and 2). An isoDGR-specific mAb was then used to measure total isoDGR-modified plasma proteins by ELISA (SFig. 1).

These analyses showed that levels of total isoDGR-modified plasma proteins were significantly increased in CAD relative to control patients (p<0.0001) (Fig. 1b). Similarly, when we specifically measured levels of native versus modified FN and FBG in blood plasma, we observed that isoDGR-modified variants of these proteins were significantly higher in CAD (Fig. 1c and d), even though total concentrations of FN and FBG were comparable between groups (Fig. 1e and f). IsoDGR formation is predominantly a consequence of aging, but it is unclear whether accumulation of this motif can also be influenced by other CVD risk factors. We therefore assessed total isoDGR levels in CAD and CTRL subjects were correlated with clinical hypertension (SFig. 2a), smoking status (SFig. 2b), or hyperlipidemia/statin therapy (SFig. 2c). These analyses revealed a significant association of smoking status with higher levels of plasma isoDGR in CAD patients that warrants further investigation in future.

### isoDGR formation in fibronectin induces activation of integrin ‘outside-in’ signaling pathways

Human fibronectin features a RGD integrin-binding motif and 6 potential isoDGR motifs that may significantly impact human health in advancing age. In previous work, we reported that fibronectin in carotid plaques accumulates high levels of isoDGR in 4 of 6 possible motifs, and these motifs are capable of enhancing monocyte adhesion to fibronectin-coated plates via interaction with cell-surface integrins^7^. In the current study we sought to determine the biological consequences of integrin-mediated immune cell adhesion to isoDGR-modified fibronectin. To do this, we incubated the native protein in mild alkaline condition (pH 8.0) to promote spontaneous deamidation of the NGR sequence. The deamidation of Asn amino acid residue via a succinimide intermediate generates a mixture of D- or L-Asp, and D- and L-isoAsp products. The reaction mixture was therefore separated and characterized by RPLC-MS/MS using a C18 column and ERLIC-MS/MS using WAX column^25-28^. Native fibronectin displayed little evidence of deamidation (Supplementary Data 2A), whereas 3 isoDGR motifs that were previous identified in human carotid atherosclerotic plaque were detected in the treated FN protein (Supplementary Data 2 to 6). The reaction should generate the D-/L-stereoisomers that were not distinguished by our non-chiral RPLC and ERLIC separation, likely because typical D-/L-stereoisomers require chiral chromatography for separation. However, peptides featuring NGR, D-/L-DGR and D-/L-isoDGR motifs derived from the same precursor sequence could be well-separated with ERLIC-MS/MS peptides following the expected elution order (-NGR-< - DGR- < -isoDGR-), thereby facilitating estimation of isoDGR motif yields (DGR : isoDGR, ∼1 : 3). Based on label-free quantitation using XIC of the LC-MS/MS data, our fibronectin deamidation method induced isoDGR motifs at residues 263-265 (∼10%), 501-503 (3%), and 1432-1434 (8%). The isoDGR motif at residues 263-265 is generated as the major product located on fibronectin subunit I, facing outwards from the main body of the molecule (see 3D structure in Fig. 2A and B), suggesting that this *de novo* structure is likely to be readily accessible for integrin binding.

**Figure 2:**
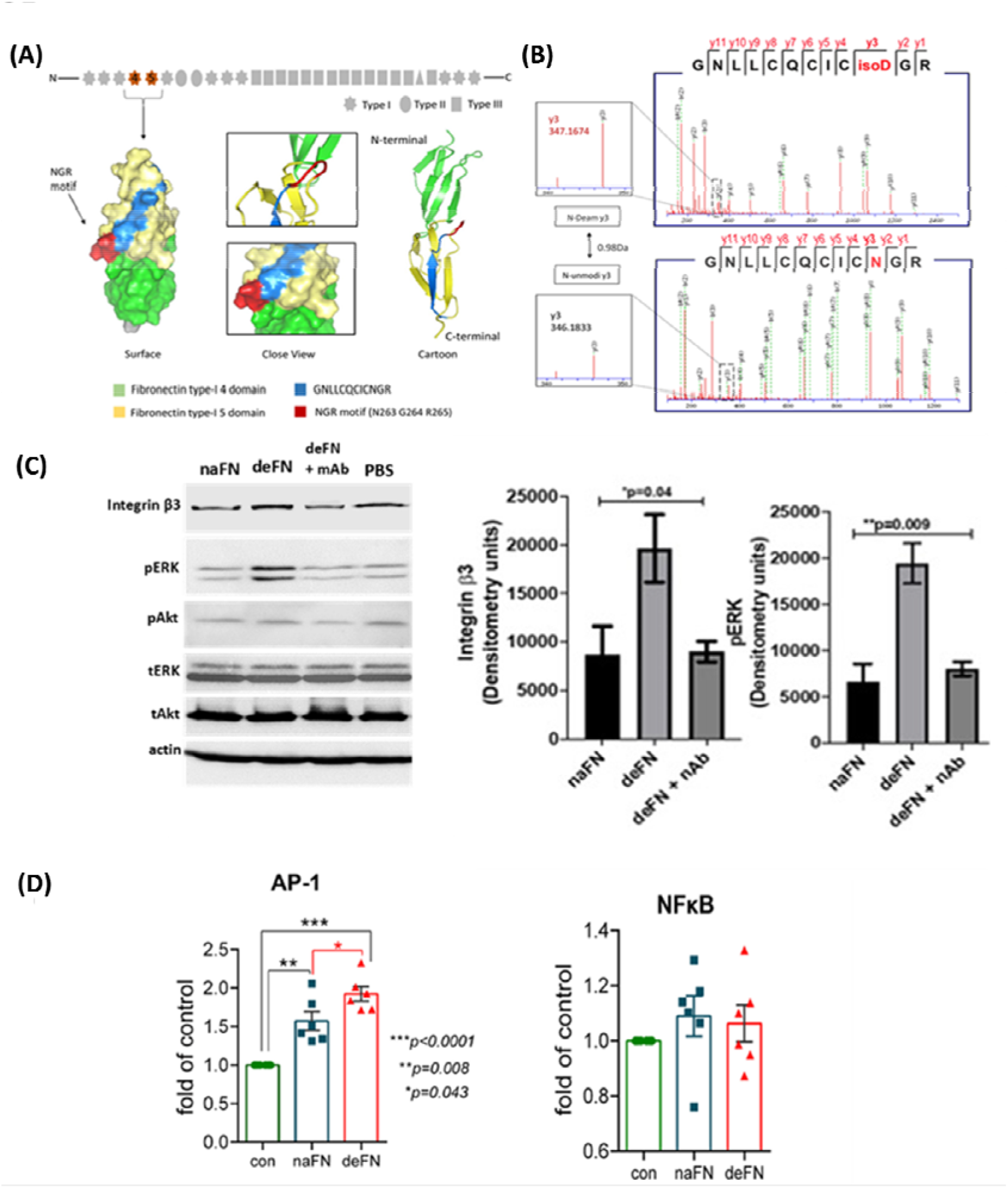
IsoDGR-modified fibronectin actives macrophages via pERK. **(A)** The NGR motif (red) is located within the type I domain and faces outwards from the main body of the molecule. **(B)** HCD mass spectra of peptide _254_GNLLCQCIC(isoD)/(N)GR_265_ confirming deamidation of the Asn residue (+0.98Da) accompanied by characteristic y ion series (y2–y11, solid red lines), and neutral losses (dashed green lines) of NHCO (43 Da) and NH3 (17 Da). **(C)** RAW cells (macrophages) were cultured in 24-well plates coated with either native or deamidated fibronectin for 24h. Exposure to isoDGR-modified protein activated RAW cells via ERK phosphorylation (lower panel) whereas an isoDGR-specific mAb was able to block these effects. Statistical differences between groups were determined by one-way ANOVA. Similar results were obtained using PMA-primed U937 cells (Fig. 3d). **(D)** U937 cells were reverse-transfected with AP-1 or NFκB-SEAP vector and cultured on native or deamidated fibronectin for 24h. U937 cell adhesion to deamidated fibronectin was associated with marked activation of the AP-1 signal transduction pathway but not NFκB. Each column represents the mean□±□SE of three biological replicates (two technical repeats per biological replicate). Statistical differences between groups were determined by unpaired T-test.

To assess the biological consequence of fibronectin deamidation, murine RAW macrophages were cultured in 24-well plates containing either native or deamidated fibronectin for 24h incubation then subjected to total protein extraction and western blot analysis. As shown in Fig. 2c, macrophages displayed high basal levels of integrin β3 expression that were increased upon adhesion to deaminated fibronectin, whereas AKT phosphorylation was unaltered by fibronectin binding state. Instead, interaction of macrophage integrins with deamidated fibronectin appeared to activate the alternative MAPK-ERK signaling pathway as indicated by phosphorylation of ERK1/2. The specific requirement for isoDGR-modified fibronectin to trigger ‘outside-in’ signaling was confirmed by addition of an isoDGR neutralizing antibody that blocked integrin β3 upregulation and ERK1/2 activity (Fig. 2c). Similar observations were confirmed in human macrophages generated from PMA-primed U937 monocytes (Fig. 3d). To further investigate the effects of NGR-to-isoDGR molecular switching on downstream pathways of integrin ‘outside-in’ signalling, we next used reporter constructs containing the secreted alkaline phosphatase (SEAP) gene under the control of either AP-1 or NF-κB promoters to quantify pathway activation. For this assay, we used human U937 cells (Fig. 2d). Monocytes binding to deamidated fibronectin displayed a significant increase in AP-1-induced SEAP activity compared to native protein-exposed cells, indicating that isoDGR motifs can promote signal transduction through this axis and potentially upregulate downstream gene targets of AP-1. Intriguingly, isoDGR-triggered integrin signalling appeared independent of NF-κB which was not activated in our assays despite this pathway playing a central role in regulating inflammation^29^.

**Figure 3:**
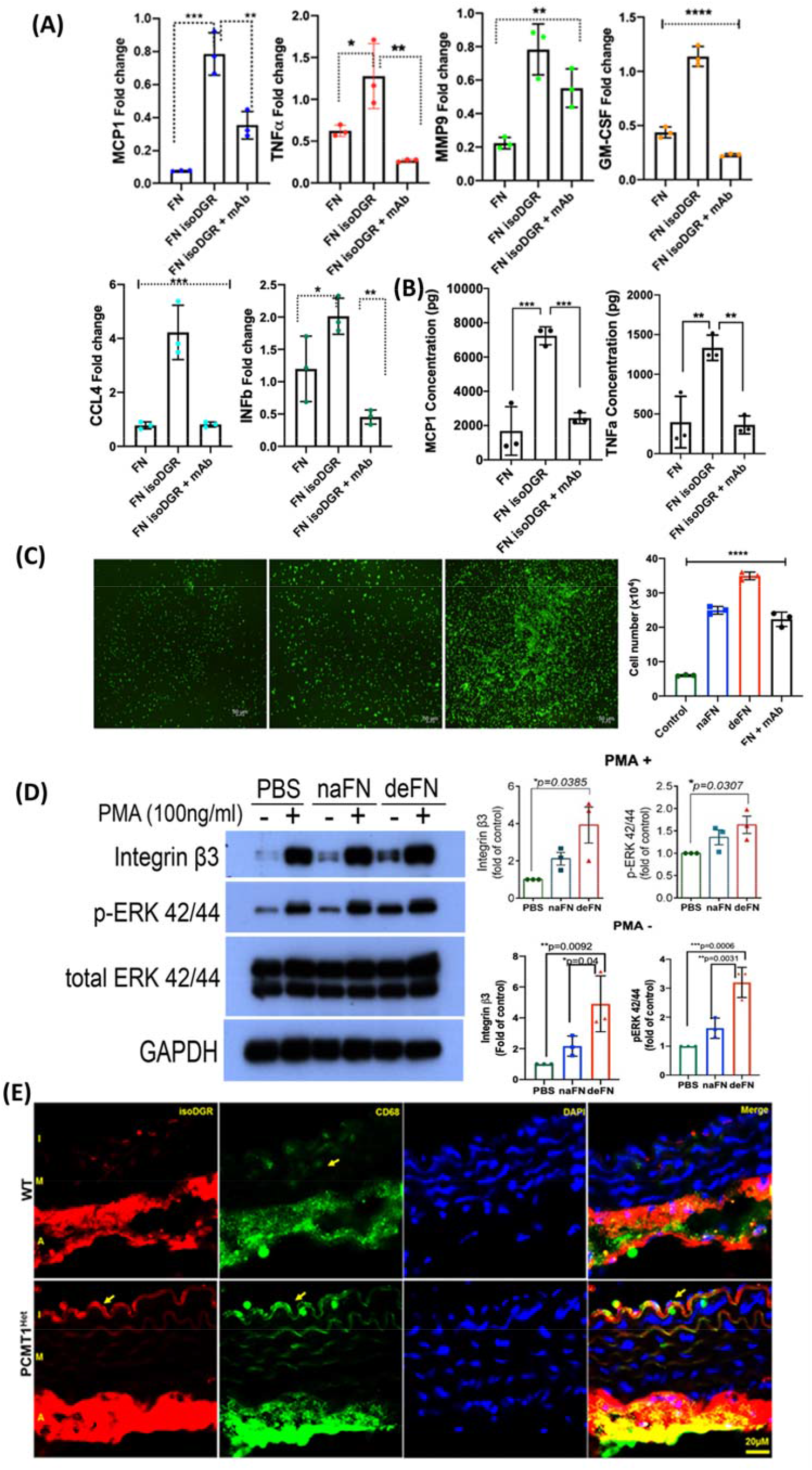
IsoDGR-modified fibronectin activates macrophages expression of chemotactic MCP-1 and TNFα. **(A)** RAW macrophage cells were cultured for 24h together with native or deamidated fibronectin before mRNA expression levels of key cytokines were assessed by qRT-PCR. This analysis revealed that macrophages exposed to isoDGR-modified fibronectin displayed significant upregulation of several chemotactic molecules, pro-inflammatory cytokines, and metalloproteinase enzyme MMP9. Experiments included n=3 biological replicates (3 technical repeats per biological replicate, data shown are mean ± S.E. and were assessed by unpaired T-test). **(B)** Legendplex^TM^ bead assay of proinflammatory cytokine secretion by RAW macrophages. Exposure to isoDGR-modified fibronectin stimulated macrophage expression of both MCP-1 (>4 fold) and TNFα (>3-fold). **(C)** In Transwell experiments, adhesion of U937 cells to deamidated fibronectin (bottom chamber) induced greater transmigration of unstimulated monocytes (matrigel-coated top chamber) than did exposure to the native protein control (assessed by microscopic or colorimetric analysis after 24h incubation). All pathological processes described in panels a-c were inhibited by addition of 10μg/ml isoDGR-specific mAb. Experiments included n=5 biological replicates. Statistical differences between groups were determined by unpaired T-test. **(D)** U937 (PMA- and PMA+) monocytes were cultured in 24-well plates coated with either native or deamidated fibronectin for 24h, during which time ERK phosphorylation was induced only in cells exposed to the isoDGR-modified protein (lower panel). Statistical differences between groups (n=3) were determined by unpaired T-test. Expression of integrin β3 and pERK were strongly increased when U937 cells (PMA-) were exposed to isoDGR-modified fibronectin, indicating active monocyte differentiation into macrophages. **(E)** Immunofluorescent imaging of aorta sections (I: intima; M: media; and A: adventitia layers) showing isoDGR-motif distribution and co-localisation of CD68+ macrophages in 8-month-old WT and *PCMT1*^*+/−*^ mice.

### NGR switching to isoDGR increases expression of chemotactic mediators

Since AP-1 transcription factors regulate cytokine gene expression in response to a variety of external signals,^30,31^ we next examined mRNA expression levels of pro-inflammatory mediators after 24h exposure of RAW (Fig. 3a) or PMA-primed U937 cells (SFig. 3) to either native or deamidated fibronectin. Fig. 3a displays expression levels of various cytokine/chemokine genes in murine macrophages, indicating that exposure to deamidated fibronectin induced substantial upregulation of several chemotactic molecules including monocyte chemoattractant protein (MCP)-1, GM-CSF, CCL4, and INF-β, as well as the metalloprotease enzyme MMP9 and associated pro-inflammatory cytokine TNFα. Since secretion of chemokine(s) by tissue-resident macrophages promotes monocyte recruitment to the vessel walls, we next measured 13 pro-inflammatory mediators in the cell-conditioned medium using a multiplex bead array. These analyses showed that levels of MCP-1 and TNFα in the conditioned medium were increased by >4-fold and >3-fold respectively upon macrophage exposure to deamidated fibronectin (Fig. 3b). Accordingly, macrophage secretion of MCP-1 and TNFα were blocked by addition of neutralizing isoDGR-specific mAb, indicating that these events depended on isoDGR motifs.

Since macrophage adhesion to isoDGR-modified fibronectin induced marked upregulation of pro-inflammatory cytokine genes and high secretion of MCP-1 and TNFα, we next hypothesized that altered patterns of cytokine expression may be a key event promoting further leukocyte accumulation and establishing a ‘positive feedback loop’ that drives pathology in CVD. To test this, we established a Transwell migration assay using PMA-primed U937 cells that were seeded into the bottom chambers of culture plates that were coated with either native or deamidated fibronectin (or uncoated control wells). After 6h incubation, unstimulated monocytes pre-labelled with CFSE were seeded into the Matrigel-coated upper chambers and their migration patterns were monitored by fluorescence microscopy. As shown in Fig. 3c, U937 cells exposed to deamidated fibronectin displayed >2-fold more transmigration than did cells exposed to either native fibronectin or uncoated control wells. The addition of isoDGR neutralizing antibody significantly reduced/blocked transmigration in these assays (SFig. 4). Together, these data confirmed that macrophage adhesion to isoDGR-modified fibronectin triggers ‘outside in’ signalling to promote the release of soluble factors that promote further leukocyte recruitment to the age-damaged matrix and likely represents a key early event in atherosclerotic plaque formation/progression. Indeed, subsequent differentiation of infiltrated monocytes into intima-resident macrophages is another key step in atherosclerosis, and U937 cells exposed to deamidated fibronectin displayed significantly increased expression of macrophage-associated integrin β3 and pERK42/44 activity (Fig 3d, PMA-data).

The above *in-vitro* functional assays strongly suggested that isoDGR-modified fibronectin is a critical mediator of vascular inflammation via interaction with resident macrophages in early stage atherosclerotic CVD. We therefore sought to determine whether this process can also be enhanced by isoDGR-modified proteins *in-vivo* using a mouse model deficient in the deamidation repair enzyme PIMT (L-isoaspartyl methyltransferase, or PIMT/PCMT1)^22^. Since *PCMT*^*−/−*^ mouse died prematurely, we used 8-month old WT and *PCMT*^*Het*^ mice to study the accumulation of isoDGR motifs and CD68+ macrophages in aorta tissues over time. When assessed by immunofluorescent imaging, we observed marked accumulation of isoDGR motifs in both intima and adventitial layers of both WT and *PCMT*^*+/−*^ mice, but more extensive accumulation of isoDGR-modified proteins with co-localised CD68+ macrophages in aorta from transgenic animals (Fig 3e). Combined with our *in vitro* results, these *in vivo* data confirm an important role for isoDGR motifs in vascular inflammation and immune cell recruitment in the early phases of atherosclerosis.

### Deamidated fibronectin activates endothelial cell adhesion via expression of integrin β1

Activated endothelium expresses a series of β1-containing integrin pairs that can mediate cell adhesion (α1β1, α2β1, α3β1, α5β1 and α6β1)^32^, so we next sought to determine whether isoDGR-modified fibronectin could alter patterns of β1. To test this, we added either native or deamidated fibronectin (25µg/ml) to PMA-primed human umbilical vein endothelial cells (HUVECs) for 24h and performed immunofluorescent staining for active integrin β1 (using antibody clone HUTS-4 which recognizes epitopes in the 355–425 region of active β1 subunits^33^). Levels of activated integrin β1 where markedly increased on HUVECs exposed to deamidated fibronectin relative to those treated with native protein alone. Active integrin β1 was also more clearly distributed to the plasma membrane in the presence of isoDGR-modified fibronectin (Fig. 4a, 4b). To further elucidate possible activation signals triggered by modified fibronectin, we next exposed PMA-primed HUVECs to a range of synthetic peptides containing either NGR, RGD, or isoDGR motifs for 24h before repeating the immunofluorescent analyses. Unlike peptides containing NGR or RGD sequences, ligands that featured an isoDGR motif potently induced integrin β1 activation and distribution to the membrane region of PMA-primed HUVECs (Fig 4c). However, HUVECs routinely failed to react to fibronectin in the absence of PMA priming, suggesting that basal integrin activation is prerequisite for isoDGR motifs to influence endothelial β1 function.

**Figure 4:**
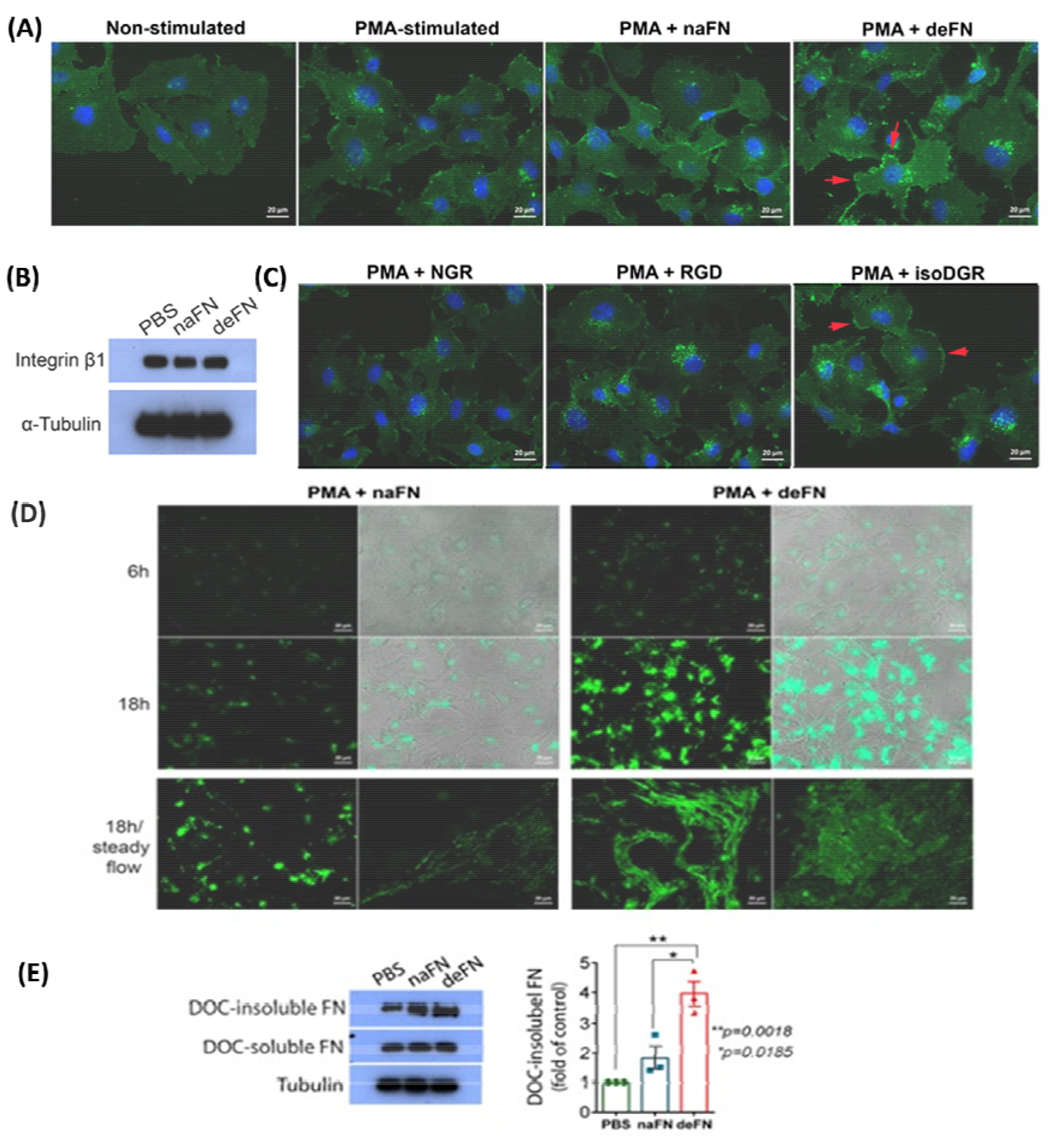
IsoDGR-modified fibronectin stimulated endothelial cell expression of integrin β1 and fibrillogenesis. **(A)** HUVEC endothelial cells treated with deamidated-fibronectin (25µg/ml) displayed higher levels of integrin β1 activation and distribution to the plasma membrane (red arrows) than were observed among control cells exposed to the native protein (n=3, scale bar 20 µm). **(B)** Total expression levels of integrin β1 were comparable between conditions upon assessment by western blot (n=3). **(C)** Activation of integrin β1 in these assays required the presence of peptides containing isoDGR motifs but not the alternative sequences NGR or RGD. Key: Native fibronectin, naFN; deamidated fibronectin, deFN (n=3, scale bar 20 µm). **(D)** PMA-primed HUVECs were incubated for 6h or 18h in the presence/absence of either native or deamidated fibronectin (FN) that had been pre-stained with AlexaFluor 488 dye. Accumulation of exogenous fibronectin on endothelial cell surfaces was markedly increased by deamidation (n=3, scale bar 20µm). When the same experiment was repeated under conditions of steady flow (100 rpm for 18h duration), deamidated fibronectin was observed to extend and form stretched fibrils associated with matrix assembly, whereas native protein displayed only limited accumulation on HUVEC monolayers (n=3, scale bar 20µm). **(E)** Upon completion of the steady-flow assay, HUVECs were treated with 1% DOC to assess matrix solubility, which confirmed that isoDGR-modified fibrils were highly resistant to detergent lysis (n=3 biological replicates). Quantification of western blot images was by Image J software. Statistical differences between groups were determined by unpaired T-test; *\*p=0*.*015, **p=0*.*0018*.

**Figure 5:**
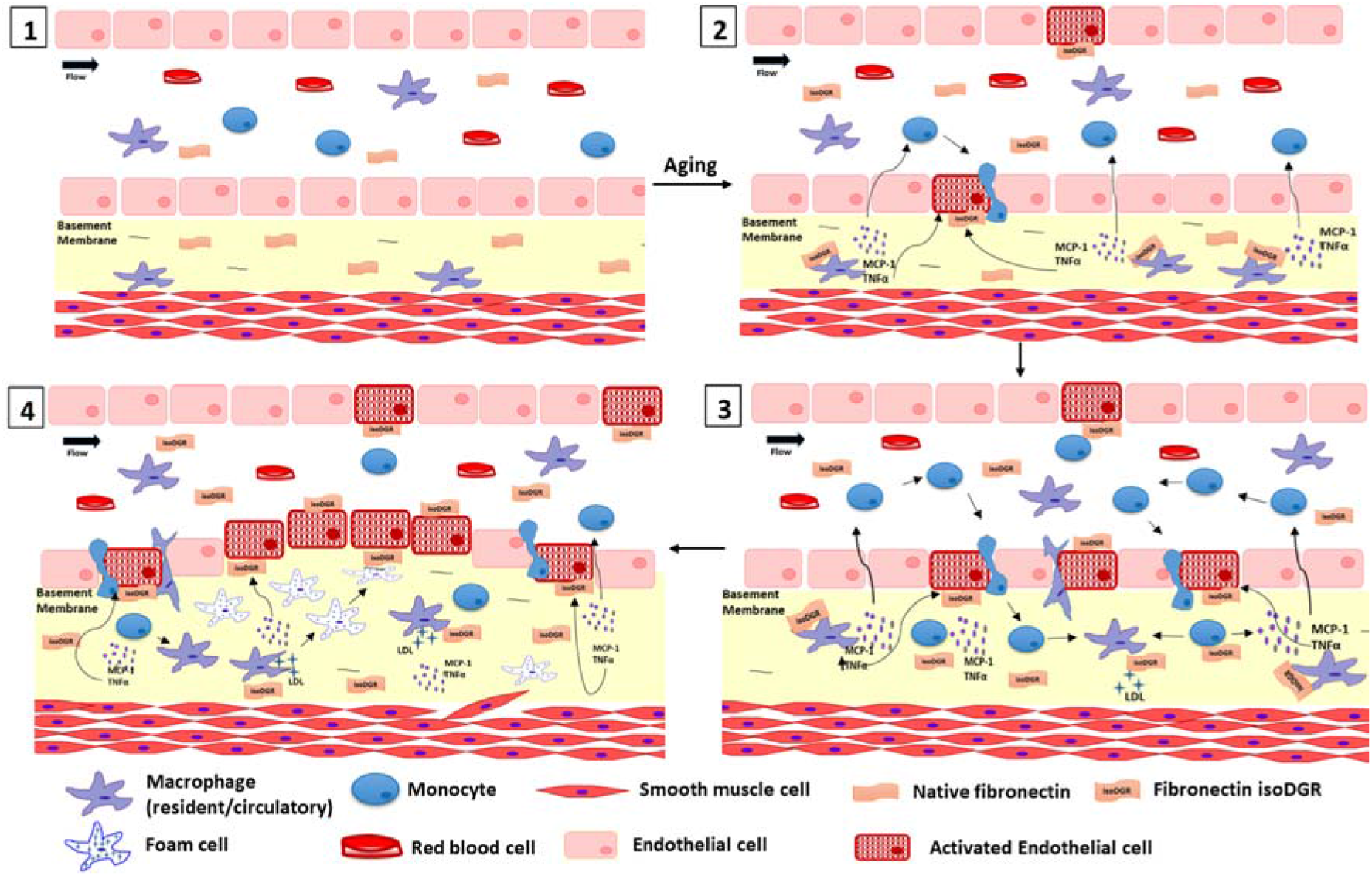
IsoDGR-modified fibronectin is a critical mediator of atherosclerotic CVD. Capacity to repair deamidated proteins declines during natural aging, leading to progressive accumulation of isoDGR-modified fibronectin in blood plasma and at the endothelial basement membrane. This age-damaged fibronectin activates tissue-resident macrophages and promotes expression of pro-inflammatory cytokines, particularly chemoattractants MCP-1 and TNFα which promote endothelial activation, vascular inflammation, and further monocyte recruitment. In addition, isoDGR-modified fibronectin can itself directly activate endothelial cells to enhance binding of circulating monocytes via upregulation of integrin β1. Protein ‘aging’ therefore triggers the early events of endothelial activation and leukocyte recruitment that are required for age-linked pathogenesis of atherosclerotic CVD.

### Fibronectin NGR-to-isoDGR transition facilitates fibrillogenesis and insoluble matrix formation

Fibronectin:integrin interactions initiate step-wise conformational changes in fibronectin:fibronectin interactions on cell surfaces that support conversion to insoluble fibrils / fibrillogenesis^34,35^. Since our data suggested that deamidated fibronectin can enhance activation of integrins - which are the principal fibronectin receptors in endothelial cells - we next examined whether protein deamidation was able to modify normal processes of polymerization, accumulation, and fibril formation that may lead to intima thickening. To do this, we incubated PMA-primed HUVECs for variable duration with either native or deamidated plasma fibronectin (25µg/ml) that had been pre-labelled with AlexaFluor488 dye. When visualized by microscopy, we rarely observed any deposition of native fibronectin on HUVECs after 6h exposure, and protein aggregation could only be detected after an extended 18h incubation period (Fig. 4d). In contrast, deposition of deamidated fibronectin was clearly visible within just 6h and protein aggregates on HUVECs were greatly increased over time. Since vascular endothelial cells are exposed to constant mechanical force *in vivo*,^36^ we next investigated the dynamics of native versus deamidated fibronectin assembly on PMA-primed HUVECs subjected to continuous flow (CO_2_ shaking incubator for 18h duration). Under these conditions, deposits of deamidated fibronectin converted to stretched and unfolded forms on endothelial cell surfaces, whereas native protein accumulated in greater quantities but failed to form extended fibrils, suggesting that deamidation of plasma fibronectin facilitates matrix assembly and formation of dense fibril networks (Fig. 4d). To further test whether isoDGR-containing fibronectin can accelerate the formation of stable and mature fibrillary networks, we next used a deoxycholate (DOC) detergent assay to assess how deamidated FN impacts matrix solubility, which is a critical determinant of overall tissue stability, rigidity, and shape^37,38^. Western blot analysis of proteins obtained by DOC buffer-extraction from HUVECs revealed a significant increase in insoluble fibronectin content among cells that had been exposed to isoDGR-containing variant relative to the unmodified protein (p=0.0185; Fig. 4e).

## Discussion

CVD is a major public health concern associated with an enormous socio-economic burden as both incidence and associated mortality continue to increase around the world^39^. Many CVD-related deaths could potentially be avoided by effective heath management after early detection of key risk factors. While it has long been established that CVD is a multifactorial disorder, aging remains the single largest risk factor for this complex disease. Over 70 years ago, the landmark Framingham Heart Study defined major risk factors for CVD which is driven by a complex interplay of age, genetics, smoking, nutrition, obesity, and other factors^40^. Progress in understanding the pathological mechanisms at play have led directly to the development of current anti-platelet, anti-thrombotic, anti-hypertensive, anti-diabetic, and anti-lipid drug therapies. Unfortunately, the mechanisms by which advancing age promotes atherosclerosis and intravascular thrombosis have remained elusive and the therapeutic opportunities these may provide are currently unknown. In the current study, we therefore sought to identify critical age-linked mediator of early events in atherosclerotic CVD.

Numerous recent studies have shown that biomolecule damage by spontaneous chemical reactions (i.e. degenerative protein modifications; DPMs) as a major contributor to age-linked diseases^13-16,41-44^. We have previously reported that specific matrix proteins in carotid atherosclerotic plaque from CVD patients exhibit marked deamidation of NGR motifs^5^, thus facilitating increased leukocyte binding to core matrix components^7^. In the current study, we generated a novel isoDGR-specific mAb enabling screening of blood plasma samples and functional studies of isoDGR biology in atherosclerotic CVD. To address the potential role of deamidated fibronectin in the vascular network and potential correlations with CVD pathology, we investigated effects of the isoDGR-modified protein on leukocytes and inflammation, as well as endothelial cell biology using a series of phenotypic and functional assays. In these experiments, isoDGR-modified fibronectin was consistently found to promote macrophage activation and monocyte recruitment/infiltration. Macrophages and monocytes can bind to ECM via integrin interactions with RGD or isoDGR motifs^7,10^. Our data now indicate that isoDGR:integrin signalling is a specific feature of the atherosclerotic matrix that triggers differential patterns of gene expression in adherent leukocytes, and that this process can be potently inhibited by isoDGR-specific mAb. Indeed, MAPK/ERK-dependent signalling and upregulation of transcription factor AP-1 were induced only in macrophage and monocytic cells binding to deamidated fibronectin, resulting in amplification of several chemokine/cytokine genes and transmigration of additional immune cells towards the assembling matrix. While AP-1 signal transduction is known to be activated by MAPK/ERK activation in response to a variety of external cues^30,45,46^, the mechanism by which the isoDGR:integrin:ERK:AP-1 axis induces signals in CVD will require further investigation. Intriguingly, isoDGR potently triggered macrophage secretion of MCP-1 and TNFα. Chemoattractant MCP-1 is a crucial mediator of monocyte/macrophage recruitment to the endothelial basement membrane during atherosclerosis development^47^, with MCP-1 knock-out animals displaying significant decreases in both plaque size and progression^47-49^. Another upregulated mediator TNF-α stimulates endothelial production of adhesion molecules that promotes binding of circulating monocytes^50^. The above results clearly indicate that age-damaged fibronectin which features isoDGR motifs can enhance binding to blood leukocytes and stimulate production of pro-inflammatory mediators MCP-1 and TNFα that drive recruitment of additional monocytic cells from the circulation. In summary, our data suggest that molecular switching from NGR to isoDGR motifs facilitates fibronectin binding to integrins on the surface of macrophages and monocytes leading to ‘outside-in’ signalling and induction of a range of potent pro-inflammatory mediators. Together, these events drive further leukocyte recruitment and accumulation in the artery wall, likely contributing to early pathological events in atherosclerotic CVD. Moreover, isoDGR motifs were also found to increase endothelial adhesion by upregulation of the integrin β1 subunit. Furthermore, isoDGR-modified fibronectin interacted with endothelial cell layers where the modified protein accelerated formation of stable, insoluble matrix that may contribute to intima thickening. These data suggest that fibronectin ‘aging’ generates isoDGR motifs that activate immune cells and endothelial cells via integrin receptor interactions.

Taken together, our results demonstrated that aged fibronectin which features isoDGR might initiate a vicious cycle driving the pathology of atherosclerotic CVD. Age-induced isoDGR activated resident macrophages to express/secrete proinflammatory cytokines and chemokines like MCP-1, thereby helping recruit yet more blood monocytes into the vascular wall. Moreover, both isoDGR stimulated proinflammatory cytokines (TNFα) and isoDGR itself can activate endothelial cells to enhance adhesion to circulating leukocytes. Upon interaction with isoDGR-modified fibronectin, infiltrated monocytes began differentiating into macrophages that are known to acquire LDL and form foam cells. This vicious cycle may repeat and amplify atherosclerotic plaque development and progression (Fig.5).

Our ELISA results revealed that CAD is associated with increased plasma levels of isoDGR-modified fibronectin and fibrinogen which are known to be critical mediators of arterial thrombosis. Moreover, the phenotypic and functional experiments suggest that isoDGR formation may be a key early event in the pathogenesis of atherosclerotic CVD that may be detected and accurately quantified using an isoDGR-specific mAb ELISA assay. Similar approaches could potentially also be used to develop routine blood tests for early detection of atherosclerotic CVD, thereby facilitating timely intervention to reduce mortality (e.g. via primary prevention strategies targeting modifiable risk factors like smoking/cholesterol/diet, or medical management with statins and blood thinners etc). However, further assessment of this possibility in larger cohorts and prospective studies will now be required since the control subjects used in the current study were limited to individuals with suspected CAD (despite subsequently diagnosis as non-CAD cases by angiography or CCTA). Our data also suggest that blocking/clearing isoDGR-motifs from the vasculature may be therapeutic and/or prophylactic in CVD and possibly also other age-linked diseases. Intriguingly, many ECM proteins with native ‘RGD’ integrin binding motif have ‘NGR’ sequence motifs susceptible to formation of isoDGR motifs during aging, and were detected in atherosclerotic plaque in our previous study^4-7^. Further investigation of these isoDGR-modified protein function and therapeutic targeting in both animal models and patient tissues should therefore be a priority for future studies.

## Data Availability

All data and results are provided as supplementary data

## Author contributions

J.E.P. and G.J. performed most *in-vitro* experiments, data analysis and drafted the manuscript; K.P. performed animal experiments and data analysis; X.G. performed LC-MS/MS; S.C.N. performed ERLIC-MS/MS analysis; K.C.K, S.S.N, S.E.L. performed qPCR and WB experiments; W.M. performed MALDI-TOF-MS analysis; S.C.L., M.K.L., D.J.P., D.K., V.S, H.H.H and N.E.M. contributed clinical samples, reagents and revised the manuscript; S.K.S. conceived and supervised the project and wrote the manuscript. All co-authors contributed to the revision of the manuscript.

## References

1. Benjamin, E. J., Virani, S. S., Callaway, C. W., Chamberlain, A. M., Chang, A. R., Cheng, S., Chiuve, S. E., Cushman, M., Delling, F. N., Deo, R., de Ferranti, S. D., Ferguson, J. F., Fornage, M., Gillespie, C., Isasi, C. R., Jimenez, M. C., Jordan, L. C., Judd, S. E., Lackland, D., Lichtman, J. H., Lisabeth, L., Liu, S., Longenecker, C. T., Lutsey, P. L., Mackey, J. S., Matchar, D. B., Matsushita, K., Mussolino, M. E., Nasir, K., O’Flaherty, M., Palaniappan, L. P., Pandey, A., Pandey, D. K., Reeves, M. J., Ritchey, M. D., Rodriguez, C. J., Roth, G. A., Rosamond, W. D., Sampson, U. K. A., Satou, G. M., Shah, S. H., Spartano, N. L., Tirschwell, D. L., Tsao, C. W., Voeks, J. H., Willey, J. Z., Wilkins, J. T., Wu, J. H., Alger, H. M., Wong, S. S. & Muntner, P. Heart Disease and Stroke Statistics-2018 Update: A Report From the American Heart Association. Circulation 137, e67–e492, (2018).

2. Herrington, W., Lacey, B., Sherliker, P., Armitage, J. & Lewington, S. Epidemiology of Atherosclerosis and the Potential to Reduce the Global Burden of Atherothrombotic Disease. Circulation research 118, 535–46, (2016).

3. Bäck, M., Yurdagul, A., Tabas, I., Öörni, K. & Kovanen, P. T. Inflammation and its resolution in atherosclerosis: mediators and therapeutic opportunities. Nat. Rev. Cardiol. 16, 389–406, (2019).

4. Cheow, E. S. H., Hao, P. L., Hao, P. L., Sorokin, V., Lee, C. N., Kleijn, D. P. V. & Sze, S. K. The Role of Protein Deamidation in Cardiovascular Disease. Proceedings of the 23rd American Peptide Symposium 17, 212–13, (2013).

5. Hao, P., Ren, Y., Pasterkamp, G., Moll, F. L., de Kleijn, D. P. & Sze, S. K. Deep proteomic profiling of human carotid atherosclerotic plaques using multidimensional LC-MS/MS. Proteomics. Clinical applications 8, 631–5, (2014).

6. Bleijerveld, O. B., Zhang, Y.-N., Beldar, S., Hoefer, I. E., Sze, S. K., Pasterkamp, G. & de Kleijn, D. P. V. Proteomics of plaques and novel sources of potential biomarkers for atherosclerosis. PROTEOMICS – Clinical Applications 7, 490–503, (2013).

7. Dutta, B., Park, J. E., Kumar, S., Hao, P., Gallart-Palau, X., Serra, A., Ren, Y., Sorokin, V., Lee, C. N., Ho, H. H., de Kleijn, D. & Sze, S. K. Monocyte adhesion to atherosclerotic matrix proteins is enhanced by Asn-Gly-Arg deamidation. Scientific Reports 7, 5765, (2017).

8. Corti, A. & Curnis, F. Isoaspartate-dependent molecular switches for integrin-ligand recognition. Journal of cell science 124, 515–22, (2011).

9. Spitaleri, A., Mari, S., Curnis, F., Traversari, C., Longhi, R., Bordignon, C., Corti, A., Rizzardi, G. P. & Musco, G. Structural basis for the interaction of isoDGR with the RGD-binding site of alphavbeta3 integrin. The Journal of biological chemistry 283, 19757–68, (2008).

10. Curnis, F., Longhi, R., Crippa, L., Cattaneo, A., Dondossola, E., Bachi, A. & Corti, A. Spontaneous Formation of L-Isoaspartate and Gain of Function in Fibronectin. Journal of Biological Chemistry 281, 36466–76, (2006).

11. Geiger, T. & Clarke, S. Deamidation, isomerization, and racemization at asparaginyl and aspartyl residues in peptides. Succinimide-linked reactions that contribute to protein degradation. The Journal of biological chemistry 262, 785–94, (1987).

12. Hao, P., Adav, S. S., Gallart-Palau, X. & Sze, S. K. Recent advances in mass spectrometric analysis of protein deamidation. Mass spectrometry reviews 36, 677–92, (2017).

13. Gallart-Palau, X., Tan, L. M., Serra, A., Gao, Y., Ho, H. H., Richards, A. M., Kandiah, N., Chen, C. P., Kalaria, R. N. & Sze, S. K. Degenerative protein modifications in the aging vasculature and central nervous system: A problem shared is not always halved. Ageing research reviews 53, 100909, (2019).

14. Adav, S. S. & Sze, S. K. Hypoxia-Induced Degenerative Protein Modifications Associated with Aging and Age-Associated Disorders. Aging and disease 11, 341–64, (2020).

15. Gallart-Palau, X., Serra, A. & Sze, S. K. Uncovering Neurodegenerative Protein Modifications via Proteomic Profiling. International review of neurobiology 121, 87–116, (2015).

16. Truscott, R. J. W., Schey, K. L. & Friedrich, M. G. Old Proteins in Man: A Field in its Infancy. Trends in biochemical sciences 41, 654–64, (2016).

17. Robinson, N. E. & Robinson, A. B. Molecular clocks. Proceedings of the National Academy of Sciences 98, 944–49, (2001).

18. Rohwedder, I., Montanez, E., Beckmann, K., Bengtsson, E., Dunér, P., Nilsson, J., Soehnlein, O. & Fässler, R. Plasma fibronectin deficiency impedes atherosclerosis progression and fibrous cap formation. EMBO molecular medicine 4, 564–76, (2012).

19. Farhadian, F., Contard, F., Sabri, A., Samuel, J. L. & Rappaport, L. Fibronectin and basement membrane in cardiovascular organogenesis and disease pathogenesis. Cardiovascular research 32, 433–42, (1996).

20. Young, L., Sung, J., Stacey, G. & Masters, J. R. Detection of Mycoplasma in cell cultures. Nature protocols 5, 929–34, (2010).

21. Wierzbicka-Patynowski, I., Mao, Y. & Schwarzbauer, J. E. Analysis of Fibronectin Matrix Assembly. Current Protocols in Cell Biology 25, 10.12.1-10.12.10, (2004).

22. Kim, E., Lowenson, J. D., Clarke, S. & Young, S. G. Phenotypic analysis of seizure-prone mice lacking L-isoaspartate (D-aspartate) O-methyltransferase. The Journal of biological chemistry 274, 20671–8, (1999).

23. Cho, J. & Mosher, D. F. Enhancement of thrombogenesis by plasma fibronectin cross-linked to fibrin and assembled in platelet thrombi. Blood 107, 3555–63, (2006).

24. Pereira, M., Rybarczyk, B. J., Odrljin, T. M., Hocking, D. C., Sottile, J. & Simpson-Haidaris, P. J. The incorporation of fibrinogen into extracellular matrix is dependent on active assembly of a fibronectin matrix. Journal of cell science 115, 609–17, (2002).

25. Hao, P., Qian, J., Dutta, B., Cheow, E. S., Sim, K. H., Meng, W., Adav, S. S., Alpert, A. & Sze, S. K. Enhanced separation and characterization of deamidated peptides with RP-ERLIC-based multidimensional chromatography coupled with tandem mass spectrometry. Journal of proteome research 11, 1804–11, (2012).

26. Serra, A., Gallart-Palau, X., Wei, J. & Sze, S. K. Characterization of Glutamine Deamidation by Long-Length Electrostatic Repulsion-Hydrophilic Interaction Chromatography-Tandem Mass Spectrometry (LERLIC-MS/MS) in Shotgun Proteomics. Analytical chemistry 88, 10573–82, (2016).

27. Hao, P., Ren, Y., Alpert, A. J. & Sze, S. K. Detection, evaluation and minimization of nonenzymatic deamidation in proteomic sample preparation. Molecular & cellular proteomics: MCP 10, O111.009381, (2011).

28. Sze, S. K., JebaMercy, G. & Ngan, S. C. Profiling the ‘deamidome’ of complex biosamples using mixed-mode chromatography-coupled tandem mass spectrometry. Methods (San Diego, Calif.), (2020).

29. Liu, T., Zhang, L., Joo, D. & Sun, S.-C. NF-κB signaling in inflammation. Signal Transduct Target Ther 2, 17023, (2017).

30. Qiao, Y., He, H., Jonsson, P., Sinha, I., Zhao, C. & Dahlman-Wright, K. AP-1 Is a Key Regulator of Proinflammatory Cytokine TNFα-mediated Triple-negative Breast Cancer Progression. Journal of Biological Chemistry 291, 5068–79, (2016).

31. Cahill, C. M., Zhu, W., Oziolor, E., Yang, Y.-J., Tam, B., Rajanala, S., Rogers, J. T. & Walker, W. A. Differential Expression of the Activator Protein 1 Transcription Factor Regulates Interleukin-1ß Induction of Interleukin 6 in the Developing Enterocyte. PLOS ONE 11, e0145184, (2016).

32. Davis, G. E. & Senger, D. R. Endothelial Extracellular Matrix. Circulation research 97, 1093–107, p(2005).

33. Luque, A., Gómez, M., Puzon, W., Takada, Y., Sánchez-Madrid, F. & Cabañas, C. Activated Conformations of Very Late Activation Integrins Detected by a Group of Antibodies (HUTS) Specific for a Novel Regulatory Region(355425) of the Common 1 Chain. Journal of Biological Chemistry 271, 11067–75, (1996).

34. Schwarzbauer, J. E. & Sechler, J. L. Fibronectin fibrillogenesis: a paradigm for extracellular matrix assembly. Current opinion in cell biology 11, 622–7, (1999).

35. Wierzbicka-Patynowski, I. & Schwarzbauer, J. E. The ins and outs of fibronectin matrix assembly. Journal of cell science 116, 3269–76, (2003).

36. Chistiakov, D. A., Orekhov, A. N. & Bobryshev, Y. V. Effects of shear stress on endothelial cells: go with the flow. Acta Physiol 219, 382–408, (2017).

37. McKeown-Longo, P. J. & Mosher, D. F. Binding of plasma fibronectin to cell layers of human skin fibroblasts. The Journal of cell biology 97, 466–72, (1983).

38. Gattazzo, F., Urciuolo, A. & Bonaldo, P. Extracellular matrix: A dynamic microenvironment for stem cell niche. Biochim Biophys Acta 1840, 2506–19, (2014).

39. Roth, G. A., Johnson, C., Abajobir, A., Abd-Allah, F., Abera, S. F., Abyu, G., Ahmed, M., Aksut, B., Alam, T., Alam, K., Alla, F., Alvis-Guzman, N., Amrock, S., Ansari, H., Ärnlöv, J., Asayesh, H., Atey, T. M., Avila-Burgos, L., Awasthi, A., Banerjee, A., Barac, A., Bärnighausen, T., Barregard, L., Bedi, N., Belay Ketema, E., Bennett, D., Berhe, G., Bhutta, Z., Bitew, S., Carapetis, J., Carrero, J. J., Malta, D. C., Castañeda-Orjuela, C. A., Castillo-Rivas, J., Catalá-López, F., Choi, J.-Y., Christensen, H., Cirillo, M., Cooper, L., Criqui, M., Cundiff, D., Damasceno, A., Dandona, L., Dandona, R., Davletov, K., Dharmaratne, S., Dorairaj, P., Dubey, M., Ehrenkranz, R., El Sayed Zaki, M., Faraon, E. J. A., Esteghamati, A., Farid, T., Farvid, M., Feigin, V., Ding, E. L., Fowkes, G., Gebrehiwot, T., Gillum, R., Gold, A., Gona, P., Gupta, R., Habtewold, T. D., Hafezi-Nejad, N., Hailu, T., Hailu, G. B., Hankey, G., Hassen, H. Y., Abate, K. H., Havmoeller, R., Hay, S. I., Horino, M., Hotez, P. J., Jacobsen, K., James, S., Javanbakht, M., Jeemon, P., John, D., Jonas, J., Kalkonde, Y., Karimkhani, C., Kasaeian, A., Khader, Y., Khan, A., Khang, Y.-H., Khera, S., Khoja, A. T., Khubchandani, J., Kim, D., Kolte, D., Kosen, S., Krohn, K. J., Kumar, G. A., Kwan, G. F., Lal, D. K., Larsson, A., Linn, S., Lopez, A., Lotufo, P. A., El Razek, H. M. A., Malekzadeh, R., Mazidi, M., Meier, T., Meles, K. G., Mensah, G., Meretoja, A., Mezgebe, H., Miller, T., Mirrakhimov, E., Mohammed, S., Moran, A. E., Musa, K. I., Narula, J., Neal, B., Ngalesoni, F., Nguyen, G., Obermeyer, C. M., Owolabi, M., Patton, G., Pedro, J., Qato, D., Qorbani, M., Rahimi, K., Rai, R. K., Rawaf, S., Ribeiro, A., Safiri, S., Salomon, J. A., Santos, I., Santric Milicevic, M., Sartorius, B., Schutte, A., Sepanlou, S., Shaikh, M. A., Shin, M.-J., Shishehbor, M., Shore, H., Silva, D. A. S., Sobngwi, E., Stranges, S., Swaminathan, S., Tabarés-Seisdedos, R., Tadele Atnafu, N., Tesfay, F., Thakur, J. S., Thrift, A., Topor-Madry, R., Truelsen, T., Tyrovolas, S., Ukwaja, K. N., Uthman, O., Vasankari, T., Vlassov, V., Vollset, S. E., Wakayo, T., Watkins, D., Weintraub, R., Werdecker, A., Westerman, R., Wiysonge, C. S., Wolfe, C., Workicho, A., Xu, G., Yano, Y., Yip, P., Yonemoto, N., Younis, M., Yu, C., Vos, T., Naghavi, M. & Murray, C. Global, Regional, and National Burden of Cardiovascular Diseases for 10 Causes, 1990 to 2015. Journal of the American College of Cardiology 70, 1–25, (2017).

40. Andersson, C., Johnson, A. D., Benjamin, E. J., Levy, D. & Vasan, R. S. 70-year legacy of the Framingham Heart Study. Nature reviews. Cardiology 16, 687–98, (2019).

41. Guo, X., Park, J. E., Gallart-Palau, X. & Sze, S. K. Oxidative Damage to the TCA Cycle Enzyme MDH1 Dysregulates Bioenergetic Enzymatic Activity in the Aged Murine Brain. Journal of proteome research 19, 1706–17, (2020).

42. Gallart-Palau, X., Serra, A., Lee, B. S. T., Guo, X. & Sze, S. K. Brain ureido degenerative protein modifications are associated with neuroinflammation and proteinopathy in Alzheimer’s disease with cerebrovascular disease. Journal of neuroinflammation 14, 175, (2017).

43. Adav, S. S. & Sze, S. K. Insight of brain degenerative protein modifications in the pathology of neurodegeneration and dementia by proteomic profiling. Molecular brain 9, 92, (2016).

44. Hipp, M. S., Kasturi, P. & Hartl, F. U. The proteostasis network and its decline in ageing. Nature reviews. Molecular cell biology 20, 421–35, (2019).

45. Hu, Y., Cheng, L., Hochleitner, B.-W. & Xu, Q. Activation of Mitogen-Activated Protein Kinases (ERK/JNK) and AP-1 Transcription Factor in Rat Carotid Arteries After Balloon Injury. Arteriosclerosis, Thrombosis, and Vascular Biology 17, 2808–16, (1997).

46. Karin, M. The Regulation of AP-1 Activity by Mitogen-activated Protein Kinases. Journal of Biological Chemistry 270, 16483–86, (1995).

47. Georgakis, M. K., Gill, D., Rannikmäe, K., Traylor, M., Anderson, C. D., Lee, J. M., Kamatani, Y., Hopewell, J. C., Worrall, B. B., Bernhagen, J., Sudlow, C. L. M., Malik, R. & Dichgans, M. Genetically Determined Levels of Circulating Cytokines and Risk of Stroke. Circulation 139, 256–68, (2019).

48. Boring, L., Gosling, J., Cleary, M. & Charo, I. F. Decreased lesion formation in CCR2−/−mice reveals a role for chemokines in the initiation of atherosclerosis. Nature 394, 894–7, (1998).

49. Aslanian, A. M. & Charo, I. F. Targeted disruption of the scavenger receptor and chemokine CXCL16 accelerates atherosclerosis. Circulation 114, 583–90, (2006).

50. Mackay, F., Loetscher, H., Stueber, D., Gehr, G. & Lesslauer, W. Tumor necrosis factor alpha (TNF-alpha)-induced cell adhesion to human endothelial cells is under dominant control of one TNF receptor type, TNF-R55. Journal of Experimental Medicine 177, 1277–86, (1993).

