## Supplementary methods for "Aging-induced isoDGR-modified fibronectin activates monocytic and endothelial cells to promote atherosclerosis"

**Supplementary materials and methods**

***Reagents, peptides, and antibodies***

Human plasma fibronectin was purchased from Advanced Biomatrix (San Diego, CA, USA). 5-Carboxyfluorescein N-Succinimidyl ester (CFSE) fluorescent dye was obtained from Sigma-Aldrich. AlexaFluor-488-microscale labelling kit was from ThermoFisher Scientific. The following peptides were produced by GL Biochem Ltd. (Shanghai, China); GC(isoD)GRCGG-(CH2-CH2-NH2), GCDGRCGG-(CH2-CH2-NH2), GCNGRCGG-(CH2-CH2-NH2), and GCRGDCGG-(CH2-CH2-NH2). The molecular mass of each peptide was confirmed by MALDI-TOF mass spectrometry (MS) analysis. Antibodies against p44/42 MAPK (ERK1/2), phospho-p44/42 MAPK, and integrin β3 were purchased from Cell Signalling Technologies (Danvers, MA, USA). Antibodies against fibronectin, tubulin, and actin were from Santa Cruz Biotechnology (Santa Cruz, CA, USA) and anti-integrin β1 antibody (clone HUTS-4) was obtained from Chemicon (Merck Millipore, Billerica, MA). Rabbit polyclonal antibodies to fibronectin and fibrinogen were from Abcam (Cambridge, United Kingdom). SEAP reporter construct was purchased from BD Clontech (San Jose, CA, USA) and Phospha‐Light™ was from Applied Biosystems (Bedford, MA).

***Generation of monoclonal antibody against isoDGR motifs***

Hybridoma cell line that secretes isoDGR-specific monoclonal antibody motif was generated by GenScript Corporation (Piscataway, NJ, USA). Two synthetic peptides, C(isoD)GRCK and (isoD)GR conjugated to keyhole limpet hemocyanin (KLH) were used to immunize Balb/c mice. Positive clones were selected by a series of dot blot and ELISA screenings against the isoDGR-containing antigens. We then used a combination of peptide-based ELISA and dot blot assays to test reactivity of the selected hybridoma cells against four peptides containing either isoDGR or the dummy sequences DGR, RGD, and NGR. isoDGR-specific antibodies in hybridoma culture supernatants were determined by indirect ELISA (Fig. 1a). Briefly, Nunc Maxisorp 96 well plates were coated overnight at 4°C with 100 µL/well of each of the peptides (2 µg/mL), and blocked with 5% BSA. The culture supernatants (1:500) were added and incubated at 37°C for 2 hr before adding the secondary antibody. The isoDGR motifs were detected by using horseradish peroxidase (HRP) goat anti-mouse antibodies (R&D systems, Minneapolis, USA) and TMB as a substrate.  Absorbance was read at 450nm. Based on the results of these assays, we selected a single hybridoma that displayed high specificity for the isoDGR motif but lacked reactivity to DGR, RGD, or NGR (Fig. 1a). Monoclonal antibody was purified from the supernatant of hybridoma cell cultures using protein A/G agarose columns according to the manufacturer’s instructions (Thermo Fisher Scientific, San Jose, USA).

***Indirect and sandwich ELISA for isoDGR detection in human plasma samples***

The same method was followed as described previously in peptide ELISA method (SFig. 1a). Briefly, total of 5µg plasma protein in 100µl PBS was coated into 96-well plate and incubated at 4°C overnight. Mouse monoclonal anti-isoDGR, FN or FBG antibody (5µg/ml) and the secondary anti-mouse IgG antibody conjugated to horseradish peroxidase (Pierce, Rockford, IL) was used to detect total FN, FBG or deamidated NGR motifs. To enable rapid detection of isoDGR motifs in FN and FBG from human plasma samples, we next developed a sandwich ELISA approach using our custom antibody (SFig. 1b). A total volume of 100µl anti-isoDGR mAb was added to each well for overnight incubation at 4°C, after which the wells were washed three times with 200µl PBS. Non-specific binding sites were blocked using 3% BSA in PBS for 1h at RT. After washing, 100µl samples were added to each well and incubated for 2h at RT. Following washing steps, rabbit polyclonal antibodies against FN or FBG were added and incubated for 2h at RT. After washing with PBST, the HRP-coupled donkey mAb anti-rabbit antibodies (Cell Signalling) was added for 1hr at RT. the wells were washed 3 times with PBST and 100µl TMB was added to each well for 10min to enable colour development. To stop the colour reaction, 100µl 2N H_2_SO_4_ was added before reading optical density at 450nm.

***Fibronectin deamidation and tandem mass spectrometry***

Two forms of plasma FN (native and deamidated) were used in the experiments. Native FN was dissolved in ammonium acetate buffer at pH 6.5, while deamidated FN was prepared by incubating FN in 0.1M ammonium bicarbonate buffer (pH 8.0) for 48h at 37°C before adjusting to pH 6.5 by addition of 1% acetic acid. Native and deamidated FN structures were confirmed by LC-MS/MS and then used for cell treatment or plate coating. A 20μg mass of FN in solution (pH 6.5) was reduced with 10mM dithiothreitol for 2h at RT, then alkylated using 55mM iodoacetamide for 1h at RT in the dark. The sample was then incubated at 37°C for overnight trypsin digestion (trypsin 1:50 protein w/w ratio; Promega, Madison, WI). The digestion reaction was quenched by adding formic acid (FA) until the final acid concentration reached 0.5%. Tryptic peptides were dried in a vacuum concentrator (Concentrator Plus, Eppendorf AG, Hamburg, Germany). The FN tryptic peptides were analysed by RPLC-MS/MS using C18 column and ERLIC-MS/MS using WAX column as described previously ^1-3^. In RPLC-MS/MS experiment, the peptide samples were reconstituted in 3% acetonitrile (ACN), 0.1% FA buffer, and then injected into a Dionex Ultimate 3000 RSLCnano HPLC (Thermo Fisher Scientific, San Jose, CA, USA). Peptide separation was performed on a Dionex EASY-Spray 75 μm x 10 cm column packed with PepMap C18 3μm, 100 Å (Thermo Fisher Scientific, Inc). Separation of peptides was performed with mobile phase solvent A (0.1% FA in HPLC water) and solvent B (0.1% FA in 90% ACN) at flow rate of 300nl/min with a 60min gradient. The sample was sprayed with an EASY nanospray source (ThermoFisher Scientific, Inc.) at an electrospray potential of 1.5 kV. In ERLIC-MS/MS run, the peptide samples were dissolved in 90% ACN, 0.1% acetic acid (AA) buffer, and injected into an in-house packed 200 μm x 10 cm WAX column (Bulk material: PolyWAX 3 μm, 300 Å particles) using a Vanquish HPLC (Thermo Fisher Scientific, Inc.). ERLIC separation of peptides was performed with mobile phase solvent A (90% ACN, 0.1%AA) and solvent B (0.1% FA in HPLC water) at flow rate of 3μl/min with a 60min gradient. Peptide ionization was through a ESI source (ThermoFisher Scientific, Inc.) at an electrospray voltage of 3.5 kV. Both RPLC- and ERLIC- separation were coupled to a Q Exactive MS instrument for LC-MS/MS analysis. The Q Exactive was set to perform data acquisition in positive ion mode and a full MS scan (350-1,600 m/z range) was acquired at a resolution of 70,000 at m/z 200. The 10 most intense ions were selected for higher energy collisional dissociation (HCD) fragmentation using 28% normalized collision energy. The AGC setting was 1E6 for the full MS scan and 2E5 for the MS2 scan. Single and unassigned charged ions were excluded from MS2.

***Label-free proteomic data analysis***

Raw data files were converted into the mascot generic file format (MGF) using Proteome Discoverer version 1.4 (Thermo Electron, Bremen, Germany) with the MS2 spectrum processor for de-isotoping and de-convoluting MS/MS spectra. Label-free raw data file searches were carried out using an in-house Mascot server (version 2.6.02, Matrix Science, MA) with precursor ion tolerance of 5ppm and fragment ion tolerance of 30ppm ^3^. The UniProt human database was used for protein database searches (downloaded on June 23, 2015, including 180,822 sequences and 71,773,890 residues). Variable modifications were set as deamidation of N and Q, as well as oxidation of M, while carbamidomethylation of C was set as a fixed modification. Two missed trypsin cleavage sites per peptide were tolerated. The area under the extracted ion chromatogram (XIC) curve is used to determine the peptide abundance in the samples. The XIC was integrated over the calculated m/z of the precursor ions with +/- 3ppm tolerance.

***Quantitative real-time PCR***

Quantitative real-time PCR (qRT‐PCR) was used to measure mRNA expression levels of cytokines by U937 and THP-1 monocytic cells and RAW macrophage cells. Total RNA extraction was performed using Nucleospin RNA kits (MACHEREY-NAGEL GmbH & Co.) according to the manufacturer’s protocol. qRT‐PCR was performed using a CFX96 Real-Time PCR Detection System (Bio-Rad) with KAPA SYBR® FAST qPCR Master Mix. To normalize quantification, levels of 18s RNA detected on the same plate as the target genes were used as internal controls. The primers of RT-PCR were listed in Supplementary Data 7 (STable 3). The qRT‐PCR reaction was as follows; denaturation at 95°C for 15s followed by annealing at 58°C for 15s and extension at 72°C for 15s then final extension at 72°C for 5min over a total of 40 cycles.

***Western blot analysis***

Cells were washed in ice-cold PBS and lysed in modified RIPA buffer (50mM Tris–HCl, 150mM NaCl, 1% NP-40, pH 8.0, 1 × protease inhibitor cocktail, phosphatase inhibitors). Lysates were clarified by centrifugation (16,000 × g, 30min) and subjected to western blotting using the indicated primary antibodies at 1:1000 dilution. Protein-antibody conjugates were visualized using a chemiluminescence detection kit (Thermo Fisher Scientific). The original (whole) images of the Western blot is showed in Supplementary figure 5.

***Reference***

1.	Serra, A., Gallart-Palau, X., Wei, J. & Sze, S. K. Characterization of Glutamine Deamidation by Long-Length Electrostatic Repulsion-Hydrophilic Interaction Chromatography-Tandem Mass Spectrometry (LERLIC-MS/MS) in Shotgun Proteomics. *Anal Chem* **88**, 10573-82, (2016).

2.	Hao, P., Qian, J., Dutta, B., Cheow, E. S., Sim, K. H., Meng, W., Adav, S. S., Alpert, A. & Sze, S. K. Enhanced separation and characterization of deamidated peptides with RP-ERLIC-based multidimensional chromatography coupled with tandem mass spectrometry. *J Proteome Res* **11**, 1804-11, (2012).

3.	Hao, P., Ren, Y., Alpert, A. J. & Sze, S. K. Detection, evaluation and minimization of nonenzymatic deamidation in proteomic sample preparation. *Mol Cell Proteomics* **10**, O111.009381, (2011).
