## Supplementary figures for "Aging-induced isoDGR-modified fibronectin activates monocytic and endothelial cells to promote atherosclerosis"

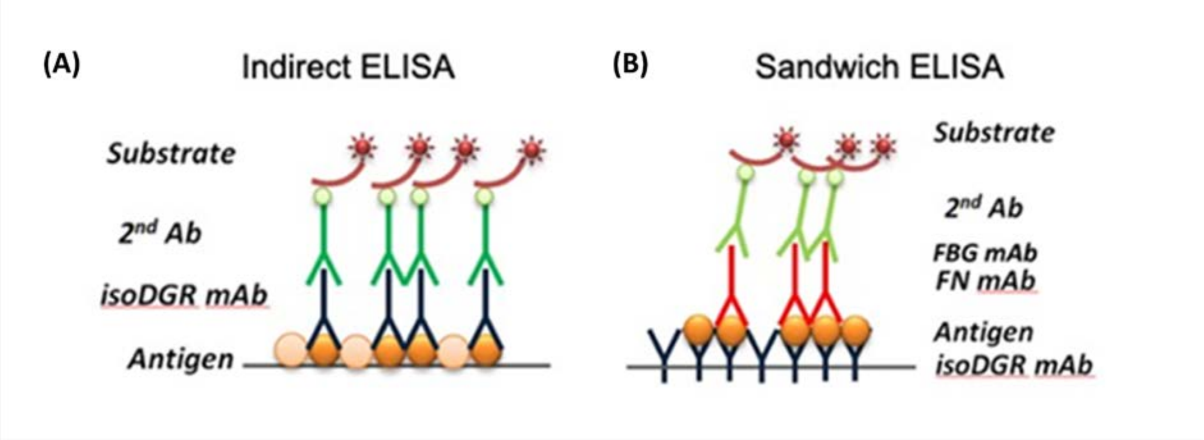


**Supplementary figure 1: Schematic diagram of the indirect ELISA and sandwich ELISA methods (A)** Indirect ELISA method used to measure total isoDGR-modified protein levels in CAD patient plasma. **(B)** Schematic diagram of the sandwich ELISA technique used to determine isoDGR modifcation of fibronectin (FN) and fibrinogen (FBG) proteins.

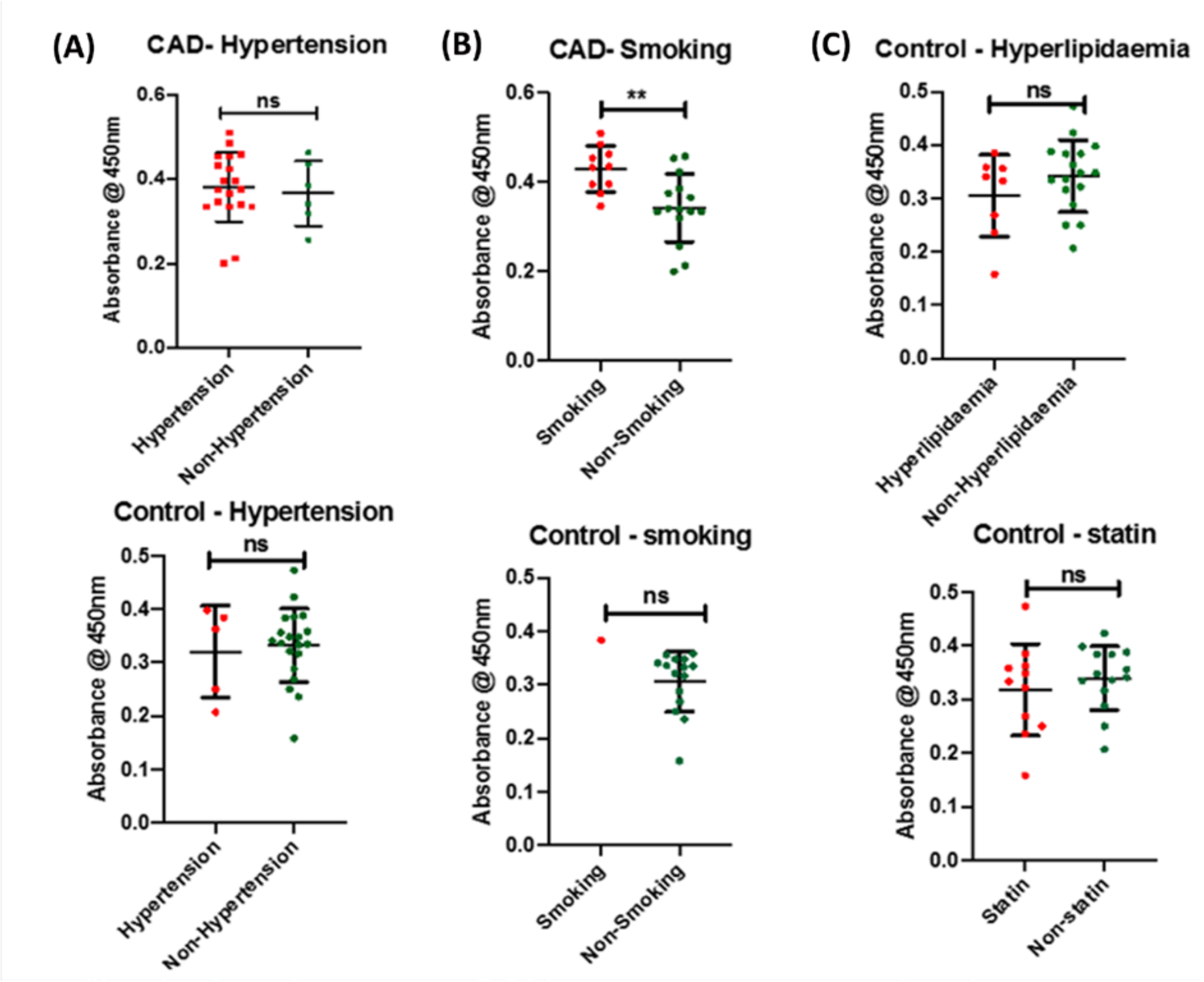


**Supplementary file 2**: **Effect of clinical/lifestyle features in plasma isoDGR levels** Total isoDGR levels in blood plasma from CAD patients and controls were subject to sub-analyses according to clinical data: (A) Hypertension; **(B)** Smoking; **(C)** Lipid level and statin therapy. Results showed that smoking status (but not other patient features) was associated with elevated concentration of isoDGR-modified plasma proteins. Correlation of isoDGR levels with hyperlipidemia and statin therapy were determined only in CTRL donors (since all CAD patients have a background of hyperlipidemia and statin treatment).

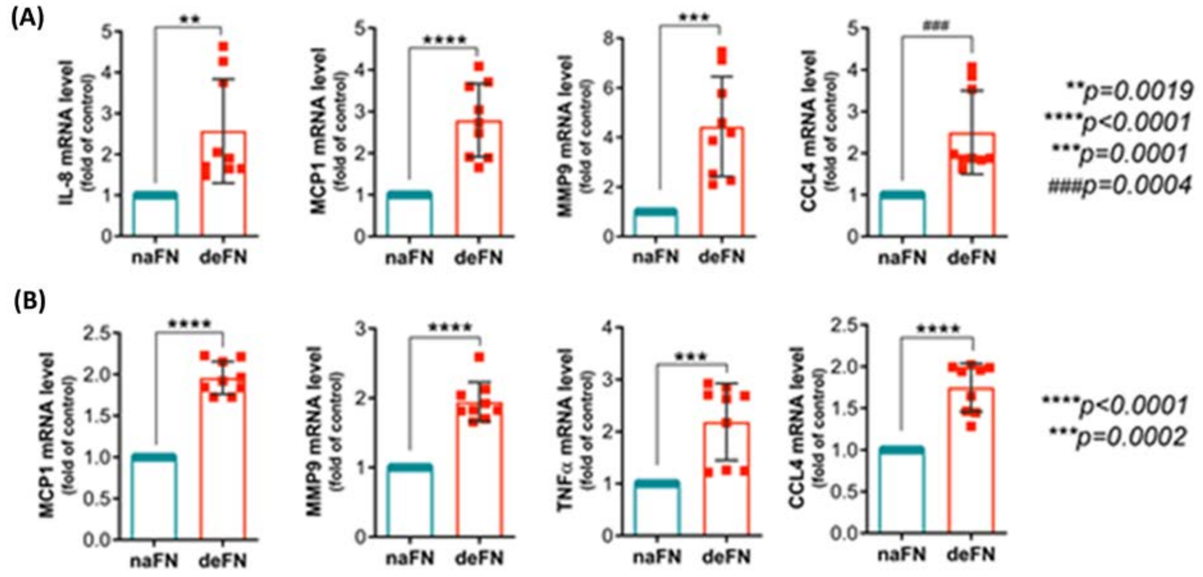


**Supplementary figure 3: IsoDGR-modified fibronectin induces monocyte expression of pro-inflammatory cytokine/chemokines. (A)** Monocytic U937 cells  or **(B)** THP-1 cells were primed with PMA then cultured for 24h together with native or deamidated fibronectin before mRNA expression levels of key cytokines were assessed by qRT-PCR. This analysis revealed that monocytes exposed to isoDGR-modified fibronectin displayed significant upregulation of several chemotactic molecules, pro-inflammatory cytokines, and metalloproteinase enzyme MMP9. Experiments included n=3 biological replicates (3 technical repeats per biological replicate, data shown are mean ± S.E. and were assessed by unpaired T-test). Statistical differences between groups were determined by unpaired T-test.

**Supplementary figure 4: isoDGR-fibronectin promotes monocytes migration and infiltration** Transwell assay shown in Figure 3 was repeated with addition of a neutralizing antibody control. **(A)** Levels of monocytes infiltration in PBS, native fibronectin, deamidated fibronectin and deamidation fibronectin with neutralizing isoDGR-specific mAb; **(B)** image of infiltrated monocytes in PBS control condition; **(C)** image of infiltrated monocytes in native fibronectin; **(D)** image of infiltrated monocytes in deamidated fibronectin; **(E)** image of infiltrated monocytes in deamidated fibronectin in the presence of neutralizing isoDGR mAb. Results showed that adhesion of U937 cells to deamidated fibronectin (bottom chamber) induced greater transmigration of unstimulated monocytes (matrigel-coated top chamber) than did exposure to the native protein control (assessed by microscopic or colorimetric analysis after 24h incubation). Experiments included n=3 biological replicates. Statistical differences between groups were determined by one way ANOVA.

A

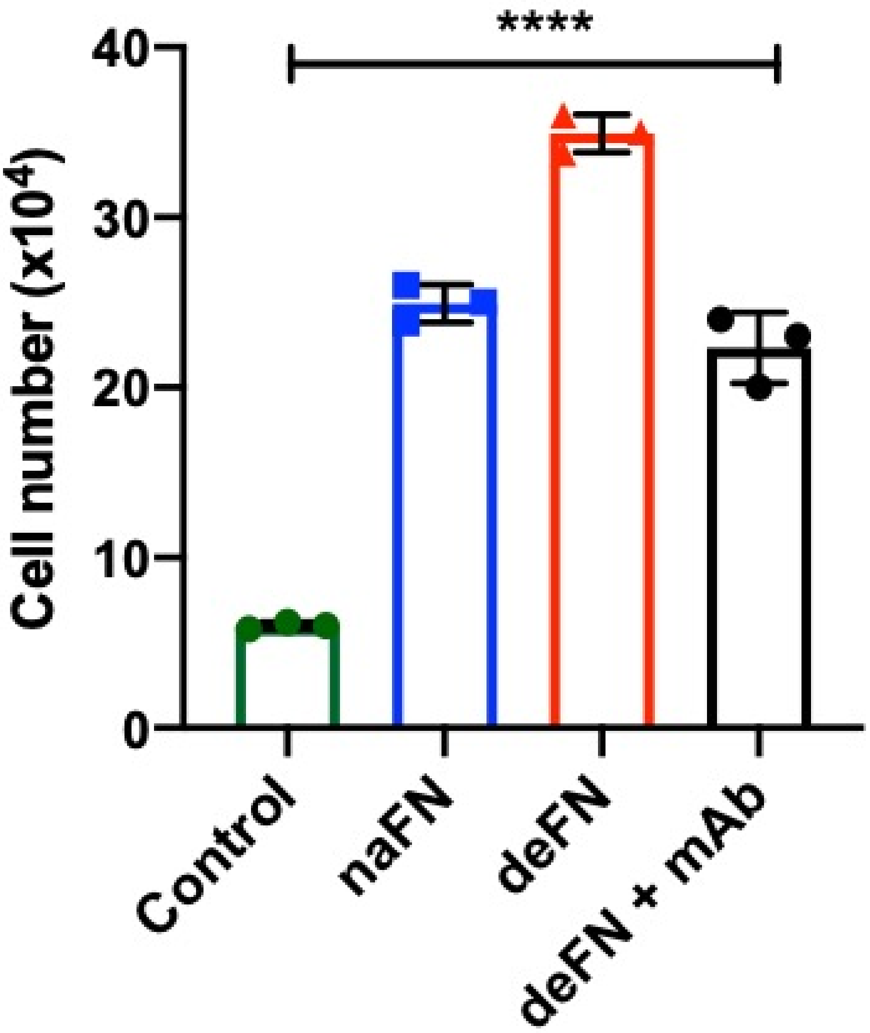

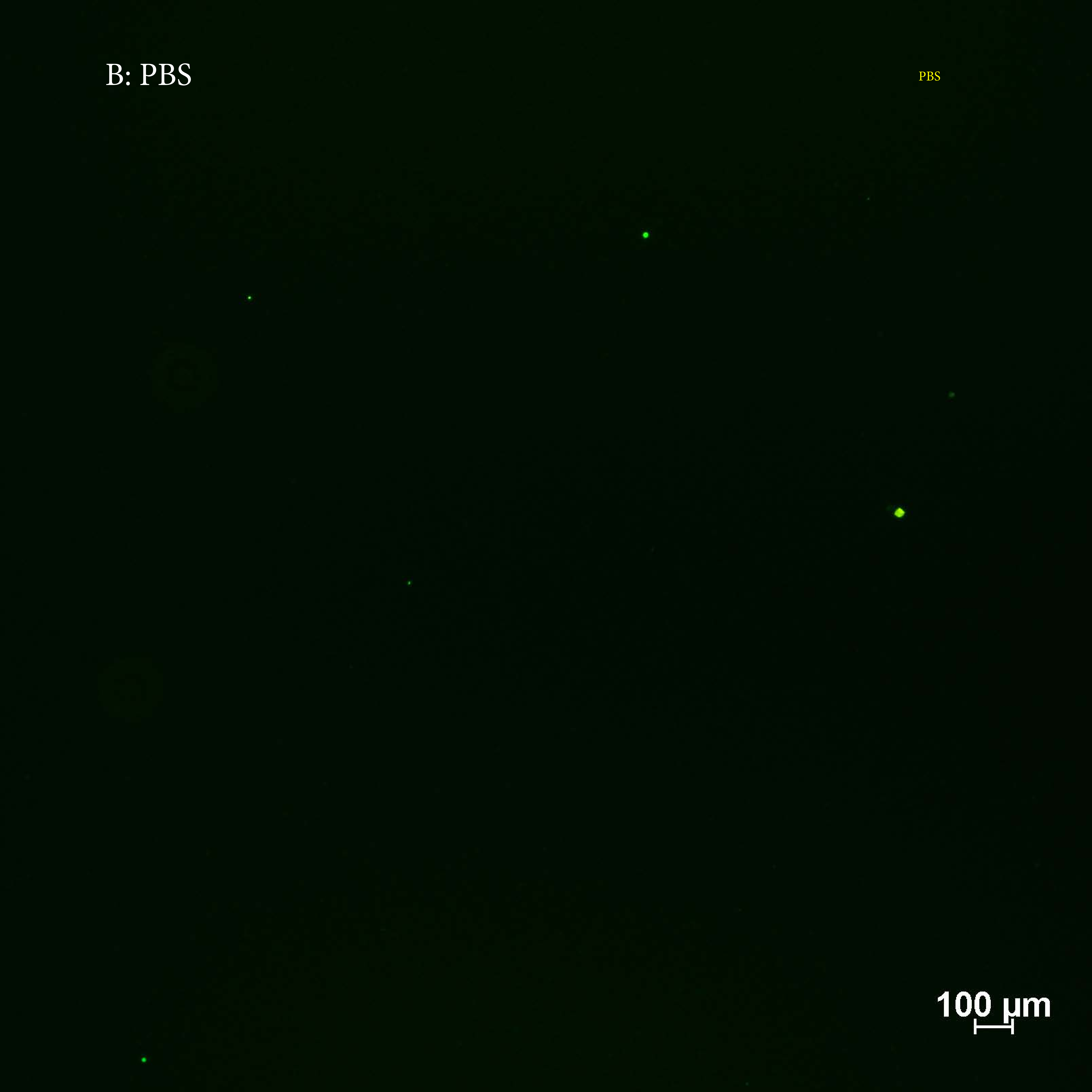
B: PBS
PBS
100 µm

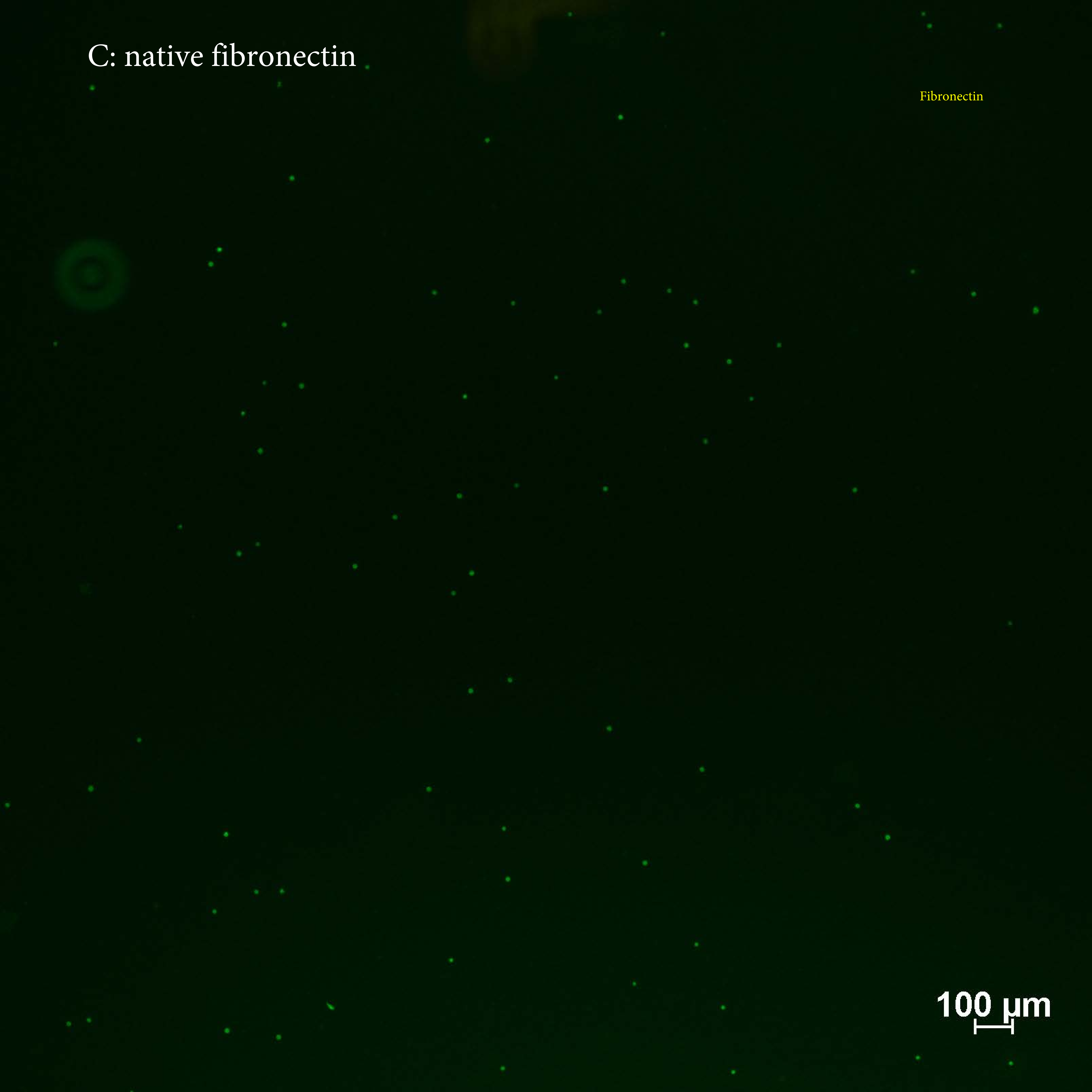
C: native fibronectin
Fibronectin
100 µm

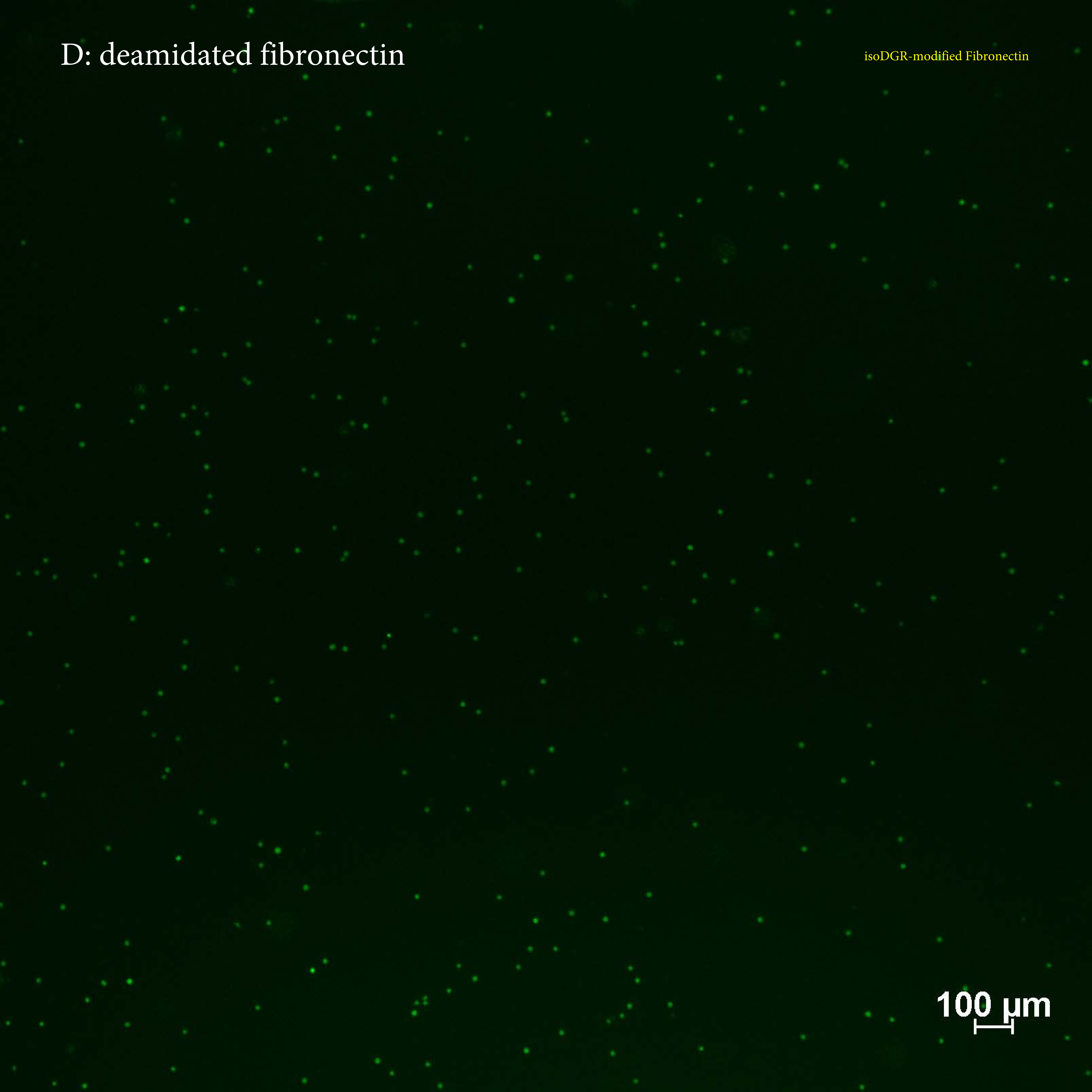
D: deamidated fibronectin
isoDGR-modified Fibronectin
100 µm

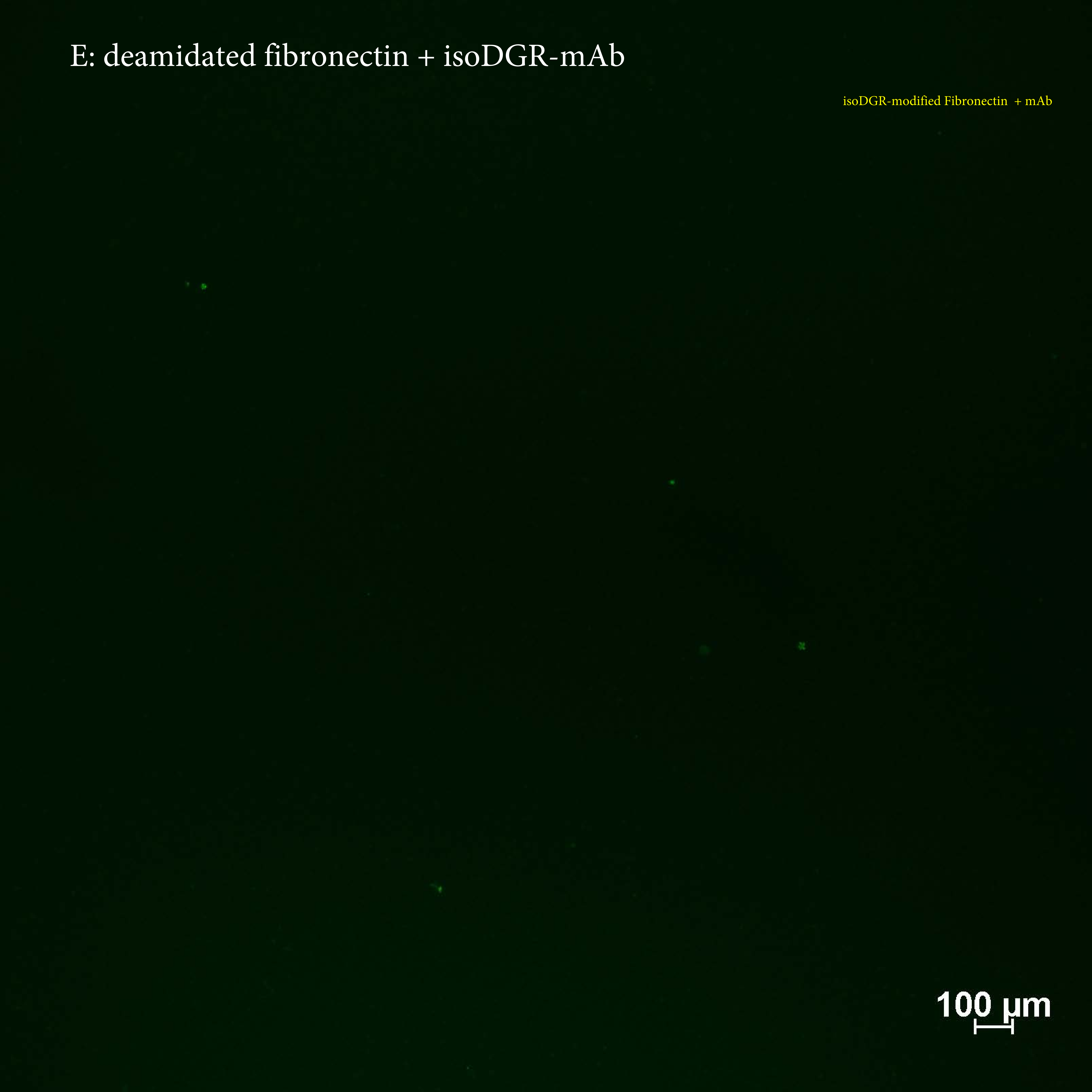
E: deamidated fibronectin + isoDGR-mAb
isoDGR-modified Fibronectin + mAb
100 µm

### Supplementary figure 5: Full images of Western blot

#### RAW cell Lysates

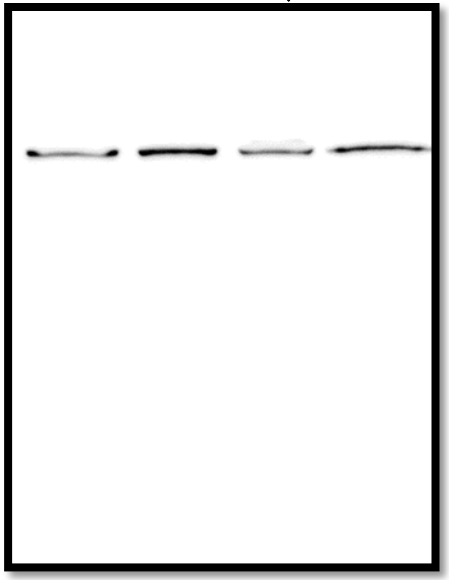


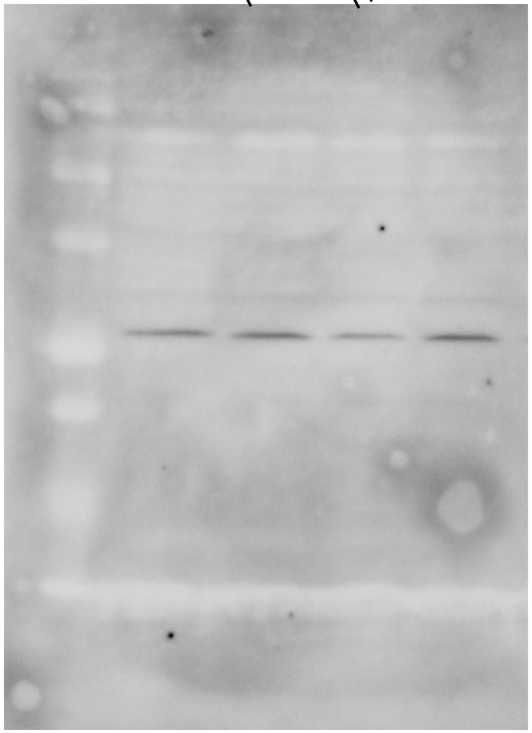

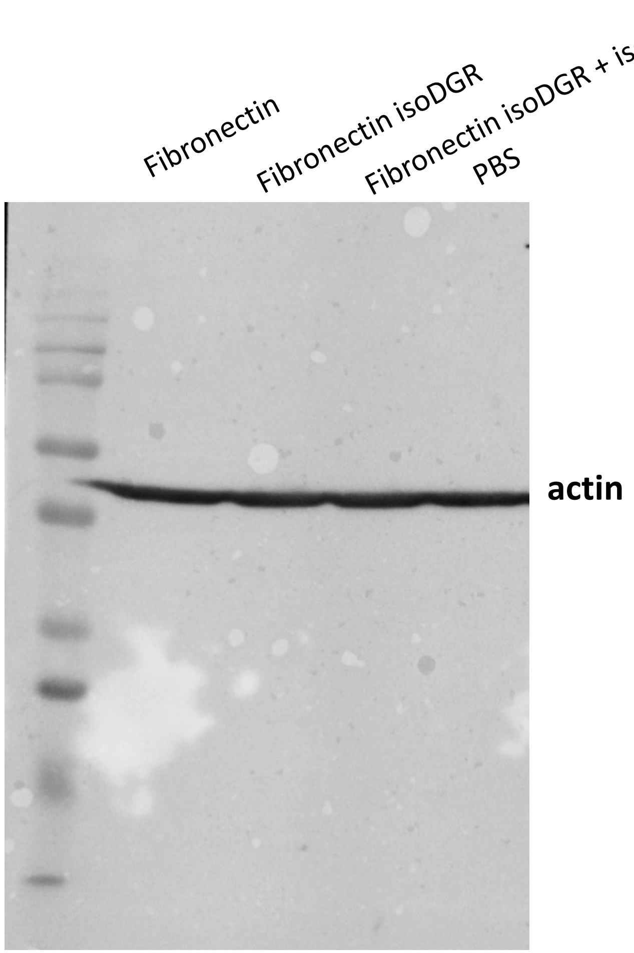
Fibronectin
Fibronectin isoDGR
Fibronectin isoDGR + isoDGR
mab
PBS
actin

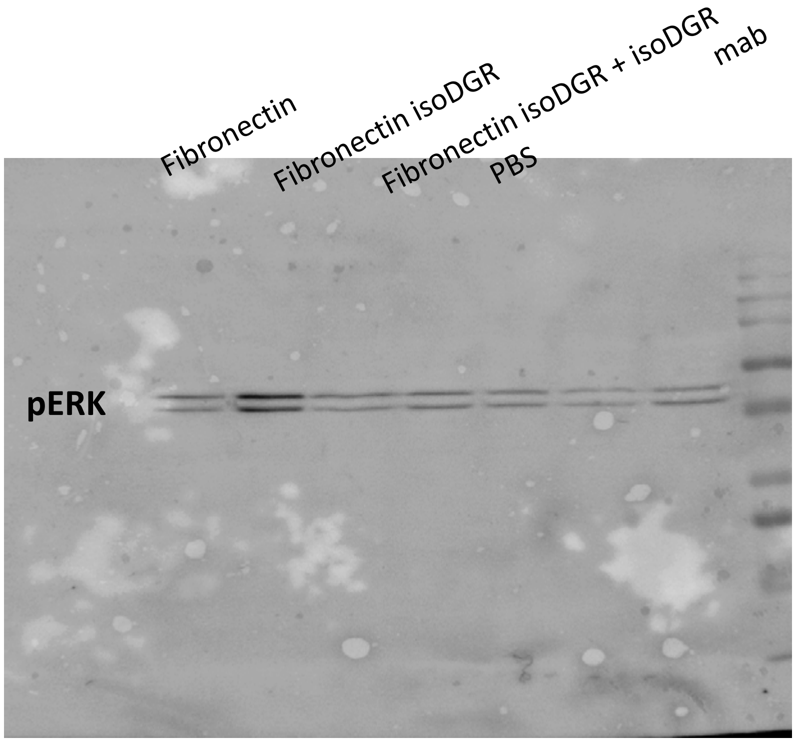
Fibronectin
Fibronectin isoDGR
Fibronectin isoDGR + isoDGR
mab
PBS
pERK

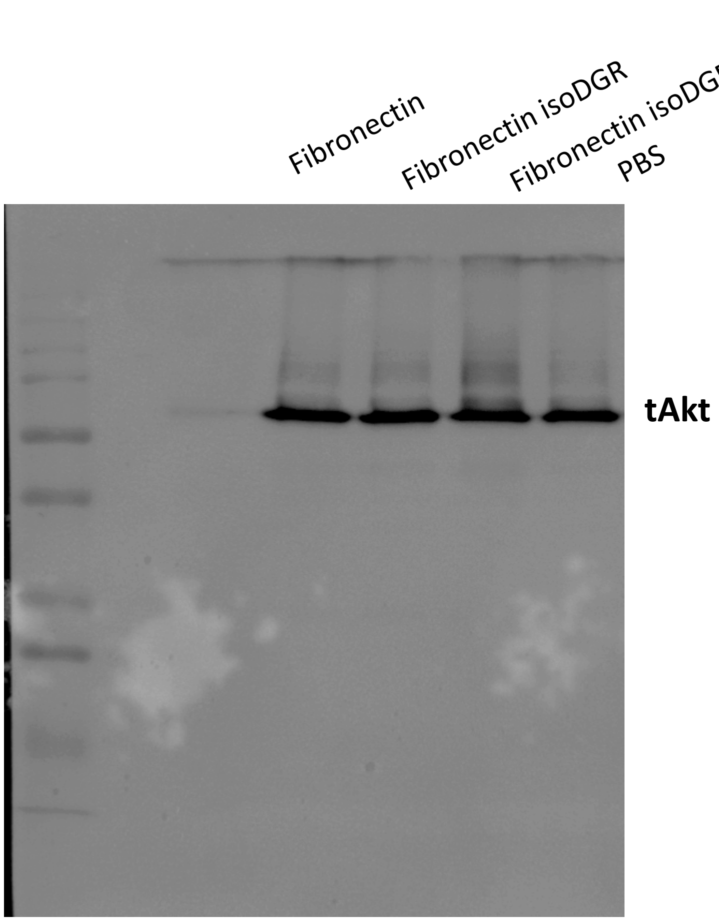
Fibronectin
Fibronectin isoDGR
Fibronectin isoDGR + isoDGR mab
PBS
tAkt

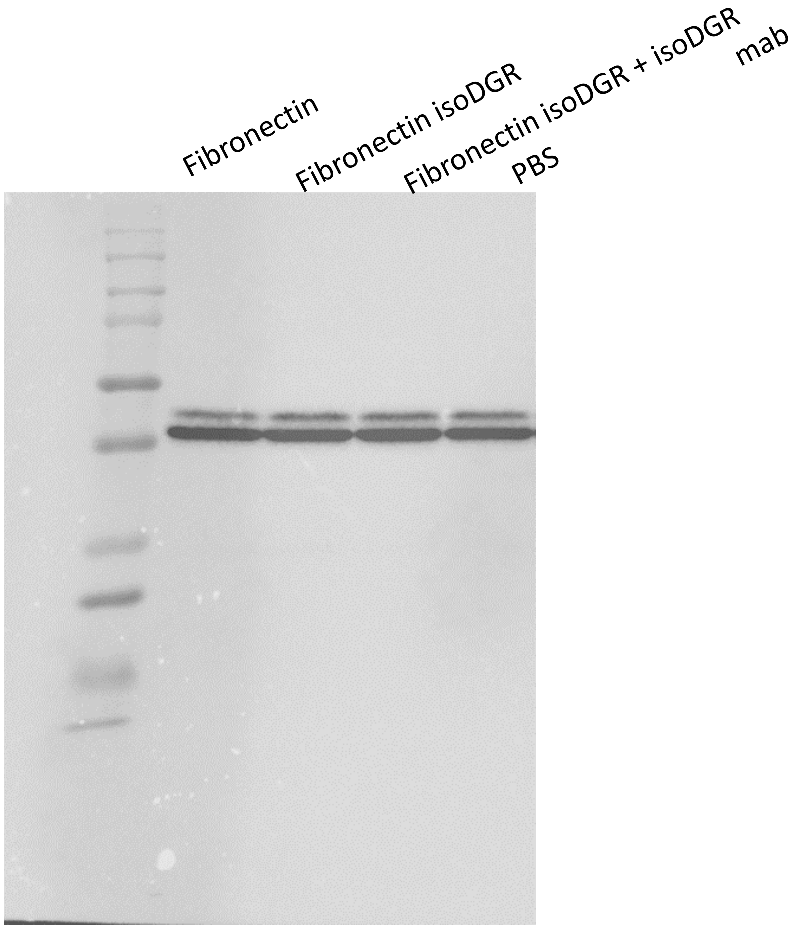
Fibronectin
Fibronectin isoDGR
Fibronectin isoDGR + isoDGR mab
PBS

### U937 monocytes cell lysates

**PBS naFN deFN**

**- + - + - +**

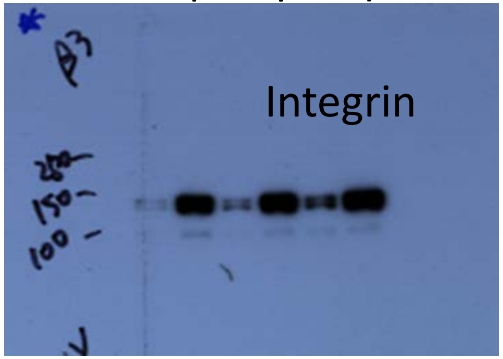


**PBS naFN deFN**

**- + - + - +**

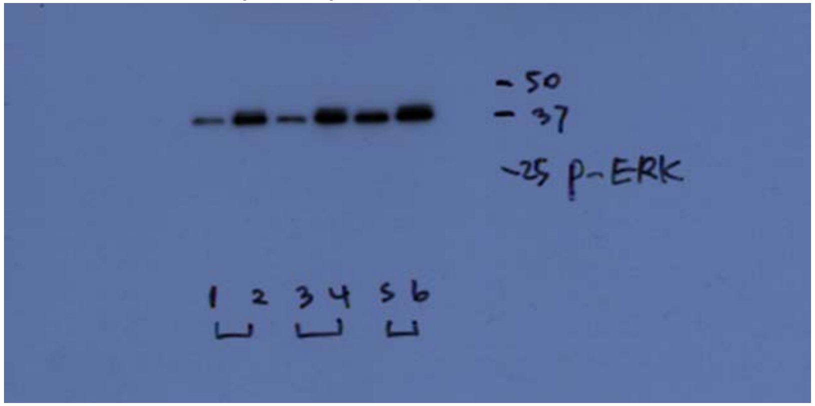


**PBS naFN deFN**

**- + - + - +**

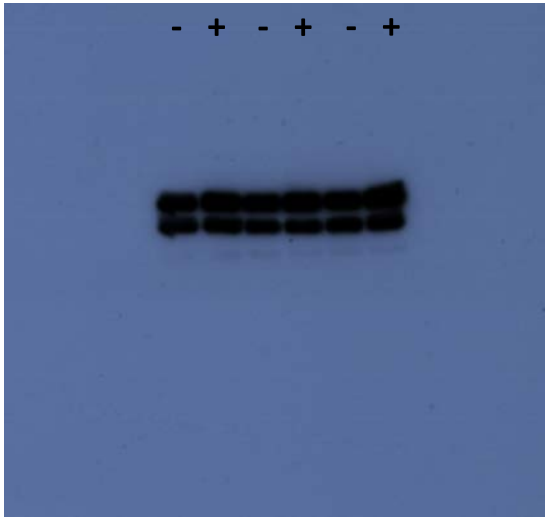

**PBS naFN deFN**

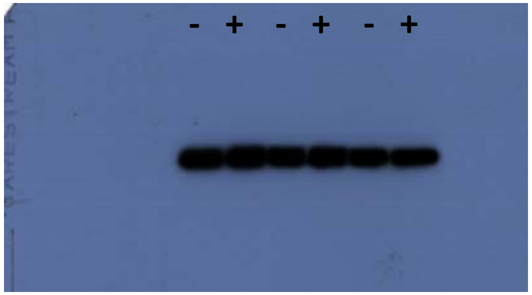


**PBS naFN deFN**

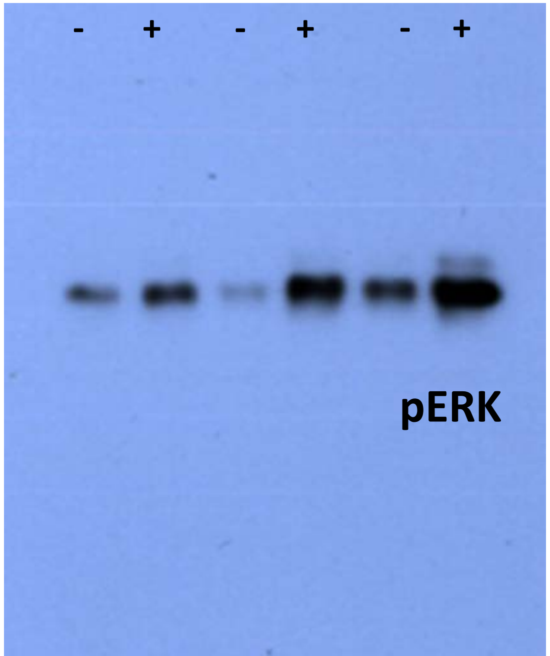
