## Supplementary Data 3 for "Aging-induced isoDGR-modified fibronectin activates monocytic and endothelial cells to promote atherosclerosis"

### MATRIX SCIENCE Mascot Search Results

#### Peptide View

MS/MS Fragmentation of **DNRGNLLQCICTGNGRGEWK**
Found in **sp|P02751|FINC_HUMAN** in **uni_human_i**, sp|P02751|FINC_HUMAN Fibronectin OS=Homo sapiens GN=FN1 PE=1 SV=4

Match to Query 4674: 2347.101162 from(783.374330,3+) intensity(24035.0781) rtinseconds(1391) scans(3711) index(3049)
Title: Fibronectin_deamidation_Spectrum010857_scans__3711_RTINSECONDS=1391
Data file G:__NewmanPaper_Park2\\isoDGR\\FN_C18\\mgf\\T\\TFibronectin_deamidation.mgf

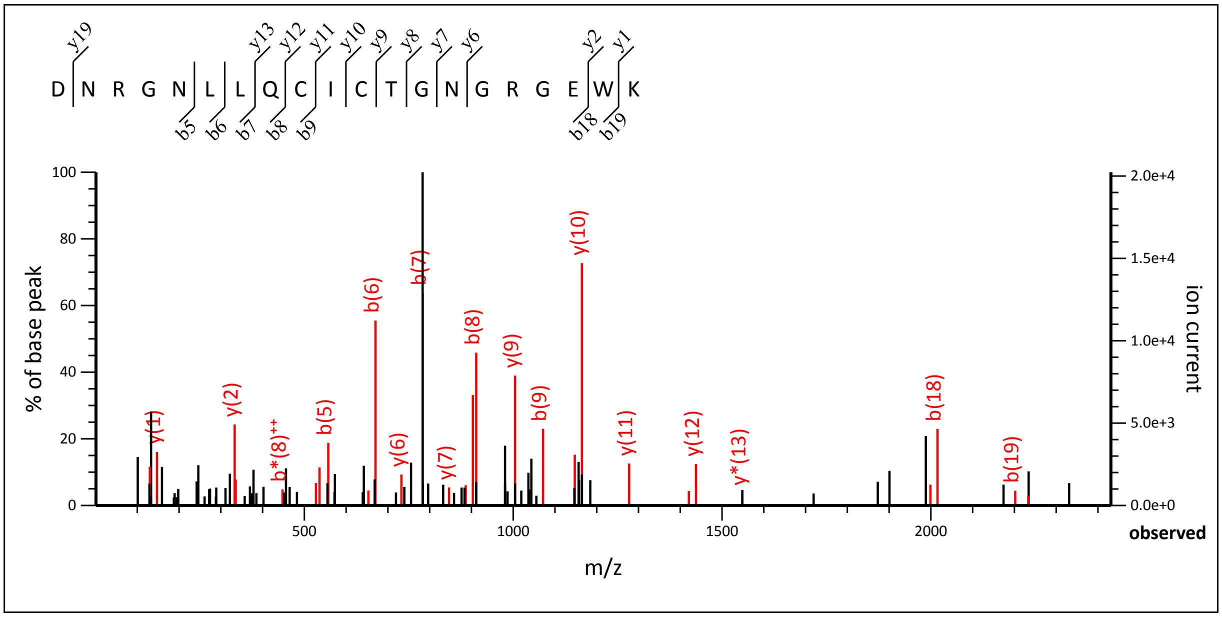


1.07 to 2431.06

Label all possible matches ○ Label matches used for scoring ◉

**Monoisotopic mass of neutral peptide Mr(calc):** 2347.0910
**Fixed modifications:** Carbamidomethyl (C) (apply to specified residues or termini only)
**Ions Score:** 50 **Expect:** 0.0015
**Matches :** 28/222 fragment ions using 60 most intense peaks (help)

| # | b | b++ | b* | b*++ | b0 | b0++ | Seq. | y | y++ | y* | y*++ | y0 | y0++ | # |
|---|---|---|---|---|---|---|---|---|---|---|---|---|---|---|
| 1 | 116.0342 | 58.5207 | | | 98.0237 | 49.5155 | D | | | | | | | 20 |
| 2 | 230.0771 | 115.5422 | 213.0506 | 107.0289 | 212.0666 | 106.5369 | N | **2233.0713** | 1117.0393 | 2216.0448 | 1108.5260 | 2215.0608 | 1108.0340 | 19 |
| 3 | 386.1783 | 193.5928 | 369.1517 | 185.0795 | 368.1677 | 184.5875 | R | 2119.0284 | 1060.0178 | 2102.0018 | 1051.5046 | 2101.0178 | 1051.0125 | 18 |
| 4 | 443.1997 | 222.1035 | 426.1732 | 213.5902 | 425.1892 | 213.0982 | G | 1962.9273 | 981.9673 | 1945.9007 | 973.4540 | 1944.9167 | 972.9620 | 17 |
| 5 | **557.2426** | 279.1250 | 540.2161 | 270.6117 | 539.2321 | 270.1197 | N | 1905.9058 | 953.4565 | 1888.8793 | 944.9433 | 1887.8952 | 944.4513 | 16 |
| 6 | **670.3267** | **335.6670** | 653.3002 | 327.1537 | 652.3161 | 326.6617 | L | 1791.8629 | 896.4351 | 1774.8363 | 887.9218 | 1773.8523 | 887.4298 | 15 |
| 7 | **783.4108** | 392.2090 | 766.3842 | 383.6958 | 765.4002 | 383.2037 | L | 1678.7788 | 839.8930 | 1661.7523 | 831.3798 | 1660.7683 | 830.8878 | 14 |
| 8 | **911.4694** | 456.2383 | 894.4428 | 447.7250 | 893.4588 | 447.2330 | Q | 1565.6948 | 783.3510 | 1548.6682 | 774.8377 | 1547.6842 | 774.3457 | 13 |
| 9 | **1071.5000** | **536.2536** | 1054.4735 | 527.7404 | 1053.4894 | 527.2484 | C | **1437.6362** | 719.3217 | 1420.6096 | 710.8085 | 1419.6256 | 710.3164 | 12 |
| 10 | 1184.5841 | 592.7957 | 1167.5575 | 584.2824 | 1166.5735 | 583.7904 | I | **1277.6055** | 639.3064 | 1260.5790 | 630.7931 | 1259.5950 | 630.3011 | 11 |
| 11 | 1344.6147 | 672.8110 | 1327.5882 | 664.2977 | 1326.6042 | 663.8057 | C | **1164.5215** | 582.7644 | 1147.4949 | 574.2511 | 1146.5109 | 573.7591 | 10 |
| 12 | 1445.6624 | 723.3348 | 1428.6358 | 714.8216 | 1427.6518 | 714.3296 | T | **1004.4908** | 502.7490 | 987.4643 | 494.2358 | 986.4803 | 493.7438 | 9 |
| 13 | 1502.6839 | 751.8456 | 1485.6573 | 743.3323 | 1484.6733 | 742.8403 | G | **903.4431** | 452.2252 | 886.4166 | 443.7119 | 885.4326 | 443.2199 | 8 |
| 14 | 1616.7268 | 808.8670 | 1599.7002 | 800.3538 | 1598.7162 | 799.8617 | N | **846.4217** | 423.7145 | 829.3951 | 415.2012 | 828.4111 | 414.7092 | 7 |
| 15 | 1673.7482 | 837.3778 | 1656.7217 | 828.8645 | 1655.7377 | 828.3725 | G | **732.3787** | 366.6930 | 715.3522 | 358.1797 | 714.3682 | 357.6877 | 6 |
| 16 | 1829.8494 | 915.4283 | 1812.8228 | 906.9150 | 1811.8388 | 906.4230 | R | 675.3573 | 338.1823 | 658.3307 | 329.6690 | 657.3467 | 329.1770 | 5 |
| 17 | 1886.8708 | 943.9391 | 1869.8443 | 935.4258 | 1868.8603 | 934.9338 | G | 519.2562 | 260.1317 | 502.2296 | 251.6185 | 501.2456 | 251.1264 | 4 |
| 18 | **2015.9134** | 1008.4603 | 1998.8869 | 999.9471 | 1997.9029 | 999.4551 | E | 462.2347 | 231.6210 | 445.2082 | 223.1077 | 444.2241 | 222.6157 | 3 |

| | | | | | | | | | | | | | | |
|---|---|---|---|---|---|---|---|---|---|---|---|---|---|---|
| **19** | *2201.9927* | 1101.5000 | 2184.9662 | 1092.9867 | 2183.9822 | 1092.4947 | **W** | *333.1921* | 167.0997 | 316.1656 | 158.5864 | | | **2** |
| **20** | | | | | | | **K** | *147.1128* | 74.0600 | 130.0863 | 65.5468 | | | **1** |

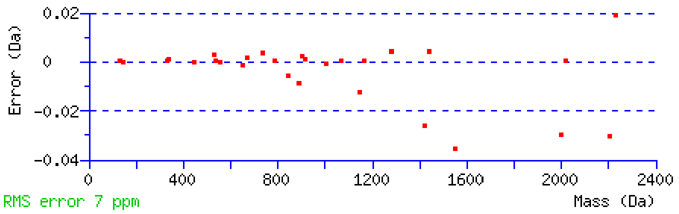


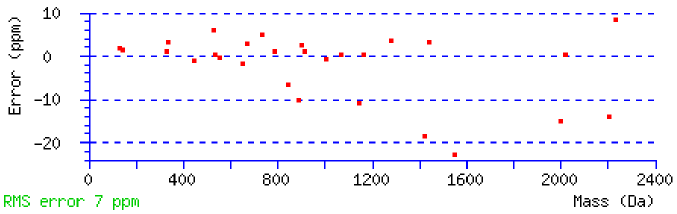


NCBI **BLAST** search of DNRGNLLQCICTGNGRGEWK
(Parameters: blastp, nr protein database, expect=20000, no filter, PAM30)
Other BLAST web gateways

**All matches to this query**

| Score | Mr(calc) | Delta | Sequence |
|---|---|---|---|
| 50.1 | 2347.0910 | 0.0102 | DNRGNLLQCICTGNGRGEWK |

**Mascot:** http://www.matrixscience.com/

### MATRIX SCIENCE Mascot Search Results

#### Peptide View

MS/MS Fragmentation of **DNRGNLLQCICTGNGRGEWK**
Found in **sp|P02751|FINC_HUMAN** in **uni_human_i**, sp|P02751|FINC_HUMAN Fibronectin OS=Homo sapiens GN=FN1 PE=1 SV=4

Match to Query 4677: 2348.077842 from(783.699890,3+) intensity(13478.5449) rtinseconds(1530) scans(4068) index(3371)
Title: Fibronectin_deamidation_Spectrum011179_scans__4068_RTINSECONDS=1530
Data file G:__NewmanPaper_Park2\\isoDGR\\FN_C18\\mgf\\T\\TFibronectin_deamidation.mgf

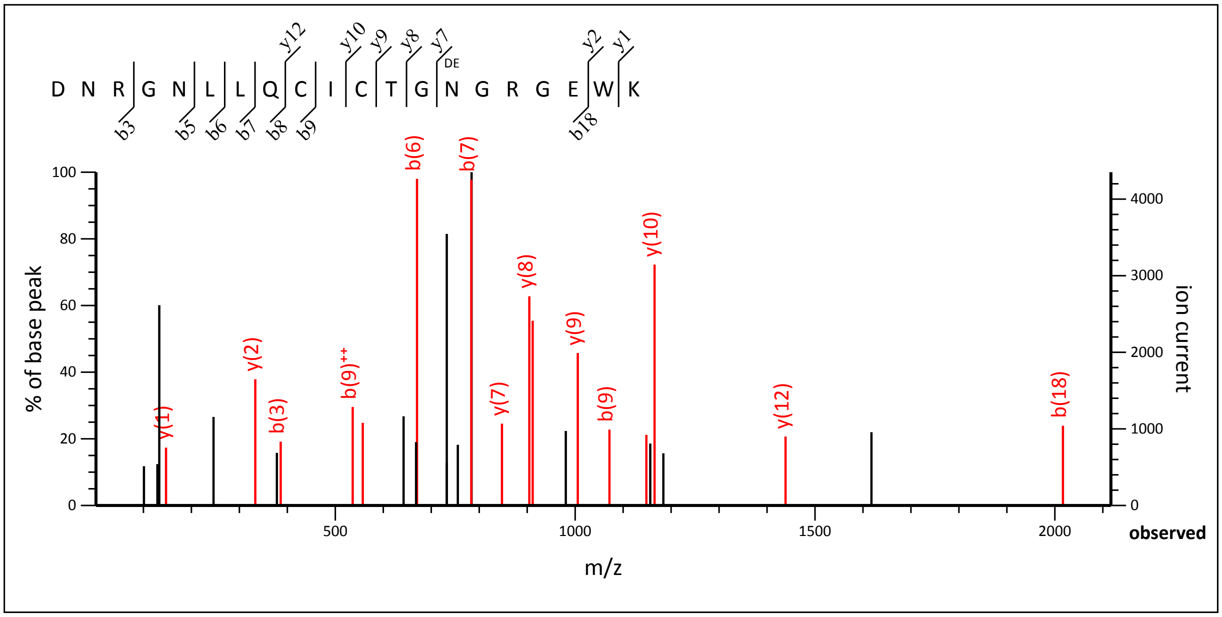


1.07 to 2116.86

Label all possible matches ○ Label matches used for scoring ◉

**Monoisotopic mass of neutral peptide Mr(calc):** 2348.0750
**Fixed modifications:** Carbamidomethyl (C) (apply to specified residues or termini only)
**Variable modifications:**
**N14** : Deamidated (NQ)
**Ions Score:** 69 **Expect:** 2.6e-005
**Matches :** 16/222 fragment ions using 21 most intense peaks (help)

| # | b | b++ | b* | b*++ | b0 | b0++ | Seq. | y | y++ | y* | y*++ | y0 | y0++ | # |
|---|---|---|---|---|---|---|---|---|---|---|---|---|---|---|
| 1 | 116.0342 | 58.5207 | | | 98.0237 | 49.5155 | D | | | | | | | 20 |
| 2 | 230.0771 | 115.5422 | 213.0506 | 107.0289 | 212.0666 | 106.5369 | N | 2234.0553 | 1117.5313 | 2217.0288 | 1109.0180 | 2216.0448 | 1108.5260 | 19 |
| 3 | ***386.1783*** | 193.5928 | 369.1517 | 185.0795 | 368.1677 | 184.5875 | R | 2120.0124 | 1060.5098 | 2102.9859 | 1051.9966 | 2102.0018 | 1051.5046 | 18 |
| 4 | 443.1997 | 222.1035 | 426.1732 | 213.5902 | 425.1892 | 213.0982 | G | 1963.9113 | 982.4593 | 1946.8847 | 973.9460 | 1945.9007 | 973.4540 | 17 |
| 5 | ***557.2426*** | 279.1250 | 540.2161 | 270.6117 | 539.2321 | 270.1197 | N | 1906.8898 | 953.9486 | 1889.8633 | 945.4353 | 1888.8793 | 944.9433 | 16 |
| 6 | ***670.3267*** | 335.6670 | 653.3002 | 327.1537 | 652.3161 | 326.6617 | L | 1792.8469 | 896.9271 | 1775.8204 | 888.4138 | 1774.8363 | 887.9218 | 15 |
| 7 | ***783.4108*** | 392.2090 | 766.3842 | 383.6958 | 765.4002 | 383.2037 | L | 1679.7628 | 840.3851 | 1662.7363 | 831.8718 | 1661.7523 | 831.3798 | 14 |
| 8 | ***911.4694*** | 456.2383 | 894.4428 | 447.7250 | 893.4588 | 447.2330 | Q | 1566.6788 | 783.8430 | 1549.6522 | 775.3298 | 1548.6682 | 774.8377 | 13 |
| 9 | ***1071.5000*** | **536.2536** | 1054.4735 | 527.7404 | 1053.4894 | 527.2484 | C | ***1438.6202*** | 719.8137 | 1421.5936 | 711.3005 | 1420.6096 | 710.8085 | 12 |
| 10 | 1184.5841 | 592.7957 | 1167.5575 | 584.2824 | 1166.5735 | 583.7904 | I | 1278.5895 | 639.7984 | 1261.5630 | 631.2851 | 1260.5790 | 630.7931 | 11 |
| 11 | 1344.6147 | 672.8110 | 1327.5882 | 664.2977 | 1326.6042 | 663.8057 | C | ***1165.5055*** | 583.2564 | 1148.4789 | 574.7431 | 1147.4949 | 574.2511 | 10 |
| 12 | 1445.6624 | 723.3348 | 1428.6358 | 714.8216 | 1427.6518 | 714.3296 | T | ***1005.4748*** | 503.2411 | 988.4483 | 494.7278 | 987.4643 | 494.2358 | 9 |
| 13 | 1502.6839 | 751.8456 | 1485.6573 | 743.3323 | 1484.6733 | 742.8403 | G | ***904.4272*** | 452.7172 | 887.4006 | 444.2039 | 886.4166 | 443.7119 | 8 |
| 14 | 1617.7108 | 809.3590 | 1600.6843 | 800.8458 | 1599.7002 | 800.3538 | N | ***847.4057*** | 424.2065 | 830.3791 | 415.6932 | 829.3951 | 415.2012 | 7 |
| 15 | 1674.7323 | 837.8698 | 1657.7057 | 829.3565 | 1656.7217 | 828.8645 | G | 732.3787 | 366.6930 | 715.3522 | 358.1797 | 714.3682 | 357.6877 | 6 |
| 16 | 1830.8334 | 915.9203 | 1813.8068 | 907.4071 | 1812.8228 | 906.9150 | R | 675.3573 | 338.1823 | 658.3307 | 329.6690 | 657.3467 | 329.1770 | 5 |
| 17 | 1887.8548 | 944.4311 | 1870.8283 | 935.9178 | 1869.8443 | 935.4258 | G | 519.2562 | 260.1317 | 502.2296 | 251.6185 | 501.2456 | 251.1264 | 4 |

| | | | | | | | | | | | | | | |
|---|---|---|---|---|---|---|---|---|---|---|---|---|---|---|
| **18** | ***2016.8974*** | 1008.9524 | 1999.8709 | 1000.4391 | 1998.8869 | 999.9471 | **E** | 462.2347 | 231.6210 | 445.2082 | 223.1077 | 444.2241 | 222.6157 | **3** |
| **19** | 2202.9767 | 1101.9920 | 2185.9502 | 1093.4787 | 2184.9662 | 1092.9867 | **W** | ***333.1921*** | 167.0997 | 316.1656 | 158.5864 | | | **2** |
| **20** | | | | | | | **K** | ***147.1128*** | 74.0600 | 130.0863 | 65.5468 | | | **1** |

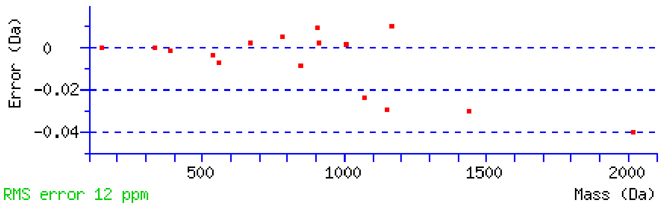


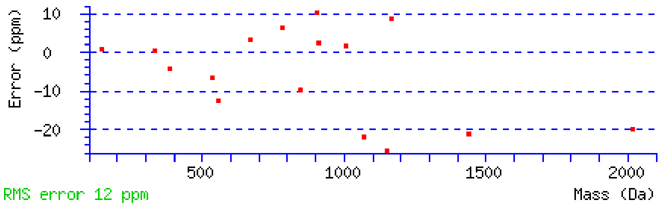


NCBI **BLAST** search of DNRGNLLQCICTGNGRGEWK
(Parameters: blastp, nr protein database, expect=20000, no filter, PAM30)
Other BLAST web gateways

**All matches to this query**

| Score | Mr(calc) | Delta | Sequence | Site Analysis |
|---|---|---|---|---|
| 68.8 | 2348.0750 | 0.0029 | DNRGNLLQCICTGNGRGEWK | Deamidated N14 100.00% |
| 12.7 | 2348.0750 | 0.0029 | DNRGNLLQCICTGNGRGEWK | Deamidated Q8 0.00% |

**Mascot:** http://www.matrixscience.com/

### MATRIX SCIENCE Mascot Search Results

#### Peptide View

MS/MS Fragmentation of **GNLLQCICTGNGRGEWK**
Found in **sp|P02751|FINC_HUMAN** in **uni_human_i**, sp|P02751|FINC_HUMAN Fibronectin OS=Homo sapiens GN=FN1 PE=1 SV=4

Match to Query 3654: 1961.918292 from(654.980040,3+) intensity(27002.9297) rtinseconds(1469) scans(3907) index(3226)
Title: Fibronectin_deamidation_Spectrum011034_scans__3907_RTINSECONDS=1469
Data file G:__NewmanPaper_Park2\\isoDGR\\FN_C18\\mgf\\T\\TFibronectin_deamidation.mgf

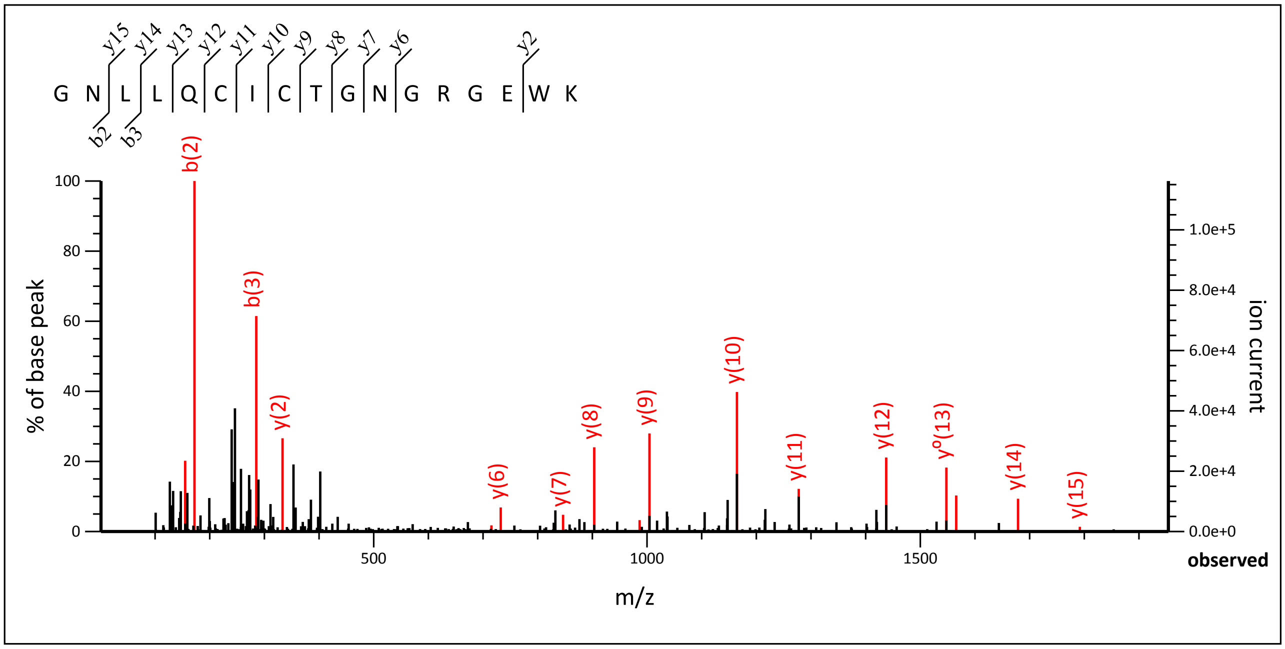


1.07 to 1953.84

Label all possible matches ○ Label matches used for scoring ◉

**Monoisotopic mass of neutral peptide Mr(calc):** 1961.9200
**Fixed modifications:** Carbamidomethyl (C) (apply to specified residues or termini only)
**Ions Score:** 74 **Expect:** 6.5e-006
**Matches :** 17/170 fragment ions using 34 most intense peaks (help)

| # | b | $b^{++}$ | b* | $b^{*++}$ | $b^0$ | $b^{0++}$ | Seq. | y | $y^{++}$ | y* | $y^{*++}$ | $y^0$ | $y^{0++}$ | # |
|---|---|---|---|---|---|---|---|---|---|---|---|---|---|---|
| 1 | 58.0287 | 29.5180 | | | | | G | | | | | | | 17 |
| 2 | **172.0717** | 86.5395 | 155.0451 | 78.0262 | | | N | 1905.9058 | 953.4565 | 1888.8793 | 944.9433 | 1887.8952 | 944.4513 | 16 |
| 3 | **285.1557** | 143.0815 | 268.1292 | 134.5682 | | | L | ***1791.8629*** | 896.4351 | 1774.8363 | 887.9218 | 1773.8523 | 887.4298 | 15 |
| 4 | 398.2398 | 199.6235 | 381.2132 | 191.1103 | | | L | ***1678.7788*** | 839.8930 | 1661.7523 | 831.3798 | 1660.7683 | 830.8878 | 14 |
| 5 | 526.2984 | 263.6528 | 509.2718 | 255.1396 | | | Q | ***1565.6948*** | 783.3510 | 1548.6682 | 774.8377 | 1547.6842 | 774.3457 | 13 |
| 6 | 686.3290 | 343.6681 | 669.3025 | 335.1549 | | | C | ***1437.6362*** | 719.3217 | 1420.6096 | 710.8085 | 1419.6256 | 710.3164 | 12 |
| 7 | 799.4131 | 400.2102 | 782.3865 | 391.6969 | | | I | ***1277.6055*** | 639.3064 | 1260.5790 | 630.7931 | 1259.5950 | 630.3011 | 11 |
| 8 | 959.4437 | 480.2255 | 942.4172 | 471.7122 | | | C | ***1164.5215*** | 582.7644 | 1147.4949 | 574.2511 | 1146.5109 | 573.7591 | 10 |
| 9 | 1060.4914 | 530.7493 | 1043.4649 | 522.2361 | 1042.4808 | 521.7441 | T | ***1004.4908*** | 502.7490 | 987.4643 | 494.2358 | 986.4803 | 493.7438 | 9 |
| 10 | 1117.5129 | 559.2601 | 1100.4863 | 550.7468 | 1099.5023 | 550.2548 | G | ***903.4431*** | 452.2252 | 886.4166 | 443.7119 | 885.4326 | 443.2199 | 8 |
| 11 | 1231.5558 | 616.2815 | 1214.5293 | 607.7683 | 1213.5452 | 607.2763 | N | ***846.4217*** | 423.7145 | 829.3951 | 415.2012 | 828.4111 | 414.7092 | 7 |
| 12 | 1288.5773 | 644.7923 | 1271.5507 | 636.2790 | 1270.5667 | 635.7870 | G | ***732.3787*** | 366.6930 | 715.3522 | 358.1797 | 714.3682 | 357.6877 | 6 |
| 13 | 1444.6784 | 722.8428 | 1427.6518 | 714.3296 | 1426.6678 | 713.8375 | R | 675.3573 | 338.1823 | 658.3307 | 329.6690 | 657.3467 | 329.1770 | 5 |
| 14 | 1501.6998 | 751.3536 | 1484.6733 | 742.8403 | 1483.6893 | 742.3483 | G | 519.2562 | 260.1317 | 502.2296 | 251.6185 | 501.2456 | 251.1264 | 4 |
| 15 | 1630.7424 | 815.8749 | 1613.7159 | 807.3616 | 1612.7319 | 806.8696 | E | 462.2347 | 231.6210 | 445.2082 | 223.1077 | 444.2241 | 222.6157 | 3 |

| | | | | | | | | | | | | | | |
|---|---|---|---|---|---|---|---|---|---|---|---|---|---|---|
| **16** | 1816.8217 | 908.9145 | 1799.7952 | 900.4012 | 1798.8112 | 899.9092 | **W** | ***333.1921*** | 167.0997 | 316.1656 | 158.5864 | | | **2** |
| **17** | | | | | | | **K** | 147.1128 | 74.0600 | 130.0863 | 65.5468 | | | **1** |

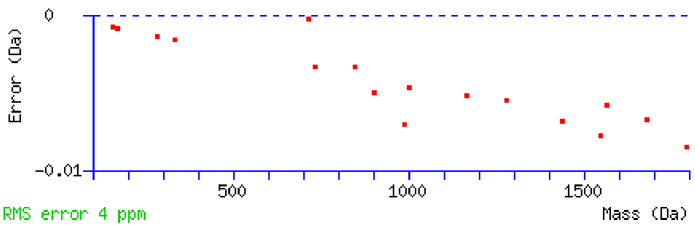


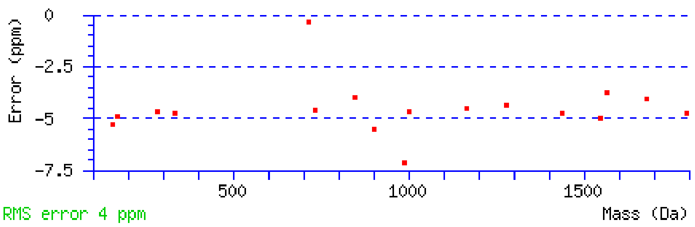


NCBI **BLAST** search of GNLLQCICTGNGRGEWK
(Parameters: blastp, nr protein database, expect=20000, no filter, PAM30)
Other BLAST web gateways

**All matches to this query**

| Score | Mr(calc) | Delta | Sequence |
|---|---|---|---|
| 74.3 | 1961.9200 | -0.0017 | GNLLQCICTGNGRGEWK |

**Mascot:** http://www.matrixscience.com/

### Mascot Search Results

#### Peptide View

MS/MS Fragmentation of **GNLLQCICTGNGRGEWK**
Found in **sp|P02751|FINC_HUMAN** in **uni_human_i**, sp|P02751|FINC_HUMAN Fibronectin OS=Homo sapiens GN=FN1 PE=1 SV=4

Match to Query 3660: 1962.904872 from(655.308900,3+) intensity(279416.8750) rtinseconds(1631) scans(4346) index(3622)
Title: Fibronectin_deamidation_Spectrum011430_scans__4346_RTINSECONDS=1631
Data file G:__NewmanPaper_Park2\\isoDGR\\FN_C18\\mgf\\T\\TFibronectin_deamidation.mgf

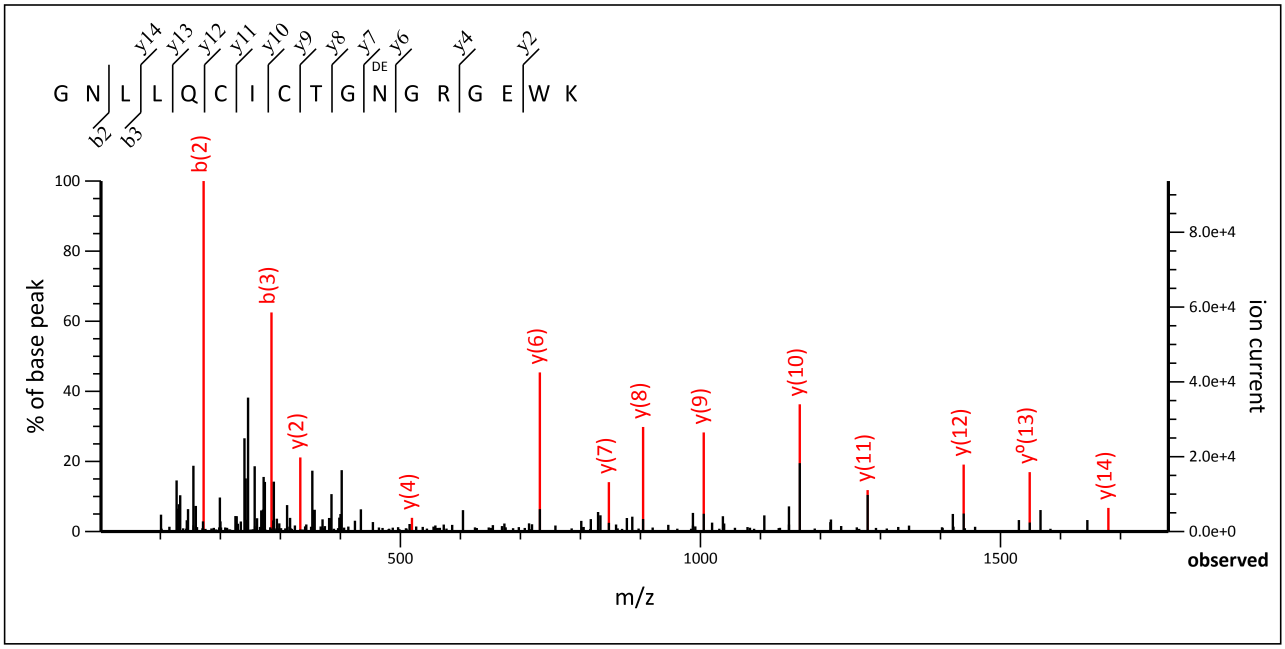


1.07 to 1779.76

Label all possible matches ○ Label matches used for scoring ◉

**Monoisotopic mass of neutral peptide Mr(calc):** 1962.9040
**Fixed modifications:** Carbamidomethyl (C) (apply to specified residues or termini only)
**Variable modifications:**
**N11** : Deamidated (NQ)
**Ions Score:** 88 **Expect:** 5.9e-007
**Matches :** 13/170 fragment ions using 16 most intense peaks (help)

| # | b | $b^{++}$ | b* | $b^{*++}$ | $b^{0}$ | $b^{0++}$ | Seq. | y | $y^{++}$ | y* | $y^{*++}$ | $y^{0}$ | $y^{0++}$ | # |
|---|---|---|---|---|---|---|---|---|---|---|---|---|---|---|
| 1 | 58.0287 | 29.5180 | | | | | G | | | | | | | 17 |
| 2 | **172.0717** | 86.5395 | 155.0451 | 78.0262 | | | N | 1906.8898 | 953.9486 | 1889.8633 | 945.4353 | 1888.8793 | 944.9433 | 16 |
| 3 | **285.1557** | 143.0815 | 268.1292 | 134.5682 | | | L | 1792.8469 | 896.9271 | 1775.8204 | 888.4138 | 1774.8363 | 887.9218 | 15 |
| 4 | 398.2398 | 199.6235 | 381.2132 | 191.1103 | | | L | ***1679.7628*** | 840.3851 | 1662.7363 | 831.8718 | 1661.7523 | 831.3798 | 14 |
| 5 | 526.2984 | 263.6528 | 509.2718 | 255.1396 | | | Q | 1566.6788 | 783.8430 | 1549.6522 | 775.3298 | 1548.6682 | 774.8377 | 13 |
| 6 | 686.3290 | 343.6681 | 669.3025 | 335.1549 | | | C | ***1438.6202*** | 719.8137 | 1421.5936 | 711.3005 | 1420.6096 | 710.8085 | 12 |
| 7 | 799.4131 | 400.2102 | 782.3865 | 391.6969 | | | I | ***1278.5895*** | 639.7984 | 1261.5630 | 631.2851 | 1260.5790 | 630.7931 | 11 |
| 8 | 959.4437 | 480.2255 | 942.4172 | 471.7122 | | | C | ***1165.5055*** | 583.2564 | 1148.4789 | 574.7431 | 1147.4949 | 574.2511 | 10 |
| 9 | 1060.4914 | 530.7493 | 1043.4649 | 522.2361 | 1042.4808 | 521.7441 | T | ***1005.4748*** | 503.2411 | 988.4483 | 494.7278 | 987.4643 | 494.2358 | 9 |
| 10 | 1117.5129 | 559.2601 | 1100.4863 | 550.7468 | 1099.5023 | 550.2548 | G | ***904.4272*** | 452.7172 | 887.4006 | 444.2039 | 886.4166 | 443.7119 | 8 |
| 11 | 1232.5398 | 616.7735 | 1215.5133 | 608.2603 | 1214.5293 | 607.7683 | N | ***847.4057*** | 424.2065 | 830.3791 | 415.6932 | 829.3951 | 415.2012 | 7 |
| 12 | 1289.5613 | 645.2843 | 1272.5347 | 636.7710 | 1271.5507 | 636.2790 | G | ***732.3787*** | 366.6930 | 715.3522 | 358.1797 | 714.3682 | 357.6877 | 6 |
| 13 | 1445.6624 | 723.3348 | 1428.6358 | 714.8216 | 1427.6518 | 714.3296 | R | 675.3573 | 338.1823 | 658.3307 | 329.6690 | 657.3467 | 329.1770 | 5 |
| 14 | 1502.6839 | 751.8456 | 1485.6573 | 743.3323 | 1484.6733 | 742.8403 | G | ***519.2562*** | 260.1317 | 502.2296 | 251.6185 | 501.2456 | 251.1264 | 4 |

| | | | | | | | | | | | | | | |
|---|---|---|---|---|---|---|---|---|---|---|---|---|---|---|
| **15** | 1631.7265 | 816.3669 | 1614.6999 | 807.8536 | 1613.7159 | 807.3616 | **E** | 462.2347 | 231.6210 | 445.2082 | 223.1077 | 444.2241 | 222.6157 | **3** |
| **16** | 1817.8058 | 909.4065 | 1800.7792 | 900.8932 | 1799.7952 | 900.4012 | **W** | *333.1921* | 167.0997 | 316.1656 | 158.5864 | | | **2** |
| **17** | | | | | | | **K** | 147.1128 | 74.0600 | 130.0863 | 65.5468 | | | **1** |

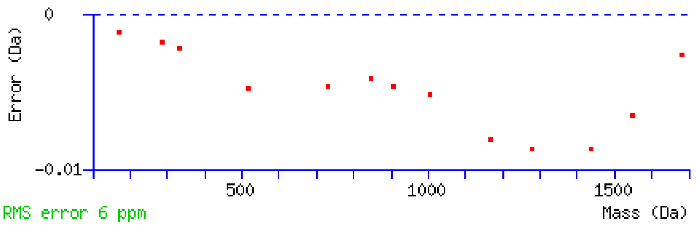


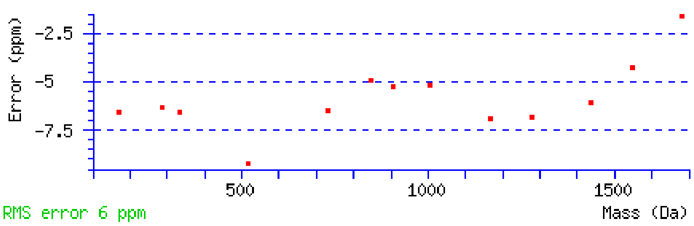


NCBI **BLAST** search of GNLLQCICTGNGRGEWK
(Parameters: blastp, nr protein database, expect=20000, no filter, PAM30)
Other BLAST web gateways

**All matches to this query**

| Score | Mr(calc) | Delta | Sequence | Site Analysis |
|---|---|---|---|---|
| 87.8 | 1962.9040 | 0.0009 | GNLLQCICTGNGRGEWK | Deamidated N11 100.00% |
| 12.4 | 1962.9040 | 0.0009 | GNLLQCICTGNGRGEWK | Deamidated Q5 0.00% |

**Mascot:** http://www.matrixscience.com/

### MATRIX SCIENCE Mascot Search Results

#### Peptide View

MS/MS Fragmentation of **GNLLQCICTGNGRGEWKCER**
Found in **sp|P02751|FINC_HUMAN** in **uni_human_i**, sp|P02751|FINC_HUMAN Fibronectin OS=Homo sapiens GN=FN1 PE=1 SV=4

Match to Query 4809: 2407.097682 from(803.373170,3+) intensity(16363.6309) rtinseconds(1268) scans(3394) index(2763)
Title: Fibronectin_deamidation_Spectrum010571_scans__3394_RTINSECONDS=1268
Data file G:__NewmanPaper_Park2\\isoDGR\\FN_C18\\mgf\\T\\TFibronectin_deamidation.mgf

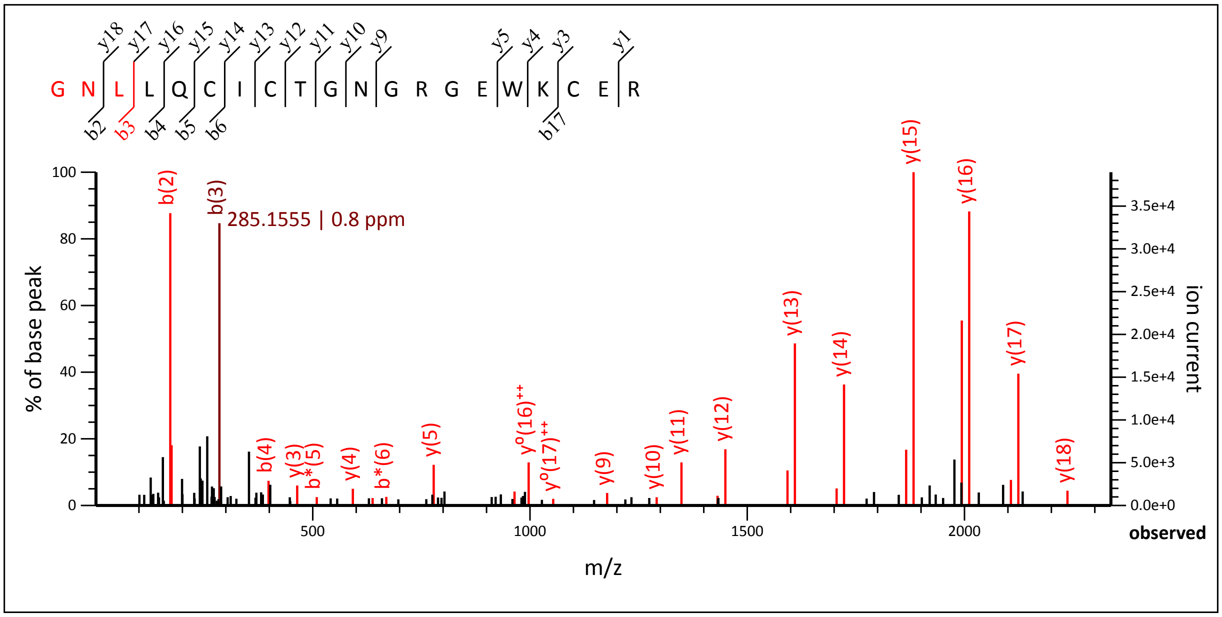


1.07 to 2337.04

Label all possible matches ○ Label matches used for scoring ◉

**Monoisotopic mass of neutral peptide Mr(calc):** 2407.0943
**Fixed modifications:** Carbamidomethyl (C) (apply to specified residues or termini only)
**Ions Score:** 127 **Expect:** 2.7e-011
**Matches :** 30/208 fragment ions using 39 most intense peaks (help)

| # | b | $b^{++}$ | b* | $b^{*++}$ | $b^0$ | $b^{0++}$ | Seq. | y | $y^{++}$ | y* | $y^{*++}$ | $y^0$ | $y^{0++}$ | # |
|---|---|---|---|---|---|---|---|---|---|---|---|---|---|---|
| 1 | 58.0287 | 29.5180 | | | | | G | | | | | | | 20 |
| 2 | 172.0717 | 86.5395 | 155.0451 | 78.0262 | | | N | 2351.0802 | 1176.0437 | 2334.0536 | 1167.5304 | 2333.0696 | 1167.0384 | 19 |
| 3 | 285.1557 | 143.0815 | 268.1292 | 134.5682 | | | L | 2237.0372 | 1119.0223 | 2220.0107 | 1110.5090 | 2219.0267 | 1110.0170 | 18 |
| 4 | 398.2398 | 199.6235 | 381.2132 | 191.1103 | | | L | 2123.9532 | 1062.4802 | 2106.9266 | 1053.9670 | 2105.9426 | 1053.4749 | 17 |
| 5 | 526.2984 | 263.6528 | 509.2718 | 255.1396 | | | Q | 2010.8691 | 1005.9382 | 1993.8426 | 997.4249 | 1992.8585 | 996.9329 | 16 |
| 6 | 686.3290 | 343.6681 | 669.3025 | 335.1549 | | | C | 1882.8105 | 941.9089 | 1865.7840 | 933.3956 | 1864.8000 | 932.9036 | 15 |
| 7 | 799.4131 | 400.2102 | 782.3865 | 391.6969 | | | I | 1722.7799 | 861.8936 | 1705.7533 | 853.3803 | 1704.7693 | 852.8883 | 14 |
| 8 | 959.4437 | 480.2255 | 942.4172 | 471.7122 | | | C | 1609.6958 | 805.3515 | 1592.6693 | 796.8383 | 1591.6853 | 796.3463 | 13 |
| 9 | 1060.4914 | 530.7493 | 1043.4649 | 522.2361 | 1042.4808 | 521.7441 | T | 1449.6652 | 725.3362 | 1432.6386 | 716.8229 | 1431.6546 | 716.3309 | 12 |
| 10 | 1117.5129 | 559.2601 | 1100.4863 | 550.7468 | 1099.5023 | 550.2548 | G | 1348.6175 | 674.8124 | 1331.5909 | 666.2991 | 1330.6069 | 665.8071 | 11 |
| 11 | 1231.5558 | 616.2815 | 1214.5293 | 607.7683 | 1213.5452 | 607.2763 | N | 1291.5960 | 646.3017 | 1274.5695 | 637.7884 | 1273.5855 | 637.2964 | 10 |
| 12 | 1288.5773 | 644.7923 | 1271.5507 | 636.2790 | 1270.5667 | 635.7870 | G | 1177.5531 | 589.2802 | 1160.5266 | 580.7669 | 1159.5425 | 580.2749 | 9 |
| 13 | 1444.6784 | 722.8428 | 1427.6518 | 714.3296 | 1426.6678 | 713.8375 | R | 1120.5316 | 560.7695 | 1103.5051 | 552.2562 | 1102.5211 | 551.7642 | 8 |
| 14 | 1501.6998 | 751.3536 | 1484.6733 | 742.8403 | 1483.6893 | 742.3483 | G | 964.4305 | 482.7189 | 947.4040 | 474.2056 | 946.4200 | 473.7136 | 7 |
| 15 | 1630.7424 | 815.8749 | 1613.7159 | 807.3616 | 1612.7319 | 806.8696 | E | 907.4091 | 454.2082 | 890.3825 | 445.6949 | 889.3985 | 445.2029 | 6 |
| 16 | 1816.8217 | 908.9145 | 1799.7952 | 900.4012 | 1798.8112 | 899.9092 | W | 778.3665 | 389.6869 | 761.3399 | 381.1736 | 760.3559 | 380.6816 | 5 |
| 17 | 1944.9167 | 972.9620 | 1927.8902 | 964.4487 | 1926.9061 | 963.9567 | K | 592.2872 | 296.6472 | 575.2606 | 288.1339 | 574.2766 | 287.6419 | 4 |
| 18 | 2104.9474 | 1052.9773 | 2087.9208 | 1044.4640 | 2086.9368 | 1043.9720 | C | 464.1922 | 232.5997 | 447.1656 | 224.0865 | 446.1816 | 223.5945 | 3 |

| | | | | | | | | | | | | | |
|---|---|---|---|---|---|---|---|---|---|---|---|---|---|
| 19 | 2233.9900 | 1117.4986 | 2216.9634 | 1108.9853 | 2215.9794 | 1108.4933 | E | 304.1615 | 152.5844 | 287.1350 | 144.0711 | 286.1510 | 143.5791 | 2 |
| 20 | | | | | | | R | *175.1190* | 88.0631 | 158.0924 | 79.5498 | | | 1 |

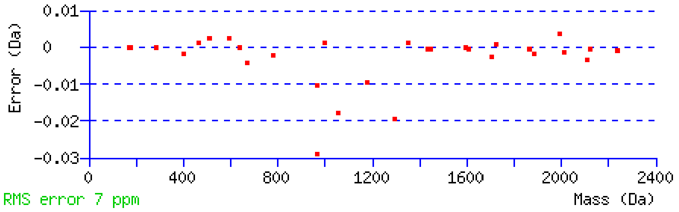


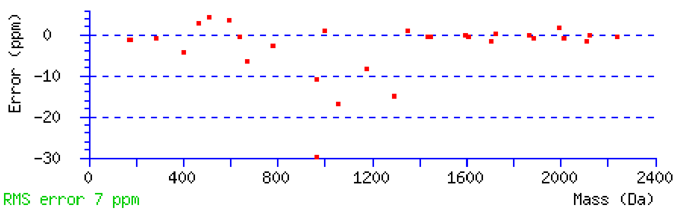


NCBI **BLAST** search of GNLLQCICTGNGRGEWKCER
(Parameters: blastp, nr protein database, expect=20000, no filter, PAM30)
Other BLAST web gateways

**All matches to this query**

| Score | Mr(calc) | Delta | Sequence |
|---|---|---|---|
| 127.4 | 2407.0943 | 0.0033 | GNLLQCICTGNGRGEWKCER |
| 3.5 | 2406.1024 | 0.9952 | MQLSTFFSFMLENYTHIHK |
| 3.0 | 2406.1024 | 0.9952 | MQLSTFFSFMLENYTHIHK |

**Mascot:** http://www.matrixscience.com/

### MATRIX SCIENCE Mascot Search Results

#### Peptide View

MS/MS Fragmentation of **GNLLQCICTGNGRGEWKCER**
Found in **sp|P02751|FINC_HUMAN** in **uni_human_i**, sp|P02751|FINC_HUMAN Fibronectin OS=Homo sapiens GN=FN1 PE=1 SV=4

Match to Query 4810: 2408.078772 from(803.700200,3+) intensity(27722.1465) rtinseconds(1418) scans(3784) index(3114)
Title: Fibronectin_deamidation_Spectrum010922_scans__3784_RTINSECONDS=1418
Data file G:__NewmanPaper_Park2\\isoDGR\\FN_C18\\mgf\\T\\TFibronectin_deamidation.mgf

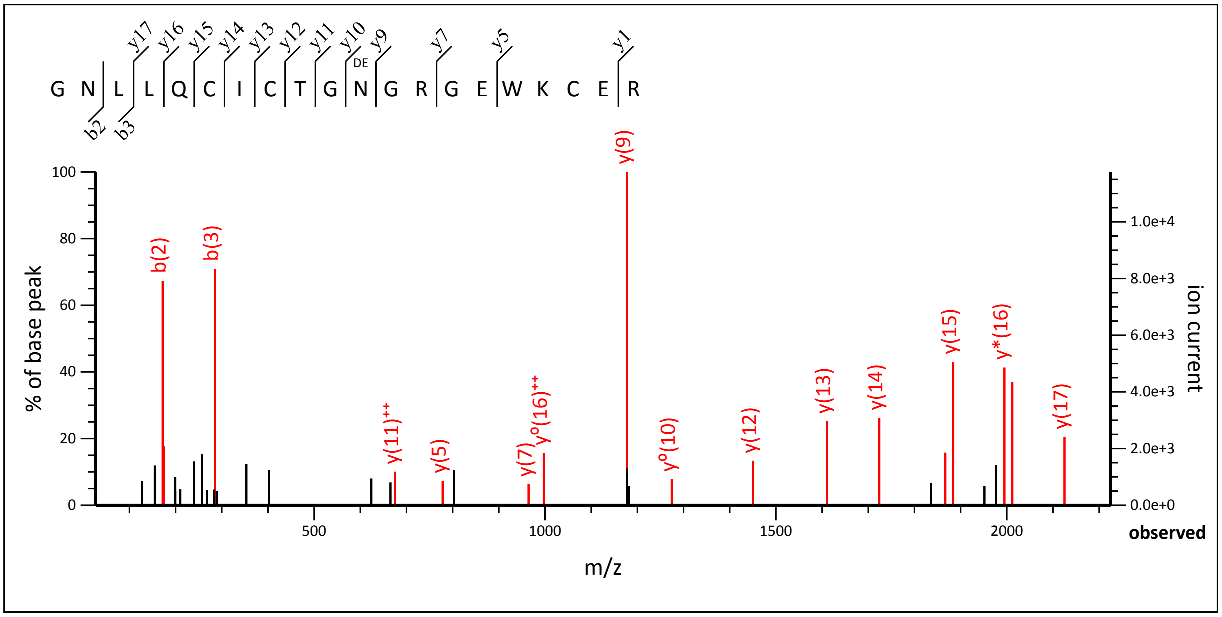


27.05 to 2224.95

Label all possible matches ○ Label matches used for scoring ●

**Monoisotopic mass of neutral peptide Mr(calc):** 2408.0784
**Fixed modifications:** Carbamidomethyl (C) (apply to specified residues or termini only)
**Variable modifications:**
**N11** : Deamidated (NQ)
**Ions Score:** 77 **Expect:** 2.3e-006
**Matches :** 17/208 fragment ions using 24 most intense peaks (help)

| # | b | $b^{++}$ | b* | $b^{*++}$ | $b^{0}$ | $b^{0++}$ | Seq. | y | $y^{++}$ | y* | $y^{*++}$ | $y^{0}$ | $y^{0++}$ | # |
|---|---|---|---|---|---|---|---|---|---|---|---|---|---|---|
| 1 | 58.0287 | 29.5180 | | | | | G | | | | | | | 20 |
| 2 | **172.0717** | 86.5395 | 155.0451 | 78.0262 | | | N | 2352.0642 | 1176.5357 | 2335.0376 | 1168.0225 | 2334.0536 | 1167.5304 | 19 |
| 3 | **285.1557** | 143.0815 | 268.1292 | 134.5682 | | | L | 2238.0213 | 1119.5143 | 2220.9947 | 1111.0010 | 2220.0107 | 1110.5090 | 18 |
| 4 | 398.2398 | 199.6235 | 381.2132 | 191.1103 | | | L | ***2124.9372*** | 1062.9722 | 2107.9106 | 1054.4590 | 2106.9266 | 1053.9670 | 17 |
| 5 | 526.2984 | 263.6528 | 509.2718 | 255.1396 | | | Q | ***2011.8531*** | 1006.4302 | 1994.8266 | 997.9169 | 1993.8426 | 997.4249 | 16 |
| 6 | 686.3290 | 343.6681 | 669.3025 | 335.1549 | | | C | ***1883.7946*** | 942.4009 | 1866.7680 | 933.8876 | 1865.7840 | 933.3956 | 15 |
| 7 | 799.4131 | 400.2102 | 782.3865 | 391.6969 | | | I | ***1723.7639*** | 862.3856 | 1706.7374 | 853.8723 | 1705.7533 | 853.3803 | 14 |
| 8 | 959.4437 | 480.2255 | 942.4172 | 471.7122 | | | C | ***1610.6798*** | 805.8436 | 1593.6533 | 797.3303 | 1592.6693 | 796.8383 | 13 |
| 9 | 1060.4914 | 530.7493 | 1043.4649 | 522.2361 | 1042.4808 | 521.7441 | T | ***1450.6492*** | 725.8282 | 1433.6226 | 717.3150 | 1432.6386 | 716.8229 | 12 |
| 10 | 1117.5129 | 559.2601 | 1100.4863 | 550.7468 | 1099.5023 | 550.2548 | G | 1349.6015 | **675.3044** | 1332.5750 | 666.7911 | 1331.5909 | 666.2991 | 11 |
| 11 | 1232.5398 | 616.7735 | 1215.5133 | 608.2603 | 1214.5293 | 607.7683 | N | 1292.5800 | 646.7937 | 1275.5535 | 638.2804 | 1274.5695 | 637.7884 | 10 |
| 12 | 1289.5613 | 645.2843 | 1272.5347 | 636.7710 | 1271.5507 | 636.2790 | G | ***1177.5531*** | 589.2802 | 1160.5266 | 580.7669 | 1159.5425 | 580.2749 | 9 |
| 13 | 1445.6624 | 723.3348 | 1428.6358 | 714.8216 | 1427.6518 | 714.3296 | R | 1120.5316 | 560.7695 | 1103.5051 | 552.2562 | 1102.5211 | 551.7642 | 8 |
| 14 | 1502.6839 | 751.8456 | 1485.6573 | 743.3323 | 1484.6733 | 742.8403 | G | ***964.4305*** | 482.7189 | 947.4040 | 474.2056 | 946.4200 | 473.7136 | 7 |
| 15 | 1631.7265 | 816.3669 | 1614.6999 | 807.8536 | 1613.7159 | 807.3616 | E | 907.4091 | 454.2082 | 890.3825 | 445.6949 | 889.3985 | 445.2029 | 6 |
| 16 | 1817.8058 | 909.4065 | 1800.7792 | 900.8932 | 1799.7952 | 900.4012 | W | ***778.3665*** | 389.6869 | 761.3399 | 381.1736 | 760.3559 | 380.6816 | 5 |
| 17 | 1945.9007 | 973.4540 | 1928.8742 | 964.9407 | 1927.8902 | 964.4487 | K | 592.2872 | 296.6472 | 575.2606 | 288.1339 | 574.2766 | 287.6419 | 4 |

| | | | | | | | | | | | | | | |
|---|---|---|---|---|---|---|---|---|---|---|---|---|---|---|
| **18** | 2105.9314 | 1053.4693 | 2088.9048 | 1044.9561 | 2087.9208 | 1044.4640 | **C** | 464.1922 | 232.5997 | 447.1656 | 224.0865 | 446.1816 | 223.5945 | **3** |
| **19** | 2234.9740 | 1117.9906 | 2217.9474 | 1109.4773 | 2216.9634 | 1108.9853 | **E** | 304.1615 | 152.5844 | 287.1350 | 144.0711 | 286.1510 | 143.5791 | **2** |
| **20** | | | | | | | **R** | *175.1190* | 88.0631 | 158.0924 | 79.5498 | | | **1** |

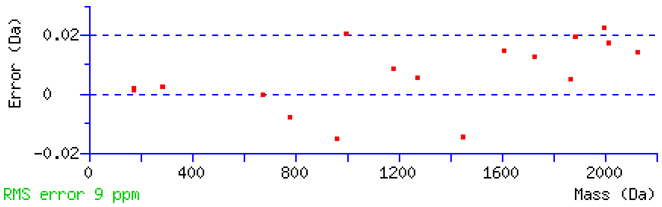


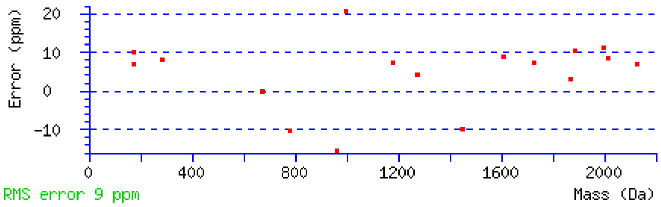


NCBI **BLAST** search of GNLLQCICTGNGRGEWKCER
(Parameters: blastp, nr protein database, expect=20000, no filter, PAM30)
Other BLAST web gateways

**All matches to this query**

| Score | Mr(calc) | Delta | Sequence | Site Analysis |
|---|---|---|---|---|
| 76.6 | 2408.0784 | 0.0004 | GNLLQCICTGNGRGEWKCER | Deamidated N11 100.00% |
| 26.0 | 2408.0784 | 0.0004 | GNLLQCICTGNGRGEWKCER | Deamidated Q5 0.00% |
| 2.8 | 2408.0784 | 0.0004 | GNLLQCICTGNGRGEWKCER | Deamidated N2 0.00% |
| 0.6 | 2407.0864 | 0.9923 | MQLSTFFSFMLENYTHIHK | |

| **Mascot:** http://www.matrixscience.com/ |
|---|

### MATRIX SCIENCE Mascot Search Results

#### Peptide View

MS/MS Fragmentation of **CTCVGNGRGEWTCIAYSQLR**
Found in **sp|P02751|FINC_HUMAN** in **uni_human_i**, sp|P02751|FINC_HUMAN Fibronectin OS=Homo sapiens GN=FN1 PE=1 SV=4

Match to Query 4762: 2387.058612 from(796.693480,3+) intensity(52610.1523) rtinseconds(1520) scans(4038) index(3344)
Title: Fibronectin_deamidation_Spectrum011152_scans__4038_RTINSECONDS=1520
Data file G:__NewmanPaper_Park2\\isoDGR\\FN_C18\\mgf\\T\\TFibronectin_deamidation.mgf

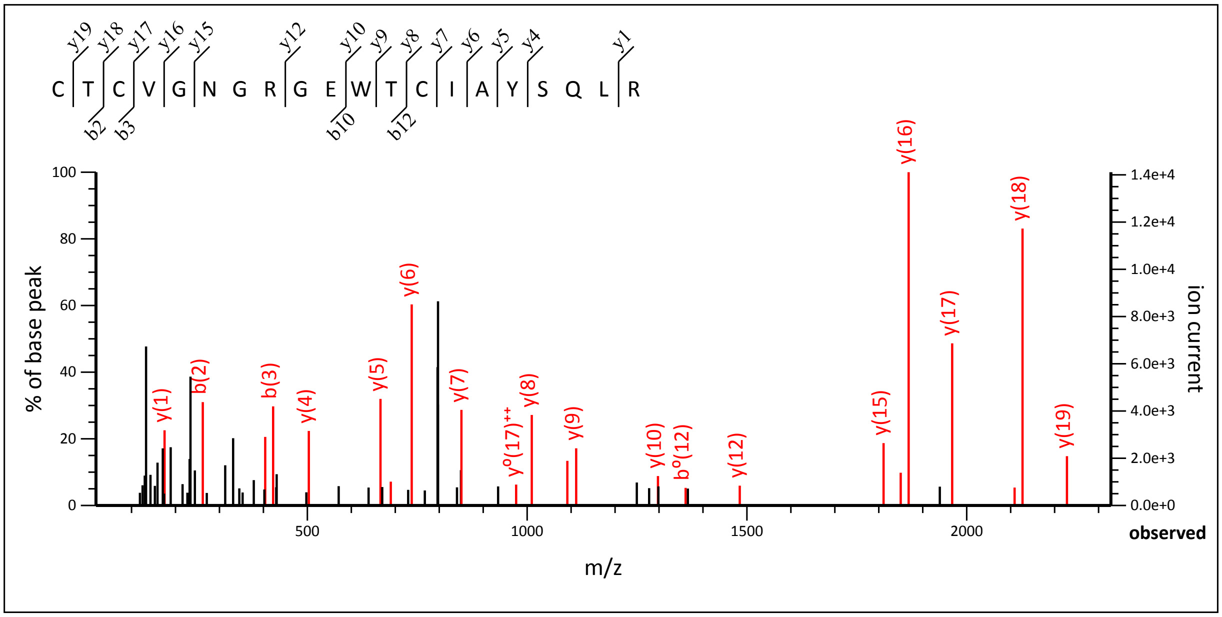


19.08 to 2328.08

Label all possible matches ○ Label matches used for scoring ●

**Monoisotopic mass of neutral peptide Mr(calc):** 2387.0569
**Fixed modifications:** Carbamidomethyl (C) (apply to specified residues or termini only)
**Ions Score:** 115 **Expect:** 4.8e-010
**Matches :** 23/210 fragment ions using 32 most intense peaks (help)

| # | b | $b^{++}$ | b* | $b^{*++}$ | $b^0$ | $b^{0++}$ | Seq. | y | $y^{++}$ | y* | $y^{*++}$ | $y^0$ | $y^{0++}$ | # |
|---|---|---|---|---|---|---|---|---|---|---|---|---|---|---|
| 1 | 161.0379 | 81.0226 | | | | | C | | | | | | | 20 |
| 2 | **262.0856** | 131.5464 | | | 244.0750 | 122.5412 | T | ***2228.0335*** | 1114.5204 | 2211.0070 | 1106.0071 | 2210.0230 | 1105.5151 | 19 |
| 3 | **422.1163** | 211.5618 | | | 404.1057 | 202.5565 | C | ***2126.9859*** | 1063.9966 | 2109.9593 | 1055.4833 | 2108.9753 | 1054.9913 | 18 |
| 4 | 521.1847 | 261.0960 | | | 503.1741 | 252.0907 | V | ***1966.9552*** | 983.9812 | 1949.9287 | 975.4680 | 1948.9446 | 974.9760 | 17 |
| 5 | 578.2061 | 289.6067 | | | 560.1956 | 280.6014 | G | ***1867.8868*** | 934.4470 | 1850.8602 | 925.9338 | 1849.8762 | 925.4418 | 16 |
| 6 | 692.2491 | 346.6282 | 675.2225 | 338.1149 | 674.2385 | 337.6229 | N | ***1810.8653*** | 905.9363 | 1793.8388 | 897.4230 | 1792.8548 | 896.9310 | 15 |
| 7 | 749.2705 | 375.1389 | 732.2440 | 366.6256 | 731.2600 | 366.1336 | G | 1696.8224 | 848.9148 | 1679.7959 | 840.4016 | 1678.8118 | 839.9096 | 14 |
| 8 | 905.3716 | 453.1895 | 888.3451 | 444.6762 | 887.3611 | 444.1842 | R | 1639.8009 | 820.4041 | 1622.7744 | 811.8908 | 1621.7904 | 811.3988 | 13 |
| 9 | 962.3931 | 481.7002 | 945.3665 | 473.1869 | 944.3825 | 472.6949 | G | ***1483.6998*** | 742.3536 | 1466.6733 | 733.8403 | 1465.6893 | 733.3483 | 12 |
| 10 | **1091.4357** | 546.2215 | 1074.4091 | 537.7082 | 1073.4251 | 537.2162 | E | 1426.6784 | 713.8428 | 1409.6518 | 705.3295 | 1408.6678 | 704.8375 | 11 |
| 11 | 1277.5150 | 639.2611 | 1260.4885 | 630.7479 | 1259.5044 | 630.2559 | W | ***1297.6358*** | 649.3215 | 1280.6092 | 640.8082 | 1279.6252 | 640.3162 | 10 |
| 12 | 1378.5627 | **689.7850** | 1361.5361 | 681.2717 | 1360.5521 | 680.7797 | T | ***1111.5565*** | 556.2819 | 1094.5299 | 547.7686 | 1093.5459 | 547.2766 | 9 |
| 13 | 1538.5933 | 769.8003 | 1521.5668 | 761.2870 | 1520.5828 | 760.7950 | C | ***1010.5088*** | 505.7580 | 993.4822 | 497.2448 | 992.4982 | 496.7527 | 8 |
| 14 | 1651.6774 | 826.3423 | 1634.6508 | 817.8291 | 1633.6668 | 817.3371 | I | ***850.4781*** | 425.7427 | 833.4516 | 417.2294 | 832.4676 | 416.7374 | 7 |
| 15 | 1722.7145 | 861.8609 | 1705.6880 | 853.3476 | 1704.7039 | 852.8556 | A | ***737.3941*** | 369.2007 | 720.3675 | 360.6874 | 719.3835 | 360.1954 | 6 |
| 16 | 1885.7778 | 943.3926 | 1868.7513 | 934.8793 | 1867.7673 | 934.3873 | Y | ***666.3570*** | 333.6821 | 649.3304 | 325.1688 | 648.3464 | 324.6768 | 5 |
| 17 | 1972.8099 | 986.9086 | 1955.7833 | 978.3953 | 1954.7993 | 977.9033 | S | ***503.2936*** | 252.1504 | 486.2671 | 243.6372 | 485.2831 | 243.1452 | 4 |
| 18 | 2100.8684 | 1050.9379 | 2083.8419 | 1042.4246 | 2082.8579 | 1041.9326 | Q | 416.2616 | 208.6344 | 399.2350 | 200.1212 | | | 3 |

| | | | | | | | | | | | | | | |
|---|---|---|---|---|---|---|---|---|---|---|---|---|---|---|
| **19** | 2213.9525 | 1107.4799 | 2196.9260 | 1098.9666 | 2195.9419 | 1098.4746 | **L** | 288.2030 | 144.6051 | 271.1765 | 136.0919 | | | **2** |
| **20** | | | | | | | **R** | *175.1190* | 88.0631 | 158.0924 | 79.5498 | | | **1** |

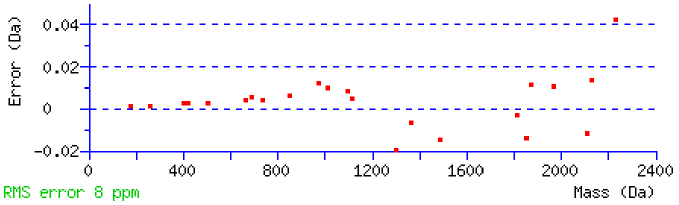


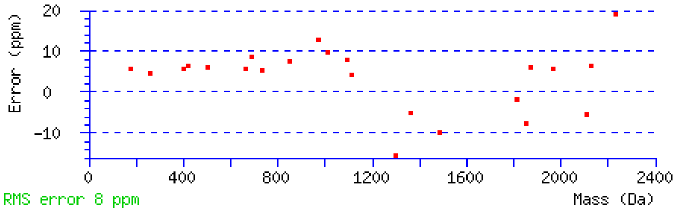


NCBI **BLAST** search of CTCVGNGRGEWTCIAYSQLR
(Parameters: blastp, nr protein database, expect=20000, no filter, PAM30)
Other BLAST web gateways

**All matches to this query**

| Score | Mr(calc) | Delta | Sequence |
|---|---|---|---|
| 114.8 | 2387.0569 | 0.0017 | CTCVGNGRGEWTCIAYSQLR |

**Mascot:** http://www.matrixscience.com/

### MATRIX SCIENCE Mascot Search Results

#### Peptide View

MS/MS Fragmentation of **CTCVGNGRGEWTCIAYSQLR**
Found in **sp|P02751|FINC_HUMAN** in **uni_human_i**, sp|P02751|FINC_HUMAN Fibronectin OS=Homo sapiens GN=FN1 PE=1 SV=4

Match to Query 4770: 2388.041712 from(797.021180,3+) intensity(24799.5410) rtinseconds(1705) scans(4556) index(3811)
Title: Fibronectin_deamidation_Spectrum011619_scans__4556_RTINSECONDS=1705
Data file G:__NewmanPaper_Park2\\isoDGR\\FN_C18\\mgf\\T\\TFibronectin_deamidation.mgf

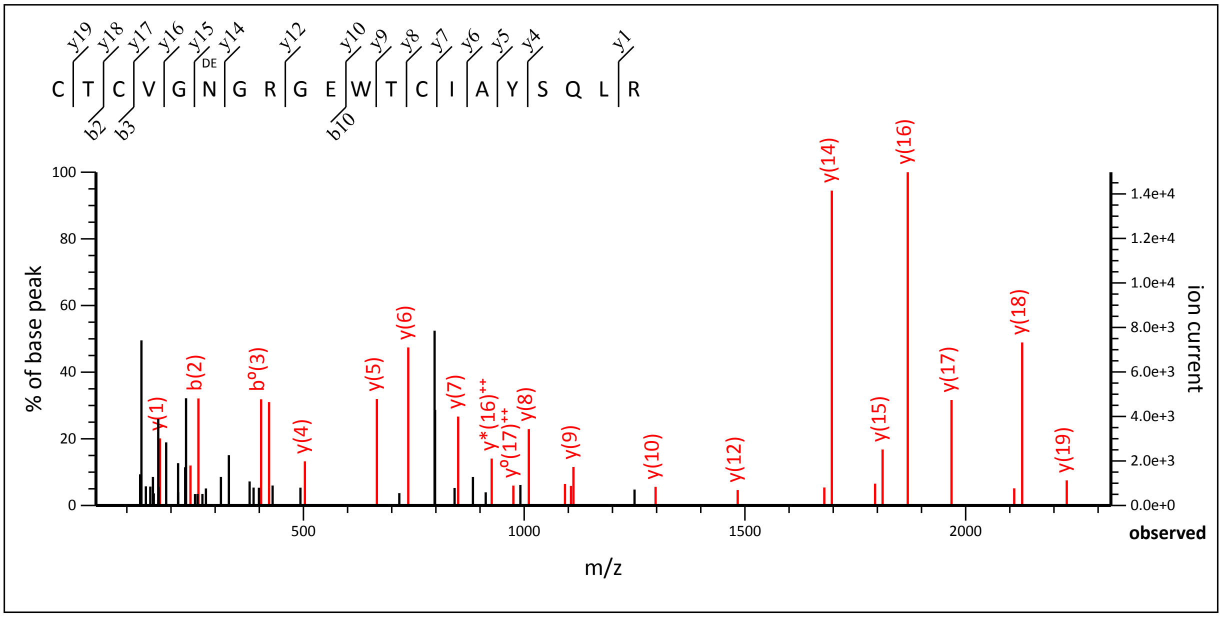


30.07 to 2329.02

Label all possible matches ○ Label matches used for scoring ●

**Monoisotopic mass of neutral peptide Mr(calc):** 2388.0409
**Fixed modifications:** Carbamidomethyl (C) (apply to specified residues or termini only)
**Variable modifications:**
**N6** : Deamidated (NQ)
**Ions Score:** 121 **Expect:** 9.9e-011
**Matches :** 26/210 fragment ions using 38 most intense peaks (help)

| # | b | $b^{++}$ | b* | $b^{*++}$ | $b^{0}$ | $b^{0++}$ | Seq. | y | $y^{++}$ | y* | $y^{*++}$ | $y^{0}$ | $y^{0++}$ | # |
|---|---|---|---|---|---|---|---|---|---|---|---|---|---|---|
| 1 | 161.0379 | 81.0226 | | | | | C | | | | | | | 20 |
| 2 | 262.0856 | 131.5464 | | | 244.0750 | 122.5412 | T | 2229.0176 | 1115.0124 | 2211.9910 | 1106.4991 | 2211.0070 | 1106.0071 | 19 |
| 3 | 422.1163 | 211.5618 | | | 404.1057 | 202.5565 | C | 2127.9699 | 1064.4886 | 2110.9433 | 1055.9753 | 2109.9593 | 1055.4833 | 18 |
| 4 | 521.1847 | 261.0960 | | | 503.1741 | 252.0907 | V | 1967.9392 | 984.4732 | 1950.9127 | 975.9600 | 1949.9287 | 975.4680 | 17 |
| 5 | 578.2061 | 289.6067 | | | 560.1956 | 280.6014 | G | 1868.8708 | 934.9390 | 1851.8443 | 926.4258 | 1850.8602 | 925.9338 | 16 |
| 6 | 693.2331 | 347.1202 | 676.2065 | 338.6069 | 675.2225 | 338.1149 | N | 1811.8493 | 906.4283 | 1794.8228 | 897.9150 | 1793.8388 | 897.4230 | 15 |
| 7 | 750.2545 | 375.6309 | 733.2280 | 367.1176 | 732.2440 | 366.6256 | G | 1696.8224 | 848.9148 | 1679.7959 | 840.4016 | 1678.8118 | 839.9096 | 14 |
| 8 | 906.3556 | 453.6815 | 889.3291 | 445.1682 | 888.3451 | 444.6762 | R | 1639.8009 | 820.4041 | 1622.7744 | 811.8908 | 1621.7904 | 811.3988 | 13 |
| 9 | 963.3771 | 482.1922 | 946.3506 | 473.6789 | 945.3665 | 473.1869 | G | 1483.6998 | 742.3536 | 1466.6733 | 733.8403 | 1465.6893 | 733.3483 | 12 |
| 10 | 1092.4197 | 546.7135 | 1075.3932 | 538.2002 | 1074.4091 | 537.7082 | E | 1426.6784 | 713.8428 | 1409.6518 | 705.3295 | 1408.6678 | 704.8375 | 11 |
| 11 | 1278.4990 | 639.7531 | 1261.4725 | 631.2399 | 1260.4885 | 630.7479 | W | 1297.6358 | 649.3215 | 1280.6092 | 640.8082 | 1279.6252 | 640.3162 | 10 |
| 12 | 1379.5467 | 690.2770 | 1362.5201 | 681.7637 | 1361.5361 | 681.2717 | T | 1111.5565 | 556.2819 | 1094.5299 | 547.7686 | 1093.5459 | 547.2766 | 9 |
| 13 | 1539.5773 | 770.2923 | 1522.5508 | 761.7790 | 1521.5668 | 761.2870 | C | 1010.5088 | 505.7580 | 993.4822 | 497.2448 | 992.4982 | 496.7527 | 8 |
| 14 | 1652.6614 | 826.8343 | 1635.6349 | 818.3211 | 1634.6508 | 817.8291 | I | 850.4781 | 425.7427 | 833.4516 | 417.2294 | 832.4676 | 416.7374 | 7 |
| 15 | 1723.6985 | 862.3529 | 1706.6720 | 853.8396 | 1705.6880 | 853.3476 | A | 737.3941 | 369.2007 | 720.3675 | 360.6874 | 719.3835 | 360.1954 | 6 |
| 16 | 1886.7619 | 943.8846 | 1869.7353 | 935.3713 | 1868.7513 | 934.8793 | Y | 666.3570 | 333.6821 | 649.3304 | 325.1688 | 648.3464 | 324.6768 | 5 |
| 17 | 1973.7939 | 987.4006 | 1956.7673 | 978.8873 | 1955.7833 | 978.3953 | S | 503.2936 | 252.1504 | 486.2671 | 243.6372 | 485.2831 | 243.1452 | 4 |

| | | | | | | | | | | | | | | |
|---|---|---|---|---|---|---|---|---|---|---|---|---|---|---|
| **18** | 2101.8525 | 1051.4299 | 2084.8259 | 1042.9166 | 2083.8419 | 1042.4246 | **Q** | 416.2616 | 208.6344 | 399.2350 | 200.1212 | | | **3** |
| **19** | 2214.9365 | 1107.9719 | 2197.9100 | 1099.4586 | 2196.9260 | 1098.9666 | **L** | 288.2030 | 144.6051 | 271.1765 | 136.0919 | | | **2** |
| **20** | | | | | | | **R** | *175.1190* | 88.0631 | 158.0924 | 79.5498 | | | **1** |

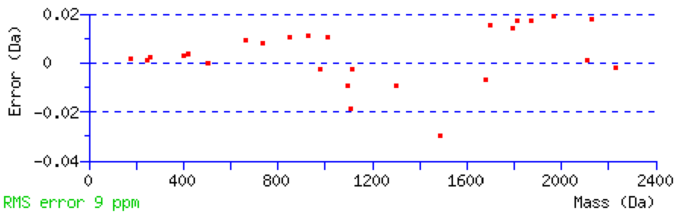


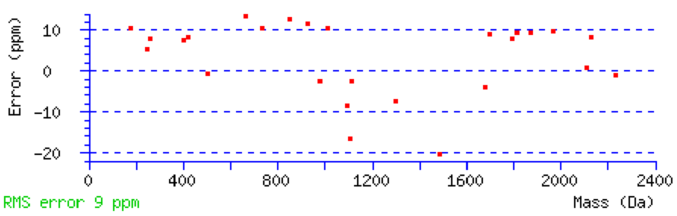


NCBI **BLAST** search of CTCVGNGRGEWTCIAYSQLR
(Parameters: blastp, nr protein database, expect=20000, no filter, PAM30)
Other BLAST web gateways

**All matches to this query**

| Score | Mr(calc) | Delta | Sequence | Site Analysis |
|---|---|---|---|---|
| 121.0 | 2388.0409 | 0.0008 | CTCVGNGRGEWTCIAYSQLR | Deamidated N6 100.00% |
| 17.1 | 2388.0409 | 0.0008 | CTCVGNGRGEWTCIAYSQLR | Deamidated Q18 0.00% |

| |
|---|
| **Mascot:** http://www.matrixscience.com/ |
