## Supplementary Data 4 for "Aging-induced isoDGR-modified fibronectin activates monocytic and endothelial cells to promote atherosclerosis"

### MATRIX SCIENCE Mascot Search Results

#### Peptide View

MS/MS Fragmentation of **GNLLQCICTGNGR**
Found in **sp|P02751|FINC_HUMAN** in **uni_human_i**, sp|P02751|FINC_HUMAN Fibronectin OS=Homo sapiens GN=FN1 PE=1 SV=4

Match to Query 9618: 1461.682588 from(731.848570,2+) intensity(37167468.0000) rtinseconds(1180) scans(8104) index(12890)
Title: Fibronectin_ERLIC_MSMS_60min_Spectrum052022_scans__8104_RTINSECONDS=1180
Data file V:\\raw\\Cam\\Fibronetin_ERLIC\\mgf\\T\\TFibronectin_ERLIC_MSMS_60min.mgf

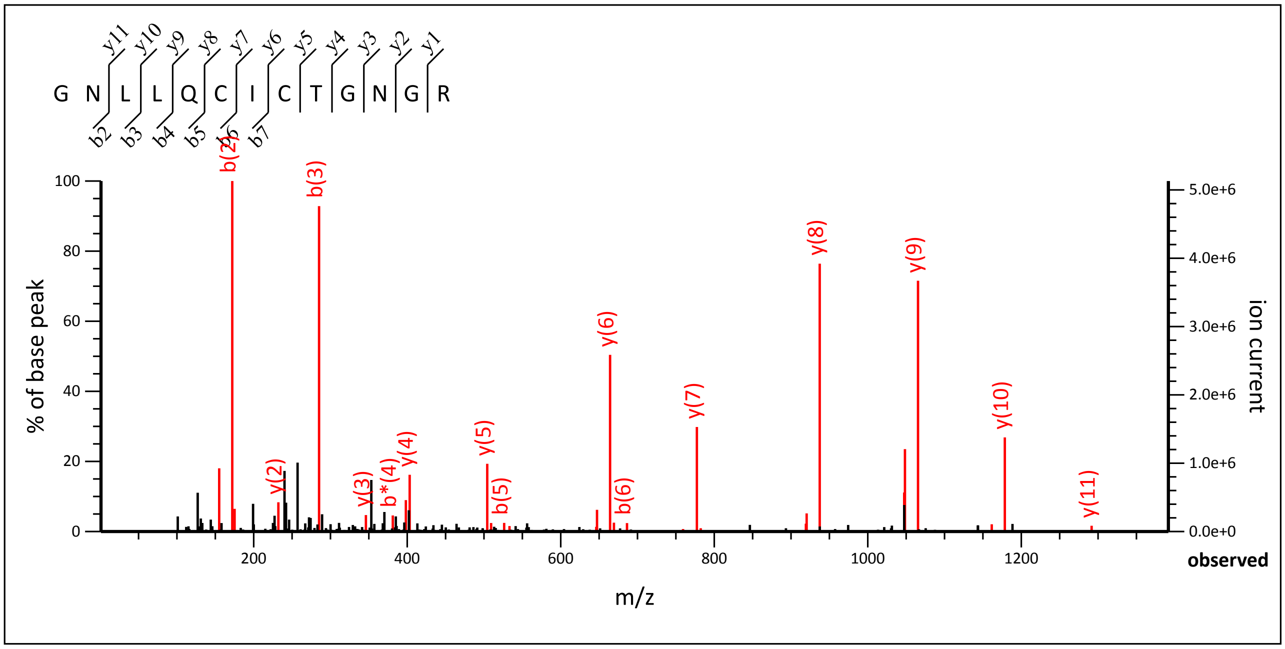


1.07 to 1391.62

Label all possible matches ○ Label matches used for scoring ◉

**Monoisotopic mass of neutral peptide Mr(calc):** 1461.6817
**Fixed modifications:** Carbamidomethyl (C) (apply to specified residues or termini only)
**Ions Score:** 82 **Expect:** 8.4e-007
**Matches :** 30/118 fragment ions using 51 most intense peaks (help)

| # | b | $b^{++}$ | b* | $b^{*++}$ | $b^0$ | $b^{0++}$ | Seq. | y | $y^{++}$ | y* | $y^{*++}$ | $y^0$ | $y^{0++}$ | # |
|---|---|---|---|---|---|---|---|---|---|---|---|---|---|---|
| 1 | 58.0287 | 29.5180 | | | | | G | | | | | | | 13 |
| 2 | **172.0717** | 86.5395 | 155.0451 | 78.0262 | | | N | 1405.6675 | 703.3374 | 1388.6409 | 694.8241 | 1387.6569 | 694.3321 | 12 |
| 3 | **285.1557** | 143.0815 | 268.1292 | 134.5682 | | | L | ***1291.6246*** | 646.3159 | 1274.5980 | 637.8026 | 1273.6140 | 637.3106 | 11 |
| 4 | **398.2398** | 199.6235 | 381.2132 | 191.1103 | | | L | ***1178.5405*** | 589.7739 | 1161.5139 | 581.2606 | 1160.5299 | 580.7686 | 10 |
| 5 | **526.2984** | 263.6528 | 509.2718 | 255.1396 | | | Q | ***1065.4564*** | **533.2319** | 1048.4299 | 524.7186 | 1047.4459 | 524.2266 | 9 |
| 6 | **686.3290** | 343.6681 | 669.3025 | 335.1549 | | | C | ***937.3978*** | 469.2026 | 920.3713 | 460.6893 | 919.3873 | 460.1973 | 8 |
| 7 | 799.4131 | 400.2102 | 782.3865 | 391.6969 | | | I | ***777.3672*** | 389.1872 | 760.3406 | 380.6740 | 759.3566 | 380.1820 | 7 |
| 8 | 959.4437 | 480.2255 | 942.4172 | 471.7122 | | | C | ***664.2831*** | 332.6452 | 647.2566 | 324.1319 | 646.2726 | 323.6399 | 6 |
| 9 | 1060.4914 | 530.7493 | 1043.4649 | 522.2361 | 1042.4808 | 521.7441 | T | ***504.2525*** | 252.6299 | 487.2259 | 244.1166 | 486.2419 | 243.6246 | 5 |
| 10 | 1117.5129 | 559.2601 | 1100.4863 | 550.7468 | 1099.5023 | 550.2548 | G | ***403.2048*** | 202.1060 | 386.1783 | 193.5928 | | | 4 |
| 11 | 1231.5558 | 616.2815 | 1214.5293 | 607.7683 | 1213.5452 | 607.2763 | N | ***346.1833*** | 173.5953 | 329.1568 | 165.0820 | | | 3 |
| 12 | 1288.5773 | 644.7923 | 1271.5507 | 636.2790 | 1270.5667 | 635.7870 | G | ***232.1404*** | 116.5738 | 215.1139 | 108.0606 | | | 2 |
| 13 | | | | | | | R | ***175.1190*** | 88.0631 | 158.0924 | 79.5498 | | | 1 |

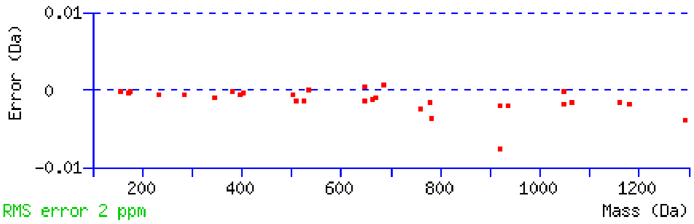


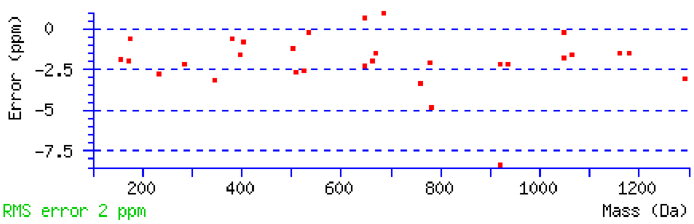


NCBI **BLAST** search of GNLLQCICTGNGR
(Parameters: blastp, nr protein database, expect=20000, no filter, PAM30)
Other BLAST web gateways

**All matches to this query**

| Score | Mr(calc) | Delta | Sequence |
|---|---|---|---|
| 82.0 | 1461.6817 | 0.0009 | GNLLQCICTGNGR |
| 4.6 | 1461.6783 | 0.0043 | QRLQMYNSQHR |

**Mascot:** http://www.matrixscience.com/

### MATRIX SCIENCE Mascot Search Results

#### Peptide View

MS/MS Fragmentation of **GNLLQCICTGNGR**
Found in **sp|P02751|FINC_HUMAN** in **uni_human_i**, sp|P02751|FINC_HUMAN Fibronectin OS=Homo sapiens GN=FN1 PE=1 SV=4

Match to Query 10319: 1462.664528 from(732.339540,2+) intensity(2065385.3750) rtinseconds(1715) scans(11923) index(18482)
Title: Fibronectin_ERLIC_MSMS_60min_Spectrum057614_scans__11923_RTINSECONDS=1715
Data file V:\\raw\\Cam\\Fibronetin_ERLIC\\mgf\\T\\TFibronectin_ERLIC_MSMS_60min.mgf

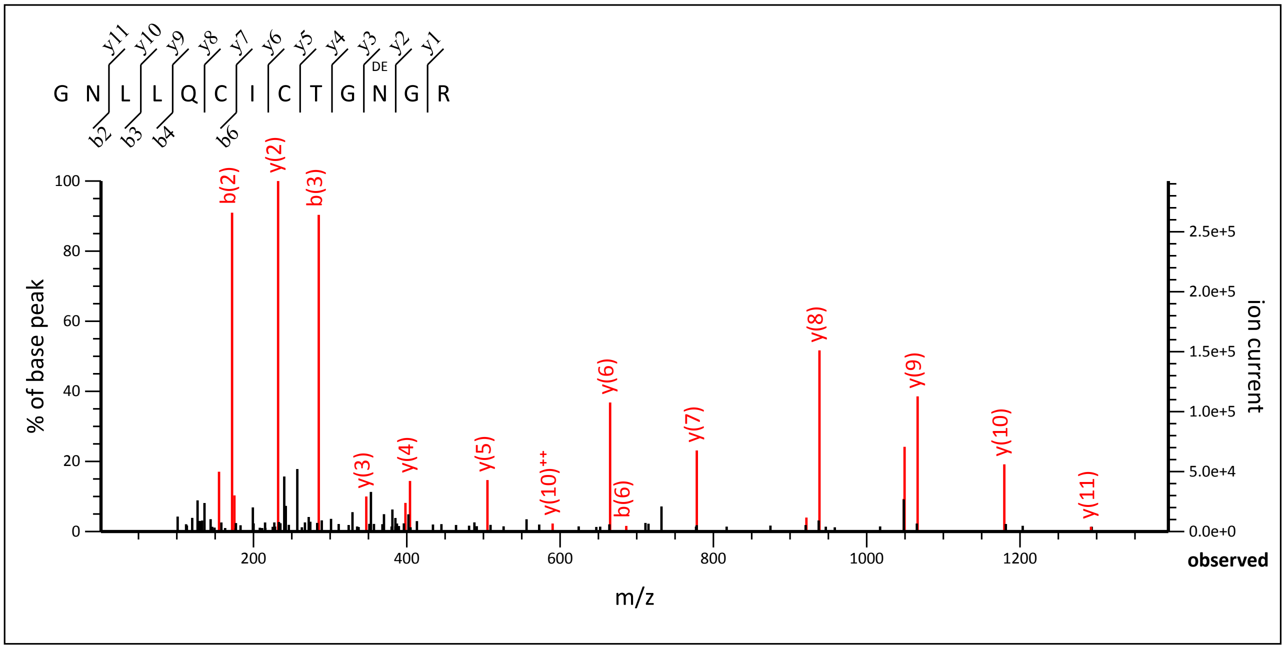


1.07 to 1393.57

Label all possible matches ○ Label matches used for scoring ◉

```
Monoisotopic mass of neutral peptide Mr(calc): 1462.6657
Fixed modifications: Carbamidomethyl (C) (apply to specified residues or termini only)
Variable modifications:
N11    : Deamidated (NQ)
Ions Score: 85  Expect: 2.8e-007
Matches : 20/118 fragment ions using 34 most intense peaks   (help)
```

| # | b | b++ | b* | b*++ | b0 | b0++ | Seq. | y | y++ | y* | y*++ | y0 | y0++ | # |
|---|---|---|---|---|---|---|---|---|---|---|---|---|---|---|
| 1 | 58.0287 | 29.5180 | | | | | G | | | | | | | 13 |
| 2 | **172.0717** | 86.5395 | 155.0451 | 78.0262 | | | N | 1406.6515 | 703.8294 | 1389.6249 | 695.3161 | 1388.6409 | 694.8241 | 12 |
| 3 | **285.1557** | 143.0815 | 268.1292 | 134.5682 | | | L | ***1292.6086*** | 646.8079 | 1275.5820 | 638.2946 | 1274.5980 | 637.8026 | 11 |
| 4 | **398.2398** | 199.6235 | 381.2132 | 191.1103 | | | L | ***1179.5245*** | **590.2659** | 1162.4980 | 581.7526 | 1161.5139 | 581.2606 | 10 |
| 5 | 526.2984 | 263.6528 | 509.2718 | 255.1396 | | | Q | ***1066.4404*** | 533.7239 | 1049.4139 | 525.2106 | 1048.4299 | 524.7186 | 9 |
| 6 | **686.3290** | 343.6681 | 669.3025 | 335.1549 | | | C | ***938.3819*** | 469.6946 | 921.3553 | 461.1813 | 920.3713 | 460.6893 | 8 |
| 7 | 799.4131 | 400.2102 | 782.3865 | 391.6969 | | | I | ***778.3512*** | 389.6792 | 761.3247 | 381.1660 | 760.3406 | 380.6740 | 7 |
| 8 | 959.4437 | 480.2255 | 942.4172 | 471.7122 | | | C | ***665.2672*** | 333.1372 | 648.2406 | 324.6239 | 647.2566 | 324.1319 | 6 |
| 9 | 1060.4914 | 530.7493 | 1043.4649 | 522.2361 | 1042.4808 | 521.7441 | T | ***505.2365*** | 253.1219 | 488.2100 | 244.6086 | 487.2259 | 244.1166 | 5 |
| 10 | 1117.5129 | 559.2601 | 1100.4863 | 550.7468 | 1099.5023 | 550.2548 | G | ***404.1888*** | 202.5980 | 387.1623 | 194.0848 | | | 4 |
| 11 | 1232.5398 | 616.7735 | 1215.5133 | 608.2603 | 1214.5293 | 607.7683 | N | ***347.1674*** | 174.0873 | 330.1408 | 165.5740 | | | 3 |
| 12 | 1289.5613 | 645.2843 | 1272.5347 | 636.7710 | 1271.5507 | 636.2790 | G | ***232.1404*** | 116.5738 | 215.1139 | 108.0606 | | | 2 |
| 13 | | | | | | | R | ***175.1190*** | 88.0631 | 158.0924 | 79.5498 | | | 1 |

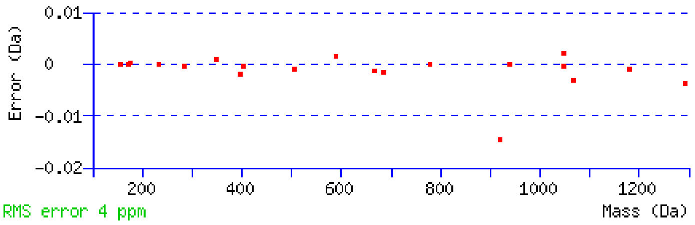


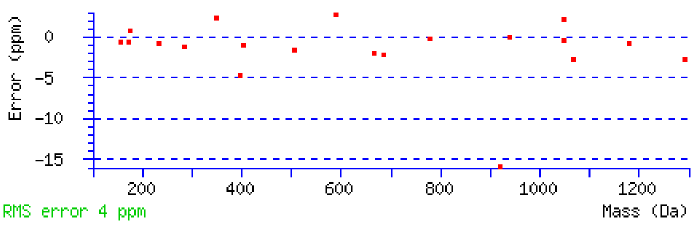


NCBI **BLAST** search of GNLLQCICTGNGR
(Parameters: blastp, nr protein database, expect=20000, no filter, PAM30)
Other BLAST web gateways

**All matches to this query**

| Score | Mr(calc) | Delta | Sequence | Site Analysis |
|---|---|---|---|---|
| 85.1 | 1462.6657 | -0.0012 | GNLLQCICTGNGR | Deamidated N11 100.00% |
| 31.6 | 1462.6657 | -0.0012 | GNLLQCICTGNGR | Deamidated Q5 0.00% |
| 4.7 | 1462.6657 | -0.0012 | GNLLQCICTGNGR | Deamidated N2 0.00% |

**Mascot:** http://www.matrixscience.com/

### MATRIX SCIENCE Mascot Search Results

#### Peptide View

MS/MS Fragmentation of **NSITLTNLTPGTEYVVSIVALNGR**
Found in **sp|P02751|FINC_HUMAN** in **uni_human_i**, sp|P02751|FINC_HUMAN Fibronectin OS=Homo sapiens GN=FN1 PE=1 SV=4

Match to Query 23984: 2531.363128 from(1266.688840,2+) intensity(5690264.0000) rtinseconds(1223) scans(8413) index(13365)
Title: Fibronectin_ERLIC_MSMS_60min_Spectrum052497_scans__8413_RTINSECONDS=1223
Data file V:\\raw\\Cam\\Fibronetin_ERLIC\\mgf\\T\\TFibronectin_ERLIC_MSMS_60min.mgf

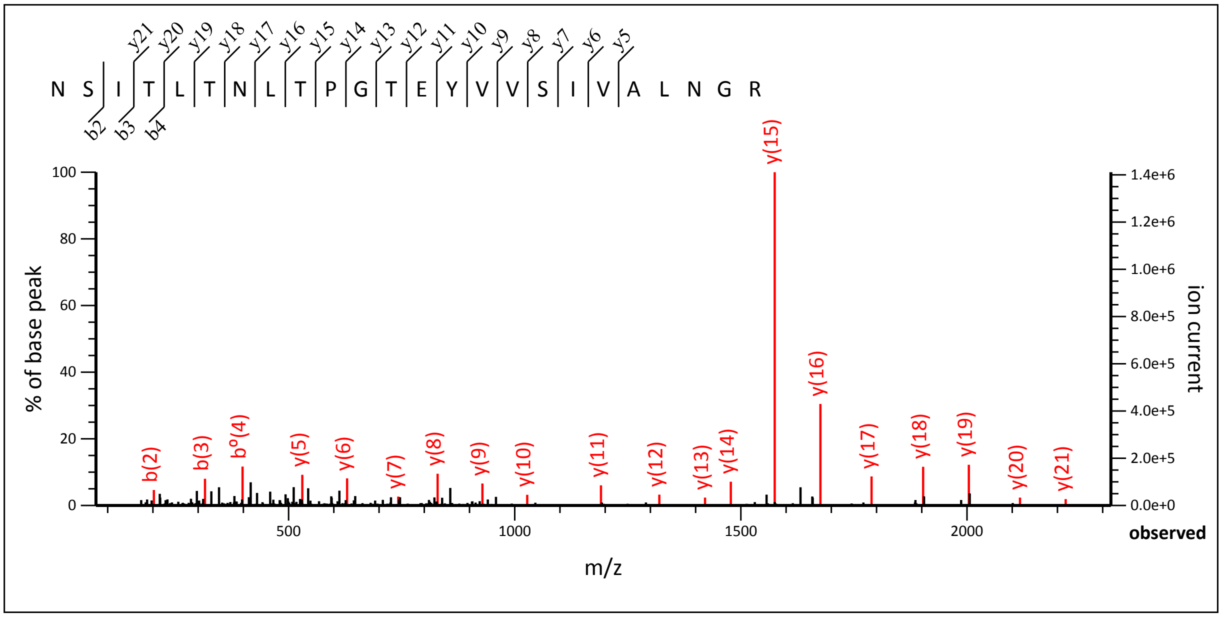


74.09 to 2318.21

Label all possible matches ○ Label matches used for scoring ◉

**Monoisotopic mass of neutral peptide Mr(calc):** 2531.3595
**Fixed modifications:** Carbamidomethyl (C) (apply to specified residues or termini only)
**Ions Score:** 192 **Expect:** 6.2e-018
**Matches :** 20/260 fragment ions using 20 most intense peaks (help)

| # | b | b$^{++}$ | b* | b*$^{++}$ | b$^{0}$ | b$^{0++}$ | Seq. | y | y$^{++}$ | y* | y*$^{++}$ | y$^{0}$ | y$^{0++}$ | # |
|---|---|---|---|---|---|---|---|---|---|---|---|---|---|---|
| 1 | 115.0502 | 58.0287 | 98.0237 | 49.5155 | | | N | | | | | | | 24 |
| 2 | **202.0822** | 101.5448 | 185.0557 | 93.0315 | 184.0717 | 92.5395 | S | 2418.3239 | 1209.6656 | 2401.2973 | 1201.1523 | 2400.3133 | 1200.6603 | 23 |
| 3 | **315.1663** | 158.0868 | 298.1397 | 149.5735 | 297.1557 | 149.0815 | I | 2331.2918 | 1166.1496 | 2314.2653 | 1157.6363 | 2313.2813 | 1157.1443 | 22 |
| 4 | 416.2140 | 208.6106 | 399.1874 | 200.0974 | 398.2034 | 199.6053 | T | ***2218.2078*** | 1109.6075 | 2201.1812 | 1101.0943 | 2200.1972 | 1100.6022 | 21 |
| 5 | 529.2980 | 265.1527 | 512.2715 | 256.6394 | 511.2875 | 256.1474 | L | ***2117.1601*** | 1059.0837 | 2100.1335 | 1050.5704 | 2099.1495 | 1050.0784 | 20 |
| 6 | 630.3457 | 315.6765 | 613.3192 | 307.1632 | 612.3352 | 306.6712 | T | ***2004.0760*** | 1002.5417 | 1987.0495 | 994.0284 | 1986.0655 | 993.5364 | 19 |
| 7 | 744.3886 | 372.6980 | 727.3621 | 364.1847 | 726.3781 | 363.6927 | N | ***1903.0284*** | 952.0178 | 1886.0018 | 943.5045 | 1885.0178 | 943.0125 | 18 |
| 8 | 857.4727 | 429.2400 | 840.4462 | 420.7267 | 839.4621 | 420.2347 | L | ***1788.9854*** | 894.9964 | 1771.9589 | 886.4831 | 1770.9749 | 885.9911 | 17 |
| 9 | 958.5204 | 479.7638 | 941.4938 | 471.2506 | 940.5098 | 470.7585 | T | ***1675.9014*** | 838.4543 | 1658.8748 | 829.9410 | 1657.8908 | 829.4490 | 16 |
| 10 | 1055.5732 | 528.2902 | 1038.5466 | 519.7769 | 1037.5626 | 519.2849 | P | ***1574.8537*** | 787.9305 | 1557.8271 | 779.4172 | 1556.8431 | 778.9252 | 15 |
| 11 | 1112.5946 | 556.8009 | 1095.5681 | 548.2877 | 1094.5840 | 547.7957 | G | ***1477.8009*** | 739.4041 | 1460.7744 | 730.8908 | 1459.7904 | 730.3988 | 14 |
| 12 | 1213.6423 | 607.3248 | 1196.6157 | 598.8115 | 1195.6317 | 598.3195 | T | ***1420.7795*** | 710.8934 | 1403.7529 | 702.3801 | 1402.7689 | 701.8881 | 13 |
| 13 | 1342.6849 | 671.8461 | 1325.6583 | 663.3328 | 1324.6743 | 662.8408 | E | ***1319.7318*** | 660.3695 | 1302.7052 | 651.8563 | 1301.7212 | 651.3642 | 12 |
| 14 | 1505.7482 | 753.3777 | 1488.7217 | 744.8645 | 1487.7377 | 744.3725 | Y | ***1190.6892*** | 595.8482 | 1173.6626 | 587.3350 | 1172.6786 | 586.8429 | 11 |
| 15 | 1604.8166 | 802.9120 | 1587.7901 | 794.3987 | 1586.8061 | 793.9067 | V | ***1027.6259*** | 514.3166 | 1010.5993 | 505.8033 | 1009.6153 | 505.3113 | 10 |
| 16 | 1703.8850 | 852.4462 | 1686.8585 | 843.9329 | 1685.8745 | 843.4409 | V | ***928.5574*** | 464.7824 | 911.5309 | 456.2691 | 910.5469 | 455.7771 | 9 |
| 17 | 1790.9171 | 895.9622 | 1773.8905 | 887.4489 | 1772.9065 | 886.9569 | S | ***829.4890*** | 415.2482 | 812.4625 | 406.7349 | 811.4785 | 406.2429 | 8 |
| 18 | 1904.0011 | 952.5042 | 1886.9746 | 943.9909 | 1885.9906 | 943.4989 | I | ***742.4570*** | 371.7321 | 725.4305 | 363.2189 | | | 7 |

| | | | | | | | | | | | | | | |
|---|---|---|---|---|---|---|---|---|---|---|---|---|---|---|
| **19** | 2003.0696 | 1002.0384 | 1986.0430 | 993.5251 | 1985.0590 | 993.0331 | **V** | ***629.3729*** | 315.1901 | 612.3464 | 306.6768 | | | **6** |
| **20** | 2074.1067 | 1037.5570 | 2057.0801 | 1029.0437 | 2056.0961 | 1028.5517 | **A** | ***530.3045*** | 265.6559 | 513.2780 | 257.1426 | | | **5** |
| **21** | 2187.1907 | 1094.0990 | 2170.1642 | 1085.5857 | 2169.1802 | 1085.0937 | **L** | 459.2674 | 230.1373 | 442.2409 | 221.6241 | | | **4** |
| **22** | 2301.2337 | 1151.1205 | 2284.2071 | 1142.6072 | 2283.2231 | 1142.1152 | **N** | 346.1833 | 173.5953 | 329.1568 | 165.0820 | | | **3** |
| **23** | 2358.2551 | 1179.6312 | 2341.2286 | 1171.1179 | 2340.2446 | 1170.6259 | **G** | 232.1404 | 116.5738 | 215.1139 | 108.0606 | | | **2** |
| **24** | | | | | | | **R** | 175.1190 | 88.0631 | 158.0924 | 79.5498 | | | **1** |

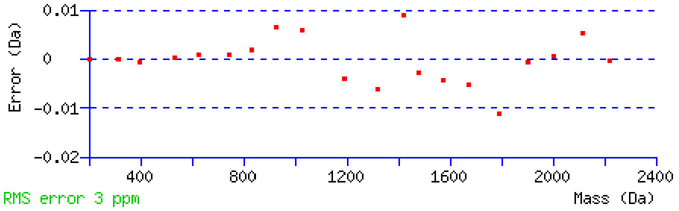


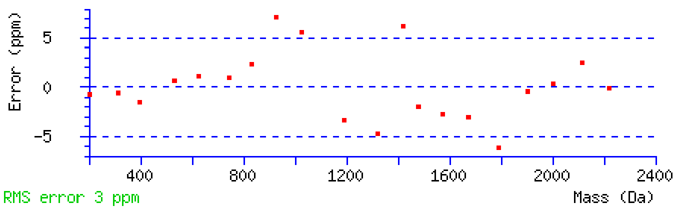


NCBI **BLAST** search of NSITLTNLTPGTEYVVSIVALNGR
(Parameters: blastp, nr protein database, expect=20000, no filter, PAM30)
Other BLAST web gateways

**All matches to this query**

| Score | Mr(calc) | Delta | Sequence |
|---|---|---|---|
| 191.9 | 2531.3595 | 0.0036 | NSITLTNLTPGTEYVVSIVALNGR |

**Mascot:** http://www.matrixscience.com/

### MATRIX SCIENCE Mascot Search Results

#### Peptide View

MS/MS Fragmentation of **NSITLTNLTPGTEYVVSIVALNGR**
Found in **sp|P02751|FINC_HUMAN** in **uni_human_i**, sp|P02751|FINC_HUMAN Fibronectin OS=Homo sapiens GN=FN1 PE=1 SV=4

Match to Query 25668: 2532.349208 from(1267.181880,2+) intensity(803011.3750) rtinseconds(1714) scans(11910) index(18465)
Title: Fibronectin_ERLIC_MSMS_60min_Spectrum057597_scans__11910_RTINSECONDS=1714
Data file V:\\raw\\Cam\\Fibronetin_ERLIC\\mgf\\T\\TFibronectin_ERLIC_MSMS_60min.mgf

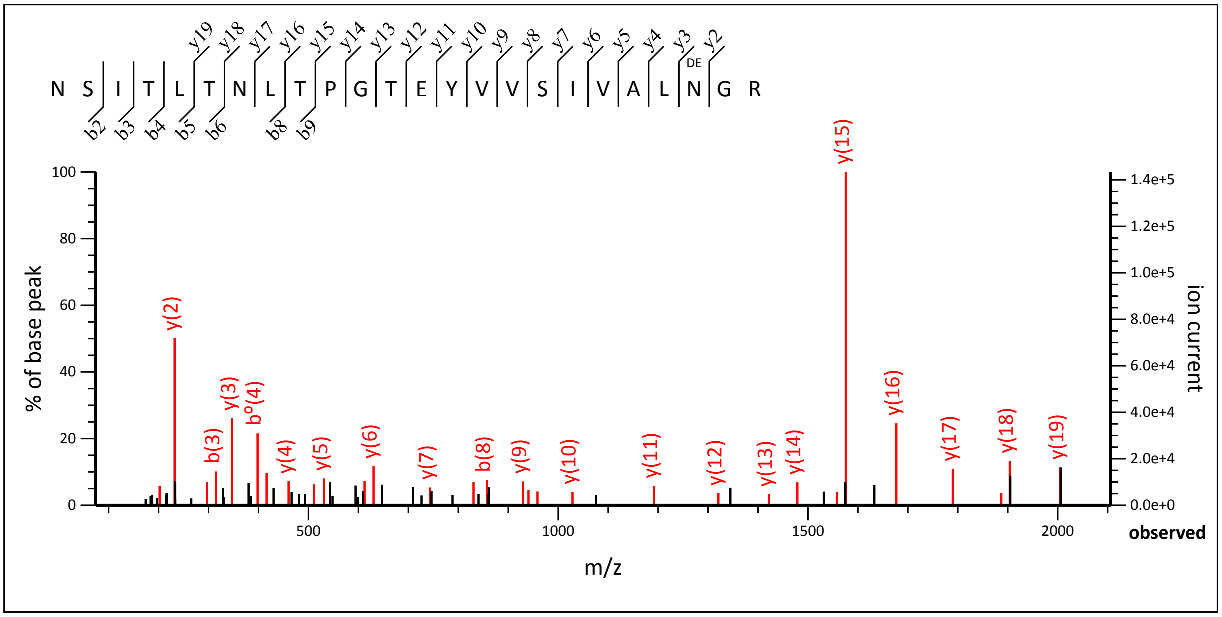


74.19 to 2106

Label all possible matches ○ Label matches used for scoring ◉

**Monoisotopic mass of neutral peptide Mr(calc):** 2532.3435
**Fixed modifications:** Carbamidomethyl (C) (apply to specified residues or termini only)
**Variable modifications:**
N22 : Deamidated (NQ)
**Ions Score:** 154 **Expect:** 5e-014
**Matches :** 33/260 fragment ions using 43 most intense peaks (help)

| # | b | $b^{++}$ | b* | $b^{*++}$ | $b^0$ | $b^{0++}$ | Seq. | y | $y^{++}$ | y* | $y^{*++}$ | $y^0$ | $y^{0++}$ | # |
|---|---|---|---|---|---|---|---|---|---|---|---|---|---|---|
| 1 | 115.0502 | 58.0287 | 98.0237 | 49.5155 | | | N | | | | | | | 24 |
| 2 | **202.0822** | 101.5448 | 185.0557 | 93.0315 | 184.0717 | 92.5395 | S | 2419.3079 | 1210.1576 | 2402.2813 | 1201.6443 | 2401.2973 | 1201.1523 | 23 |
| 3 | **315.1663** | 158.0868 | 298.1397 | 149.5735 | 297.1557 | 149.0815 | I | 2332.2759 | 1166.6416 | 2315.2493 | 1158.1283 | 2314.2653 | 1157.6363 | 22 |
| 4 | **416.2140** | 208.6106 | 399.1874 | 200.0974 | 398.2034 | 199.6053 | T | 2219.1918 | 1110.0995 | 2202.1652 | 1101.5863 | 2201.1812 | 1101.0943 | 21 |
| 5 | 529.2980 | 265.1527 | 512.2715 | 256.6394 | 511.2875 | 256.1474 | L | 2118.1441 | 1059.5757 | 2101.1176 | 1051.0624 | 2100.1335 | 1050.5704 | 20 |
| 6 | **630.3457** | 315.6765 | 613.3192 | 307.1632 | 612.3352 | 306.6712 | T | ***2005.0600*** | 1003.0337 | 1988.0335 | 994.5204 | 1987.0495 | 994.0284 | 19 |
| 7 | 744.3886 | 372.6980 | 727.3621 | 364.1847 | 726.3781 | 363.6927 | N | ***1904.0124*** | 952.5098 | 1886.9858 | 943.9965 | 1886.0018 | 943.5045 | 18 |
| 8 | **857.4727** | 429.2400 | 840.4462 | 420.7267 | 839.4621 | 420.2347 | L | ***1789.9694*** | 895.4884 | 1772.9429 | 886.9751 | 1771.9589 | 886.4831 | 17 |
| 9 | **958.5204** | 479.7638 | 941.4938 | 471.2506 | 940.5098 | 470.7585 | T | ***1676.8854*** | 838.9463 | 1659.8588 | 830.4331 | 1658.8748 | 829.9410 | 16 |
| 10 | 1055.5732 | 528.2902 | 1038.5466 | 519.7769 | 1037.5626 | 519.2849 | P | ***1575.8377*** | 788.4225 | 1558.8112 | 779.9092 | 1557.8271 | 779.4172 | 15 |
| 11 | 1112.5946 | 556.8009 | 1095.5681 | 548.2877 | 1094.5840 | 547.7957 | G | ***1478.7849*** | 739.8961 | 1461.7584 | 731.3828 | 1460.7744 | 730.8908 | 14 |
| 12 | 1213.6423 | 607.3248 | 1196.6157 | 598.8115 | 1195.6317 | 598.3195 | T | ***1421.7635*** | 711.3854 | 1404.7369 | 702.8721 | 1403.7529 | 702.3801 | 13 |
| 13 | 1342.6849 | 671.8461 | 1325.6583 | 663.3328 | 1324.6743 | 662.8408 | E | ***1320.7158*** | 660.8615 | 1303.6892 | 652.3483 | 1302.7052 | 651.8563 | 12 |
| 14 | 1505.7482 | 753.3777 | 1488.7217 | 744.8645 | 1487.7377 | 744.3725 | Y | ***1191.6732*** | 596.3402 | 1174.6467 | 587.8270 | 1173.6626 | 587.3350 | 11 |
| 15 | 1604.8166 | 802.9120 | 1587.7901 | 794.3987 | 1586.8061 | 793.9067 | V | ***1028.6099*** | 514.8086 | 1011.5833 | 506.2953 | 1010.5993 | 505.8033 | 10 |
| 16 | 1703.8850 | 852.4462 | 1686.8585 | 843.9329 | 1685.8745 | 843.4409 | V | ***929.5415*** | 465.2744 | 912.5149 | 456.7611 | 911.5309 | 456.2691 | 9 |
| 17 | 1790.9171 | 895.9622 | 1773.8905 | 887.4489 | 1772.9065 | 886.9569 | S | ***830.4730*** | 415.7402 | 813.4465 | 407.2269 | 812.4625 | 406.7349 | 8 |

| | | | | | | | | | | | | | | |
|---|---|---|---|---|---|---|---|---|---|---|---|---|---|---|
| **18** | **1904.0011** | 952.5042 | 1886.9746 | 943.9909 | 1885.9906 | 943.4989 | **I** | ***743.4410*** | 372.2241 | 726.4145 | 363.7109 | | | **7** |
| **19** | 2003.0696 | 1002.0384 | 1986.0430 | 993.5251 | 1985.0590 | 993.0331 | **V** | ***630.3570*** | 315.6821 | 613.3304 | 307.1688 | | | **6** |
| **20** | 2074.1067 | 1037.5570 | 2057.0801 | 1029.0437 | 2056.0961 | 1028.5517 | **A** | ***531.2885*** | 266.1479 | 514.2620 | 257.6346 | | | **5** |
| **21** | 2187.1907 | 1094.0990 | 2170.1642 | 1085.5857 | 2169.1802 | 1085.0937 | **L** | ***460.2514*** | 230.6293 | 443.2249 | 222.1161 | | | **4** |
| **22** | 2302.2177 | 1151.6125 | 2285.1911 | 1143.0992 | 2284.2071 | 1142.6072 | **N** | ***347.1674*** | 174.0873 | 330.1408 | 165.5740 | | | **3** |
| **23** | 2359.2391 | 1180.1232 | 2342.2126 | 1171.6099 | 2341.2286 | 1171.1179 | **G** | ***232.1404*** | 116.5738 | 215.1139 | 108.0606 | | | **2** |
| **24** | | | | | | | **R** | 175.1190 | 88.0631 | 158.0924 | 79.5498 | | | **1** |

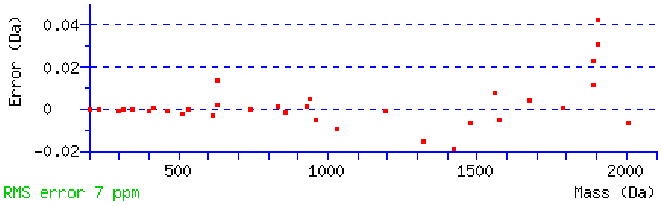


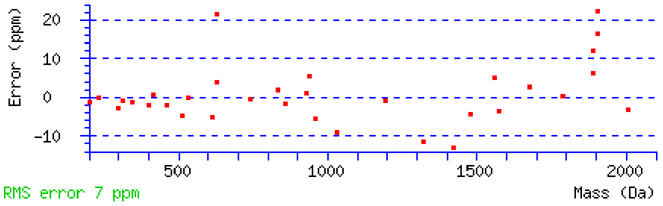


NCBI **BLAST** search of NSITLTNLTPGTEYVVSIVALNGR
(Parameters: blastp, nr protein database, expect=20000, no filter, PAM30)
Other BLAST web gateways

**All matches to this query**

| Score | Mr(calc) | Delta | Sequence | Site Analysis |
|---|---|---|---|---|
| 153.8 | 2532.3435 | 0.0057 | NSITLTNLTPGTEYVVSIVALNGR | Deamidated N22 100.00% |
| 11.6 | 2532.3435 | 0.0057 | NSITLTNLTPGTEYVVSIVALNGR | Deamidated N7 0.00% |
| 2.3 | 2531.3458 | 1.0034 | MAQKVFQSQGLGLLVWLGLDLGR | |
| 1.6 | 2532.3595 | -0.0103 | TGQLQTNRARATVAPLPMTPVPGR | |
| 1.6 | 2532.3595 | -0.0103 | TGQLQTNRARATVAPLPMTPVPGR | |

**Mascot:** http://www.matrixscience.com/

### MATRIX SCIENCE Mascot Search Results

#### Peptide View

MS/MS Fragmentation of **NSITLTNLTPGTEYVVSIVALNGR**
Found in **sp|P02751|FINC_HUMAN** in **uni_human_i**, sp|P02751|FINC_HUMAN Fibronectin OS=Homo sapiens GN=FN1 PE=1 SV=4

Match to Query 23138: 2531.360202 from(844.794010,3+) intensity(7201550.5000) rtinseconds(1219) scans(8384) index(13321)
Title: Fibronectin_ERLIC_MSMS_60min_Spectrum052453_scans__8384_RTINSECONDS=1219
Data file V:\\raw\\Cam\\Fibronetin_ERLIC\\mgf\\T\\TFibronectin_ERLIC_MSMS_60min.mgf

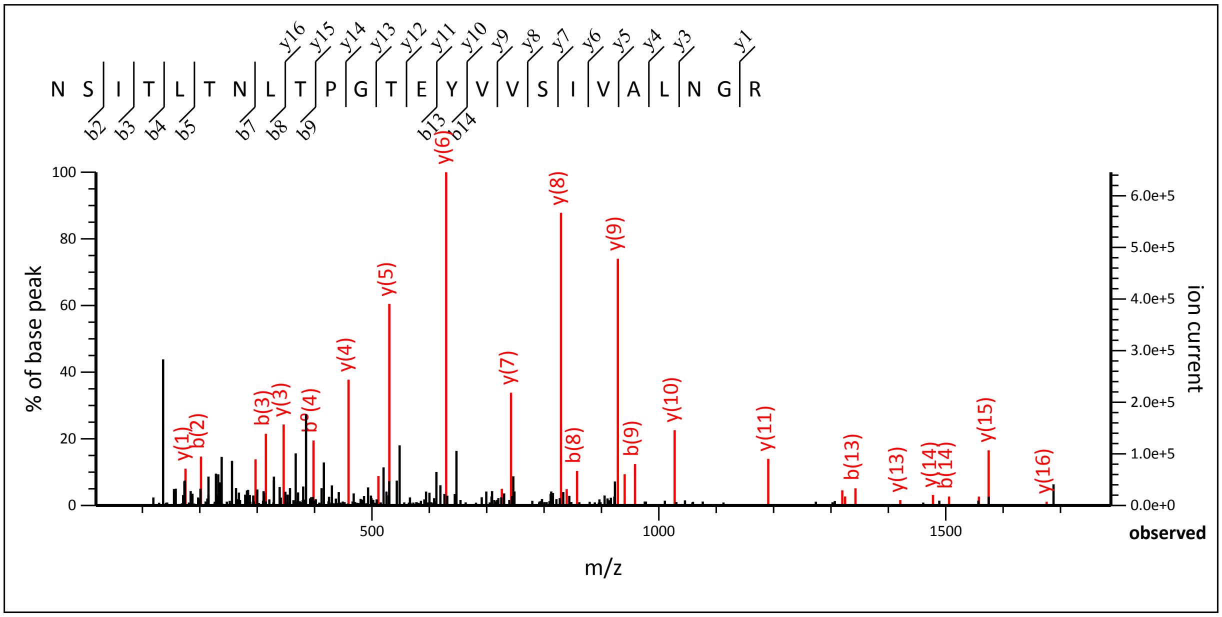


19.05 to 1787.86

Label all possible matches ○ Label matches used for scoring ●

**Monoisotopic mass of neutral peptide Mr(calc):** 2531.3595
**Fixed modifications:** Carbamidomethyl (C) (apply to specified residues or termini only)
**Ions Score:** 111 **Expect:** 7.7e-010
**Matches :** 29/260 fragment ions using 45 most intense peaks (help)

| # | b | $b^{++}$ | b* | $b^{*++}$ | $b^0$ | $b^{0++}$ | Seq. | y | $y^{++}$ | y* | $y^{*++}$ | $y^0$ | $y^{0++}$ | # |
|---|---|---|---|---|---|---|---|---|---|---|---|---|---|---|
| 1 | 115.0502 | 58.0287 | 98.0237 | 49.5155 | | | N | | | | | | | 24 |
| 2 | **202.0822** | 101.5448 | 185.0557 | 93.0315 | 184.0717 | 92.5395 | S | 2418.3239 | 1209.6656 | 2401.2973 | 1201.1523 | 2400.3133 | 1200.6603 | 23 |
| 3 | **315.1663** | 158.0868 | 298.1397 | 149.5735 | 297.1557 | 149.0815 | I | 2331.2918 | 1166.1496 | 2314.2653 | 1157.6363 | 2313.2813 | 1157.1443 | 22 |
| 4 | 416.2140 | 208.6106 | 399.1874 | 200.0974 | 398.2034 | 199.6053 | T | 2218.2078 | 1109.6075 | 2201.1812 | 1101.0943 | 2200.1972 | 1100.6022 | 21 |
| 5 | 529.2980 | 265.1527 | 512.2715 | 256.6394 | 511.2875 | 256.1474 | L | 2117.1601 | 1059.0837 | 2100.1335 | 1050.5704 | 2099.1495 | 1050.0784 | 20 |
| 6 | 630.3457 | 315.6765 | 613.3192 | 307.1632 | 612.3352 | 306.6712 | T | 2004.0760 | 1002.5417 | 1987.0495 | 994.0284 | 1986.0655 | 993.5364 | 19 |
| 7 | 744.3886 | 372.6980 | 727.3621 | 364.1847 | 726.3781 | 363.6927 | N | 1903.0284 | 952.0178 | 1886.0018 | 943.5045 | 1885.0178 | 943.0125 | 18 |
| 8 | **857.4727** | 429.2400 | 840.4462 | 420.7267 | 839.4621 | 420.2347 | L | 1788.9854 | 894.9964 | 1771.9589 | 886.4831 | 1770.9749 | 885.9911 | 17 |
| 9 | **958.5204** | 479.7638 | 941.4938 | 471.2506 | 940.5098 | 470.7585 | T | ***1675.9014*** | 838.4543 | 1658.8748 | 829.9410 | 1657.8908 | 829.4490 | 16 |
| 10 | 1055.5732 | 528.2902 | 1038.5466 | 519.7769 | 1037.5626 | 519.2849 | P | ***1574.8537*** | 787.9305 | 1557.8271 | 779.4172 | 1556.8431 | 778.9252 | 15 |
| 11 | 1112.5946 | 556.8009 | 1095.5681 | 548.2877 | 1094.5840 | 547.7957 | G | ***1477.8009*** | 739.4041 | 1460.7744 | 730.8908 | 1459.7904 | 730.3988 | 14 |
| 12 | 1213.6423 | 607.3248 | 1196.6157 | 598.8115 | 1195.6317 | 598.3195 | T | ***1420.7795*** | 710.8934 | 1403.7529 | 702.3801 | 1402.7689 | 701.8881 | 13 |
| 13 | **1342.6849** | 671.8461 | 1325.6583 | 663.3328 | 1324.6743 | 662.8408 | E | ***1319.7318*** | 660.3695 | 1302.7052 | 651.8563 | 1301.7212 | 651.3642 | 12 |
| 14 | **1505.7482** | 753.3777 | 1488.7217 | 744.8645 | 1487.7377 | 744.3725 | Y | ***1190.6892*** | 595.8482 | 1173.6626 | 587.3350 | 1172.6786 | 586.8429 | 11 |
| 15 | 1604.8166 | 802.9120 | 1587.7901 | 794.3987 | 1586.8061 | 793.9067 | V | ***1027.6259*** | 514.3166 | 1010.5993 | 505.8033 | 1009.6153 | 505.3113 | 10 |
| 16 | 1703.8850 | 852.4462 | 1686.8585 | 843.9329 | 1685.8745 | 843.4409 | V | ***928.5574*** | 464.7824 | 911.5309 | 456.2691 | 910.5469 | 455.7771 | 9 |
| 17 | 1790.9171 | 895.9622 | 1773.8905 | 887.4489 | 1772.9065 | 886.9569 | S | ***829.4890*** | 415.2482 | 812.4625 | 406.7349 | 811.4785 | 406.2429 | 8 |
| 18 | 1904.0011 | 952.5042 | 1886.9746 | 943.9909 | 1885.9906 | 943.4989 | I | ***742.4570*** | 371.7321 | 725.4305 | 363.2189 | | | 7 |

| | | | | | | | | | | | | | | |
|---|---|---|---|---|---|---|---|---|---|---|---|---|---|---|
| 19 | 2003.0696 | 1002.0384 | 1986.0430 | 993.5251 | 1985.0590 | 993.0331 | V | *629.3729* | 315.1901 | 612.3464 | 306.6768 | | | 6 |
| 20 | 2074.1067 | 1037.5570 | 2057.0801 | 1029.0437 | 2056.0961 | 1028.5517 | A | *530.3045* | 265.6559 | 513.2780 | 257.1426 | | | 5 |
| 21 | 2187.1907 | 1094.0990 | 2170.1642 | 1085.5857 | 2169.1802 | 1085.0937 | L | *459.2674* | 230.1373 | 442.2409 | 221.6241 | | | 4 |
| 22 | 2301.2337 | 1151.1205 | 2284.2071 | 1142.6072 | 2283.2231 | 1142.1152 | N | *346.1833* | 173.5953 | 329.1568 | 165.0820 | | | 3 |
| 23 | 2358.2551 | 1179.6312 | 2341.2286 | 1171.1179 | 2340.2446 | 1170.6259 | G | 232.1404 | 116.5738 | 215.1139 | 108.0606 | | | 2 |
| 24 | | | | | | | R | *175.1190* | 88.0631 | 158.0924 | 79.5498 | | | 1 |

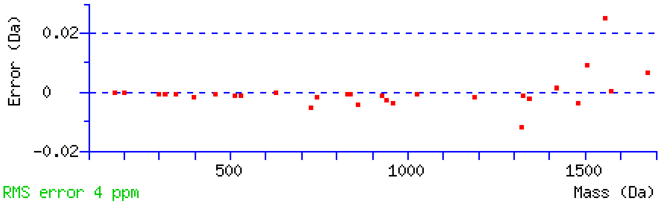


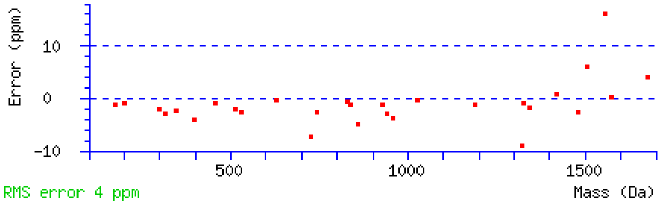


NCBI **BLAST** search of NSITLTNLTPGTEYVVSIVALNGR
(Parameters: blastp, nr protein database, expect=20000, no filter, PAM30)
Other BLAST web gateways

**All matches to this query**

| Score | Mr(calc) | Delta | Sequence |
|---|---|---|---|
| 111.2 | 2531.3595 | 0.0007 | NSITLTNLTPGTEYVVSIVALNGR |

**Mascot:** http://www.matrixscience.com/

### MATRIX SCIENCE Mascot Search Results

#### Peptide View

MS/MS Fragmentation of **NSITLTNLTPGTEYVVSIVALNGR**
Found in **sp|P02751|FINC_HUMAN** in **uni_human_i**, sp|P02751|FINC_HUMAN Fibronectin OS=Homo sapiens GN=FN1 PE=1 SV=4

Match to Query 25646: 2532.346752 from(845.122860,3+) intensity(1298501.3750) rtinseconds(1710) scans(11881) index(18426)
Title: Fibronectin_ERLIC_MSMS_60min_Spectrum057558_scans__11881_RTINSECONDS=1710
Data file V:\\raw\\Cam\\Fibronetin_ERLIC\\mgf\\T\\TFibronectin_ERLIC_MSMS_60min.mgf

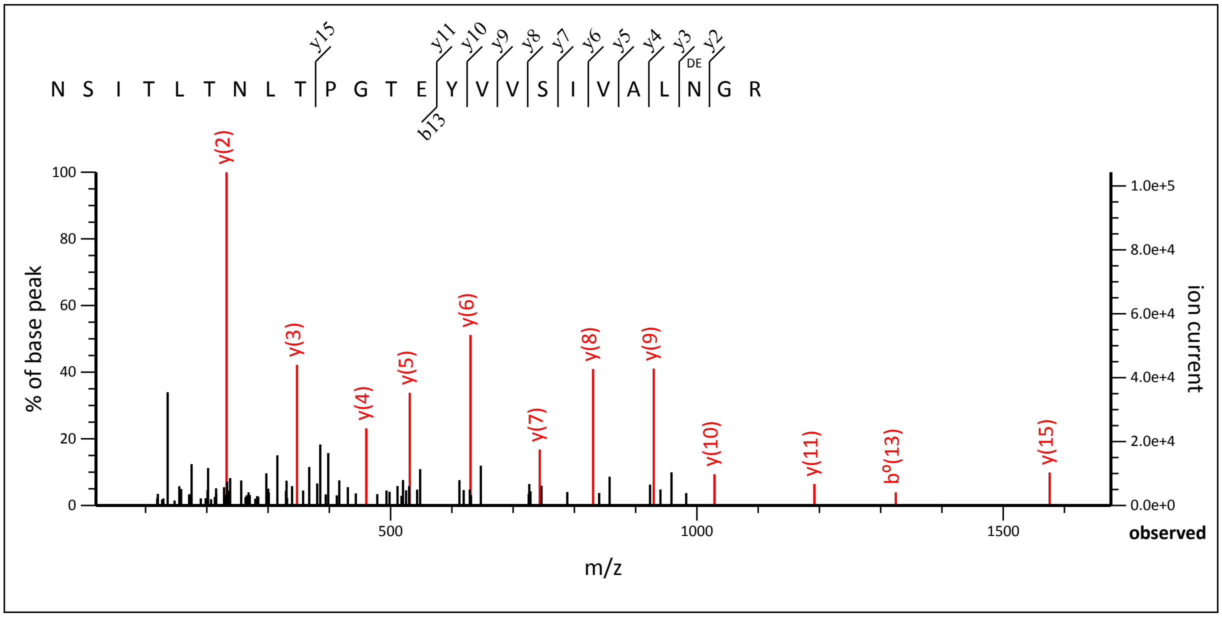


19.05 to 1675.83

Label all possible matches ○ Label matches used for scoring ◉

**Monoisotopic mass of neutral peptide Mr(calc):** 2532.3435
**Fixed modifications:** Carbamidomethyl (C) (apply to specified residues or termini only)
**Variable modifications:**
**N22** : Deamidated (NQ)
**Ions Score:** 102 **Expect:** 7.1e-009
**Matches :** 13/260 fragment ions using 13 most intense peaks (help)

| # | b | $b^{++}$ | b* | $b^{*++}$ | $b^0$ | $b^{0++}$ | Seq. | y | $y^{++}$ | y* | $y^{*++}$ | $y^0$ | $y^{0++}$ | # |
|---|---|---|---|---|---|---|---|---|---|---|---|---|---|---|
| 1 | 115.0502 | 58.0287 | 98.0237 | 49.5155 | | | N | | | | | | | 24 |
| 2 | 202.0822 | 101.5448 | 185.0557 | 93.0315 | 184.0717 | 92.5395 | S | 2419.3079 | 1210.1576 | 2402.2813 | 1201.6443 | 2401.2973 | 1201.1523 | 23 |
| 3 | 315.1663 | 158.0868 | 298.1397 | 149.5735 | 297.1557 | 149.0815 | I | 2332.2759 | 1166.6416 | 2315.2493 | 1158.1283 | 2314.2653 | 1157.6363 | 22 |
| 4 | 416.2140 | 208.6106 | 399.1874 | 200.0974 | 398.2034 | 199.6053 | T | 2219.1918 | 1110.0995 | 2202.1652 | 1101.5863 | 2201.1812 | 1101.0943 | 21 |
| 5 | 529.2980 | 265.1527 | 512.2715 | 256.6394 | 511.2875 | 256.1474 | L | 2118.1441 | 1059.5757 | 2101.1176 | 1051.0624 | 2100.1335 | 1050.5704 | 20 |
| 6 | **630.3457** | 315.6765 | 613.3192 | 307.1632 | 612.3352 | 306.6712 | T | 2005.0600 | 1003.0337 | 1988.0335 | 994.5204 | 1987.0495 | 994.0284 | 19 |
| 7 | 744.3886 | 372.6980 | 727.3621 | 364.1847 | 726.3781 | 363.6927 | N | 1904.0124 | 952.5098 | 1886.9858 | 943.9965 | 1886.0018 | 943.5045 | 18 |
| 8 | 857.4727 | 429.2400 | 840.4462 | 420.7267 | 839.4621 | 420.2347 | L | 1789.9694 | 895.4884 | 1772.9429 | 886.9751 | 1771.9589 | 886.4831 | 17 |
| 9 | 958.5204 | 479.7638 | 941.4938 | 471.2506 | 940.5098 | 470.7585 | T | 1676.8854 | 838.9463 | 1659.8588 | 830.4331 | 1658.8748 | 829.9410 | 16 |
| 10 | 1055.5732 | 528.2902 | 1038.5466 | 519.7769 | 1037.5626 | 519.2849 | P | ***1575.8377*** | 788.4225 | 1558.8112 | 779.9092 | 1557.8271 | 779.4172 | 15 |
| 11 | 1112.5946 | 556.8009 | 1095.5681 | 548.2877 | 1094.5840 | 547.7957 | G | 1478.7849 | 739.8961 | 1461.7584 | 731.3828 | 1460.7744 | 730.8908 | 14 |
| 12 | 1213.6423 | 607.3248 | 1196.6157 | 598.8115 | 1195.6317 | 598.3195 | T | 1421.7635 | 711.3854 | 1404.7369 | 702.8721 | 1403.7529 | 702.3801 | 13 |
| 13 | 1342.6849 | 671.8461 | 1325.6583 | 663.3328 | 1324.6743 | 662.8408 | E | 1320.7158 | 660.8615 | 1303.6892 | 652.3483 | 1302.7052 | 651.8563 | 12 |
| 14 | 1505.7482 | 753.3777 | 1488.7217 | 744.8645 | 1487.7377 | 744.3725 | Y | ***1191.6732*** | 596.3402 | 1174.6467 | 587.8270 | 1173.6626 | 587.3350 | 11 |
| 15 | 1604.8166 | 802.9120 | 1587.7901 | 794.3987 | 1586.8061 | 793.9067 | V | ***1028.6099*** | 514.8086 | 1011.5833 | 506.2953 | 1010.5993 | 505.8033 | 10 |
| 16 | 1703.8850 | 852.4462 | 1686.8585 | 843.9329 | 1685.8745 | 843.4409 | V | ***929.5415*** | 465.2744 | 912.5149 | 456.7611 | 911.5309 | 456.2691 | 9 |
| 17 | 1790.9171 | 895.9622 | 1773.8905 | 887.4489 | 1772.9065 | 886.9569 | S | ***830.4730*** | 415.7402 | 813.4465 | 407.2269 | 812.4625 | 406.7349 | 8 |

| | | | | | | | | | | | | | | |
|---|---|---|---|---|---|---|---|---|---|---|---|---|---|---|
| **18** | 1904.0011 | 952.5042 | 1886.9746 | 943.9909 | 1885.9906 | 943.4989 | **I** | *743.4410* | 372.2241 | 726.4145 | 363.7109 | | | **7** |
| **19** | 2003.0696 | 1002.0384 | 1986.0430 | 993.5251 | 1985.0590 | 993.0331 | **V** | *630.3570* | 315.6821 | 613.3304 | 307.1688 | | | **6** |
| **20** | 2074.1067 | 1037.5570 | 2057.0801 | 1029.0437 | 2056.0961 | 1028.5517 | **A** | *531.2885* | 266.1479 | 514.2620 | 257.6346 | | | **5** |
| **21** | 2187.1907 | 1094.0990 | 2170.1642 | 1085.5857 | 2169.1802 | 1085.0937 | **L** | *460.2514* | 230.6293 | 443.2249 | 222.1161 | | | **4** |
| **22** | 2302.2177 | 1151.6125 | 2285.1911 | 1143.0992 | 2284.2071 | 1142.6072 | **N** | *347.1674* | 174.0873 | 330.1408 | 165.5740 | | | **3** |
| **23** | 2359.2391 | 1180.1232 | 2342.2126 | 1171.6099 | 2341.2286 | 1171.1179 | **G** | *232.1404* | 116.5738 | 215.1139 | 108.0606 | | | **2** |
| **24** | | | | | | | **R** | 175.1190 | 88.0631 | 158.0924 | 79.5498 | | | **1** |

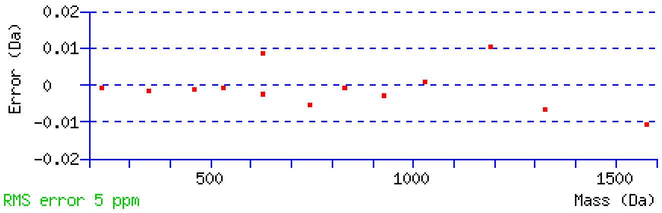


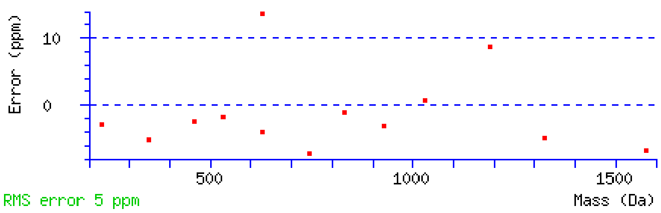


NCBI **BLAST** search of NSITLTNLTPGTEYVVSIVALNGR
(Parameters: blastp, nr protein database, expect=20000, no filter, PAM30)
Other BLAST web gateways

**All matches to this query**

| Score | Mr(calc) | Delta | Sequence | Site Analysis |
|---|---|---|---|---|
| 102.4 | 2532.3435 | 0.0032 | NSITLTNLTPGTEYVVSIVALNGR | Deamidated N22 100.00% |
| 26.8 | 2531.3458 | 1.0010 | MAQKVFQSQGLGLLVWLGLDLGR | |
| 0.3 | 2532.3435 | 0.0032 | NSITLTNLTPGTEYVVSIVALNGR | Deamidated N7 0.00% |

**Mascot:** http://www.matrixscience.com/

### MATRIX SCIENCE Mascot Search Results

#### Peptide View

MS/MS Fragmentation of **NSITLTNLTPGTEYVVSIVALNGREESPLLIGQQSTVSDVPR**
Found in **sp|P02751|FINC_HUMAN** in **uni_human_i**, sp|P02751|FINC_HUMAN Fibronectin OS=Homo sapiens GN=FN1 PE=1 SV=4

Match to Query 38991: 4468.357616 from(1118.096680,4+) intensity(334781.1875) rtinseconds(1876) scans(13057) index(20100)
Title: Fibronectin_ERLIC_MSMS_60min_Spectrum059232_scans__13057_RTINSECONDS=1876
Data file V:\\raw\\Cam\\Fibronetin_ERLIC\\mgf\\T\\TFibronectin_ERLIC_MSMS_60min.mgf

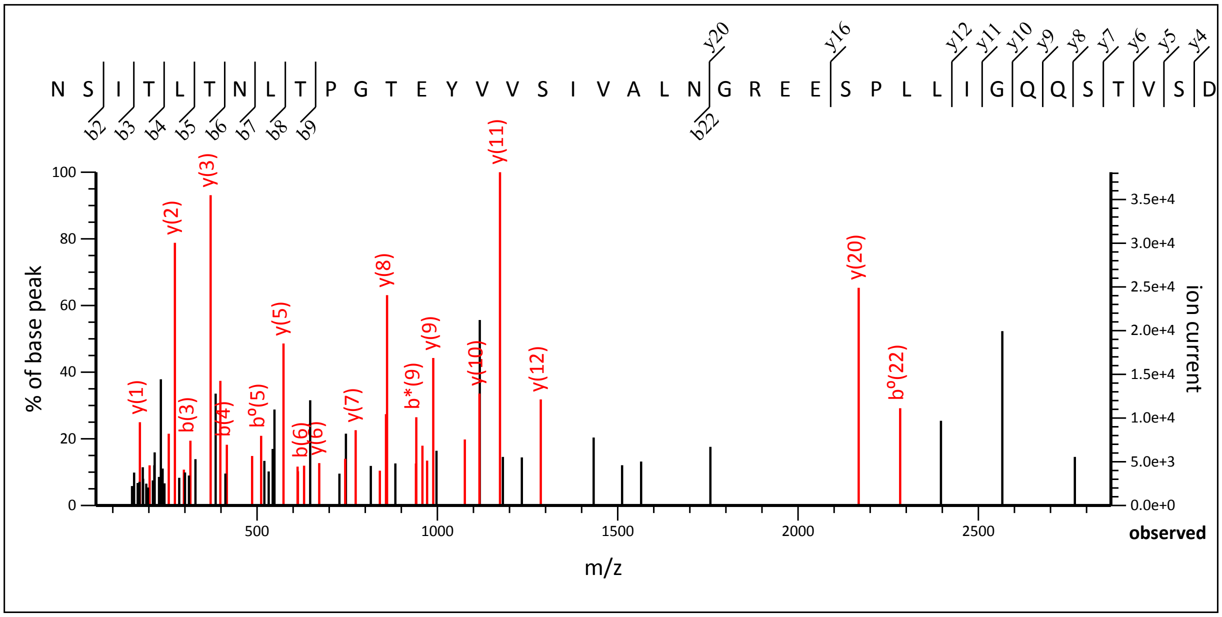


53.35 to 2867.49

Label all possible matches ○ Label matches used for scoring ◉

**Monoisotopic mass of neutral peptide Mr(calc):** 4467.3497
**Fixed modifications:** Carbamidomethyl (C) (apply to specified residues or termini only)
**Ions Score:** 68 **Expect:** 1.5e-005
**Matches :** 34/484 fragment ions using 57 most intense peaks (help)

| # | b | $b^{++}$ | b* | $b^{*++}$ | $b^0$ | $b^{0++}$ | Seq. | y | $y^{++}$ | y* | $y^{*++}$ | $y^0$ | $y^{0++}$ | # |
|---|---|---|---|---|---|---|---|---|---|---|---|---|---|---|
| 1 | 115.0502 | 58.0287 | 98.0237 | 49.5155 | | | N | | | | | | | 42 |
| 2 | ***202.0822*** | 101.5448 | 185.0557 | 93.0315 | 184.0717 | 92.5395 | S | 4354.3140 | 2177.6607 | 4337.2875 | 2169.1474 | 4336.3035 | 2168.6554 | 41 |
| 3 | ***315.1663*** | 158.0868 | 298.1397 | 149.5735 | 297.1557 | 149.0815 | I | 4267.2820 | 2134.1446 | 4250.2555 | 2125.6314 | 4249.2714 | 2125.1394 | 40 |
| 4 | ***416.2140*** | 208.6106 | 399.1874 | 200.0974 | 398.2034 | 199.6053 | T | 4154.1979 | 2077.6026 | 4137.1714 | 2069.0893 | 4136.1874 | 2068.5973 | 39 |
| 5 | 529.2980 | 265.1527 | 512.2715 | 256.6394 | 511.2875 | 256.1474 | L | 4053.1503 | 2027.0788 | 4036.1237 | 2018.5655 | 4035.1397 | 2018.0735 | 38 |
| 6 | ***630.3457*** | 315.6765 | 613.3192 | 307.1632 | 612.3352 | 306.6712 | T | 3940.0662 | 1970.5367 | 3923.0397 | 1962.0235 | 3922.0556 | 1961.5315 | 37 |
| 7 | ***744.3886*** | 372.6980 | 727.3621 | 364.1847 | 726.3781 | 363.6927 | N | 3839.0185 | 1920.0129 | 3821.9920 | 1911.4996 | 3821.0080 | 1911.0076 | 36 |
| 8 | ***857.4727*** | 429.2400 | 840.4462 | 420.7267 | 839.4621 | 420.2347 | L | 3724.9756 | 1862.9914 | 3707.9490 | 1854.4782 | 3706.9650 | 1853.9862 | 35 |
| 9 | ***958.5204*** | 479.7638 | 941.4938 | 471.2506 | 940.5098 | 470.7585 | T | 3611.8915 | 1806.4494 | 3594.8650 | 1797.9361 | 3593.8810 | 1797.4441 | 34 |
| 10 | 1055.5732 | 528.2902 | 1038.5466 | 519.7769 | 1037.5626 | 519.2849 | P | 3510.8439 | 1755.9256 | 3493.8173 | 1747.4123 | 3492.8333 | 1746.9203 | 33 |
| 11 | 1112.5946 | 556.8009 | 1095.5681 | 548.2877 | 1094.5840 | 547.7957 | G | 3413.7911 | 1707.3992 | 3396.7645 | 1698.8859 | 3395.7805 | 1698.3939 | 32 |
| 12 | 1213.6423 | 607.3248 | 1196.6157 | 598.8115 | 1195.6317 | 598.3195 | T | 3356.7696 | 1678.8885 | 3339.7431 | 1670.3752 | 3338.7591 | 1669.8832 | 31 |
| 13 | 1342.6849 | 671.8461 | 1325.6583 | 663.3328 | 1324.6743 | 662.8408 | E | 3255.7219 | 1628.3646 | 3238.6954 | 1619.8513 | 3237.7114 | 1619.3593 | 30 |
| 14 | 1505.7482 | 753.3777 | 1488.7217 | 744.8645 | 1487.7377 | 744.3725 | Y | 3126.6794 | 1563.8433 | 3109.6528 | 1555.3300 | 3108.6688 | 1554.8380 | 29 |
| 15 | 1604.8166 | 802.9120 | 1587.7901 | 794.3987 | 1586.8061 | 793.9067 | V | 2963.6160 | 1482.3117 | 2946.5895 | 1473.7984 | 2945.6055 | 1473.3064 | 28 |
| 16 | 1703.8850 | 852.4462 | 1686.8585 | 843.9329 | 1685.8745 | 843.4409 | V | 2864.5476 | 1432.7774 | 2847.5211 | 1424.2642 | 2846.5370 | 1423.7722 | 27 |
| 17 | 1790.9171 | 895.9622 | 1773.8905 | 887.4489 | 1772.9065 | 886.9569 | S | 2765.4792 | 1383.2432 | 2748.4526 | 1374.7300 | 2747.4686 | 1374.2380 | 26 |
| 18 | 1904.0011 | 952.5042 | 1886.9746 | 943.9909 | 1885.9906 | 943.4989 | I | 2678.4472 | 1339.7272 | 2661.4206 | 1331.2139 | 2660.4366 | 1330.7219 | 25 |

| | | | | | | | | | | | | | | |
|---|---|---|---|---|---|---|---|---|---|---|---|---|---|---|
| 19 | 2003.0696 | 1002.0384 | 1986.0430 | 993.5251 | 1985.0590 | 993.0331 | V | 2565.3631 | 1283.1852 | 2548.3366 | 1274.6719 | 2547.3525 | 1274.1799 | 24 |
| 20 | 2074.1067 | 1037.5570 | 2057.0801 | 1029.0437 | 2056.0961 | 1028.5517 | A | 2466.2947 | 1233.6510 | 2449.2681 | 1225.1377 | 2448.2841 | 1224.6457 | 23 |
| 21 | 2187.1907 | 1094.0990 | 2170.1642 | 1085.5857 | 2169.1802 | 1085.0937 | L | 2395.2576 | 1198.1324 | 2378.2310 | 1189.6192 | 2377.2470 | 1189.1271 | 22 |
| 22 | 2301.2337 | 1151.1205 | 2284.2071 | 1142.6072 | 2283.2231 | 1142.1152 | N | 2282.1735 | 1141.5904 | 2265.1470 | 1133.0771 | 2264.1629 | 1132.5851 | 21 |
| 23 | 2358.2551 | 1179.6312 | 2341.2286 | 1171.1179 | 2340.2446 | 1170.6259 | G | ***2168.1306*** | 1084.5689 | 2151.1040 | 1076.0557 | 2150.1200 | 1075.5636 | 20 |
| 24 | 2514.3562 | 1257.6818 | 2497.3297 | 1249.1685 | 2496.3457 | 1248.6765 | R | 2111.1091 | 1056.0582 | 2094.0826 | 1047.5449 | 2093.0986 | 1047.0529 | 19 |
| 25 | 2643.3988 | 1322.2030 | 2626.3723 | 1313.6898 | 2625.3883 | 1313.1978 | E | 1955.0080 | 978.0076 | 1937.9815 | 969.4944 | 1936.9974 | 969.0024 | 18 |
| 26 | 2772.4414 | 1386.7243 | 2755.4149 | 1378.2111 | 2754.4309 | 1377.7191 | E | 1825.9654 | 913.4863 | 1808.9389 | 904.9731 | 1807.9549 | 904.4811 | 17 |
| 27 | 2859.4734 | 1430.2404 | 2842.4469 | 1421.7271 | 2841.4629 | 1421.2351 | S | 1696.9228 | 848.9651 | 1679.8963 | 840.4518 | 1678.9123 | 839.9598 | 16 |
| 28 | 2956.5262 | 1478.7667 | 2939.4997 | 1470.2535 | 2938.5156 | 1469.7615 | P | 1609.8908 | 805.4490 | 1592.8642 | 796.9358 | 1591.8802 | 796.4438 | 15 |
| 29 | 3069.6103 | 1535.3088 | 3052.5837 | 1526.7955 | 3051.5997 | 1526.3035 | L | 1512.8380 | 756.9227 | 1495.8115 | 748.4094 | 1494.8275 | 747.9174 | 14 |
| 30 | 3182.6943 | 1591.8508 | 3165.6678 | 1583.3375 | 3164.6838 | 1582.8455 | L | 1399.7540 | 700.3806 | 1382.7274 | 691.8673 | 1381.7434 | 691.3753 | 13 |
| 31 | 3295.7784 | 1648.3928 | 3278.7519 | 1639.8796 | 3277.7678 | 1639.3876 | I | ***1286.6699*** | 643.8386 | 1269.6434 | 635.3253 | 1268.6593 | 634.8333 | 12 |
| 32 | 3352.7999 | 1676.9036 | 3335.7733 | 1668.3903 | 3334.7893 | 1667.8983 | G | ***1173.5858*** | 587.2966 | 1156.5593 | 578.7833 | 1155.5753 | 578.2913 | 11 |
| 33 | 3480.8584 | 1740.9329 | 3463.8319 | 1732.4196 | 3462.8479 | 1731.9276 | Q | ***1116.5644*** | 558.7858 | 1099.5378 | 550.2726 | 1098.5538 | 549.7805 | 10 |
| 34 | 3608.9170 | 1804.9621 | 3591.8905 | 1796.4489 | 3590.9065 | 1795.9569 | Q | ***988.5058*** | 494.7565 | 971.4793 | 486.2433 | 970.4952 | 485.7513 | 9 |
| 35 | 3695.9490 | 1848.4782 | 3678.9225 | 1839.9649 | 3677.9385 | 1839.4729 | S | ***860.4472*** | 430.7272 | 843.4207 | 422.2140 | 842.4367 | 421.7220 | 8 |
| 36 | 3796.9967 | 1899.0020 | 3779.9702 | 1890.4887 | 3778.9862 | 1889.9967 | T | ***773.4152*** | 387.2112 | 756.3886 | 378.6980 | 755.4046 | 378.2060 | 7 |
| 37 | 3896.0651 | 1948.5362 | 3879.0386 | 1940.0229 | 3878.0546 | 1939.5309 | V | ***672.3675*** | 336.6874 | 655.3410 | 328.1741 | 654.3570 | 327.6821 | 6 |
| 38 | 3983.0972 | 1992.0522 | 3966.0706 | 1983.5389 | 3965.0866 | 1983.0469 | S | ***573.2991*** | 287.1532 | 556.2726 | 278.6399 | 555.2885 | 278.1479 | 5 |
| 39 | 4098.1241 | 2049.5657 | 4081.0976 | 2041.0524 | 4080.1135 | 2040.5604 | D | ***486.2671*** | 243.6372 | 469.2405 | 235.1239 | 468.2565 | 234.6319 | 4 |
| 40 | 4197.1925 | 2099.0999 | 4180.1660 | 2090.5866 | 4179.1820 | 2090.0946 | V | ***371.2401*** | 186.1237 | 354.2136 | 177.6104 | | | 3 |
| 41 | 4294.2453 | 2147.6263 | 4277.2187 | 2139.1130 | 4276.2347 | 2138.6210 | P | ***272.1717*** | 136.5895 | 255.1452 | 128.0762 | | | 2 |
| 42 | | | | | | | R | ***175.1190*** | 88.0631 | 158.0924 | 79.5498 | | | 1 |

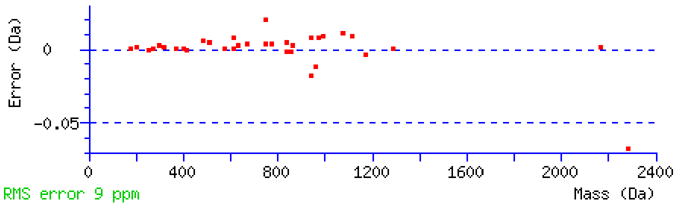


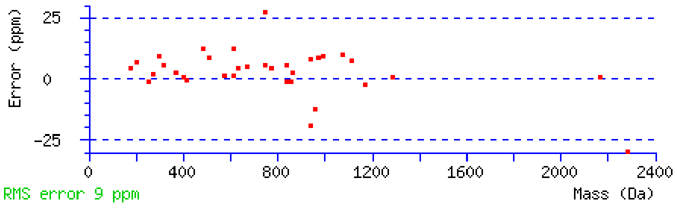


NCBI **BLAST** search of NSITLTNLTPGTEYVVSIVALNGREESPLLIGQQSTVSDVPR
(Parameters: blastp, nr protein database, expect=20000, no filter, PAM30)
Other BLAST web gateways

**All matches to this query**

| Score | Mr(calc) | Delta | Sequence |
|---|---|---|---|
| 68.1 | 4467.3497 | 1.0079 | NSITLTNLTPGTEYVVSIVALNGREESPLLIGQQSTVSDVPR |

**Mascot:** http://www.matrixscience.com/

### MATRIX SCIENCE Mascot Search Results

#### Peptide View

MS/MS Fragmentation of **NSITLTNLTPGTEYVVSIVALNGREESPLLIGQQSTVSDVPR**
Found in **sp|P02751|FINC_HUMAN** in **uni_human_i**, sp|P02751|FINC_HUMAN Fibronectin OS=Homo sapiens GN=FN1 PE=1 SV=4

Match to Query 39070: 4470.359096 from(1118.597050,4+) intensity(1009260.1250) rtinseconds(1873) scans(13033) index(20066)
Title: Fibronectin_ERLIC_MSMS_60min_Spectrum059198_scans__13033_RTINSECONDS=1873
Data file V:\\raw\\Cam\\Fibronetin_ERLIC\\mgf\\T\\TFibronectin_ERLIC_MSMS_60min.mgf

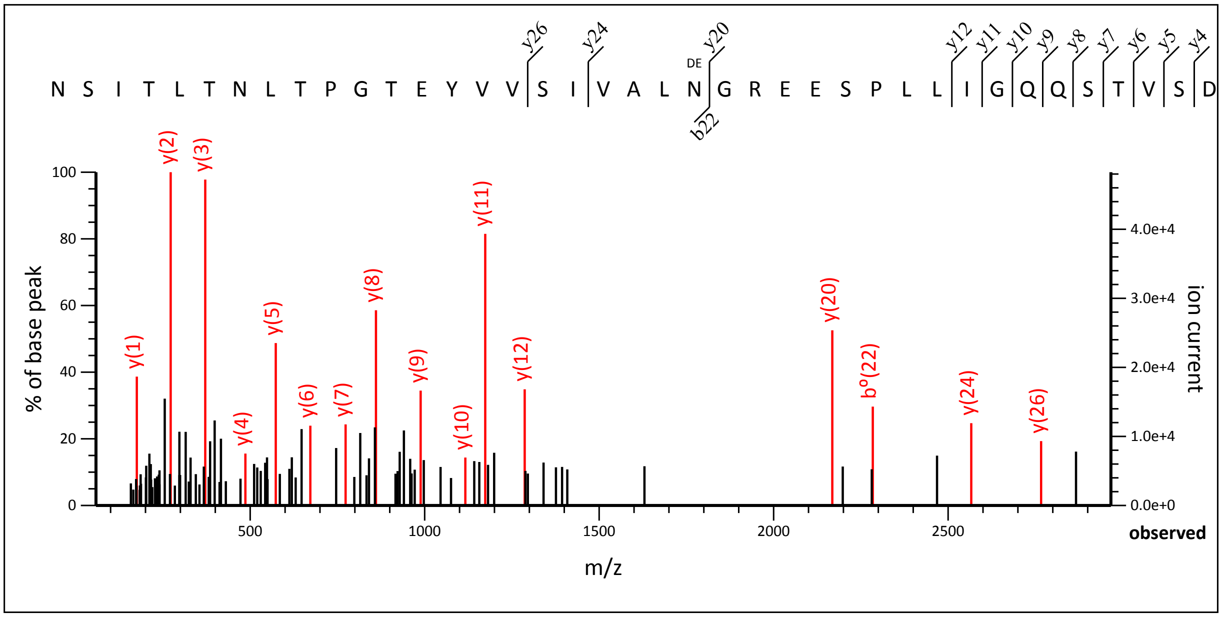


58.09 to 2966.54

Label all possible matches ○ Label matches used for scoring ◉

**Monoisotopic mass of neutral peptide Mr(calc):** 4468.3337
**Fixed modifications:** Carbamidomethyl (C) (apply to specified residues or termini only)
**Variable modifications:**
N22 : Deamidated (NQ)
**Ions Score:** 111 **Expect:** 4.4e-010
**Matches :** 16/484 fragment ions using 20 most intense peaks (help)

| # | b | b++ | b* | b*++ | b0 | b0++ | Seq. | y | y++ | y* | y*++ | y0 | y0++ | # |
|---|---|---|---|---|---|---|---|---|---|---|---|---|---|---|
| 1 | 115.0502 | 58.0287 | 98.0237 | 49.5155 | | | N | | | | | | | 42 |
| 2 | 202.0822 | 101.5448 | 185.0557 | 93.0315 | 184.0717 | 92.5395 | S | 4355.2981 | 2178.1527 | 4338.2715 | 2169.6394 | 4337.2875 | 2169.1474 | 41 |
| 3 | 315.1663 | 158.0868 | 298.1397 | 149.5735 | 297.1557 | 149.0815 | I | 4268.2660 | 2134.6367 | 4251.2395 | 2126.1234 | 4250.2555 | 2125.6314 | 40 |
| 4 | 416.2140 | 208.6106 | 399.1874 | 200.0974 | 398.2034 | 199.6053 | T | 4155.1820 | 2078.0946 | 4138.1554 | 2069.5813 | 4137.1714 | 2069.0893 | 39 |
| 5 | 529.2980 | 265.1527 | 512.2715 | 256.6394 | 511.2875 | 256.1474 | L | 4054.1343 | 2027.5708 | 4037.1077 | 2019.0575 | 4036.1237 | 2018.5655 | 38 |
| 6 | 630.3457 | 315.6765 | 613.3192 | 307.1632 | 612.3352 | 306.6712 | T | 3941.0502 | 1971.0287 | 3924.0237 | 1962.5155 | 3923.0397 | 1962.0235 | 37 |
| 7 | 744.3886 | 372.6980 | 727.3621 | 364.1847 | 726.3781 | 363.6927 | N | 3840.0025 | 1920.5049 | 3822.9760 | 1911.9916 | 3821.9920 | 1911.4996 | 36 |
| 8 | 857.4727 | 429.2400 | 840.4462 | 420.7267 | 839.4621 | 420.2347 | L | 3725.9596 | 1863.4834 | 3708.9331 | 1854.9702 | 3707.9490 | 1854.4782 | 35 |
| 9 | 958.5204 | 479.7638 | 941.4938 | 471.2506 | 940.5098 | 470.7585 | T | 3612.8756 | 1806.9414 | 3595.8490 | 1798.4281 | 3594.8650 | 1797.9361 | 34 |
| 10 | 1055.5732 | 528.2902 | 1038.5466 | 519.7769 | 1037.5626 | 519.2849 | P | 3511.8279 | 1756.4176 | 3494.8013 | 1747.9043 | 3493.8173 | 1747.4123 | 33 |
| 11 | 1112.5946 | 556.8009 | 1095.5681 | 548.2877 | 1094.5840 | 547.7957 | G | 3414.7751 | 1707.8912 | 3397.7486 | 1699.3779 | 3396.7645 | 1698.8859 | 32 |
| 12 | 1213.6423 | 607.3248 | 1196.6157 | 598.8115 | 1195.6317 | 598.3195 | T | 3357.7536 | 1679.3805 | 3340.7271 | 1670.8672 | 3339.7431 | 1670.3752 | 31 |
| 13 | 1342.6849 | 671.8461 | 1325.6583 | 663.3328 | 1324.6743 | 662.8408 | E | 3256.7060 | 1628.8566 | 3239.6794 | 1620.3433 | 3238.6954 | 1619.8513 | 30 |
| 14 | 1505.7482 | 753.3777 | 1488.7217 | 744.8645 | 1487.7377 | 744.3725 | Y | 3127.6634 | 1564.3353 | 3110.6368 | 1555.8220 | 3109.6528 | 1555.3300 | 29 |
| 15 | 1604.8166 | 802.9120 | 1587.7901 | 794.3987 | 1586.8061 | 793.9067 | V | 2964.6000 | 1482.8037 | 2947.5735 | 1474.2904 | 2946.5895 | 1473.7984 | 28 |
| 16 | 1703.8850 | 852.4462 | 1686.8585 | 843.9329 | 1685.8745 | 843.4409 | V | 2865.5316 | 1433.2695 | 2848.5051 | 1424.7562 | 2847.5211 | 1424.2642 | 27 |
| 17 | 1790.9171 | 895.9622 | 1773.8905 | 887.4489 | 1772.9065 | 886.9569 | S | *2766.4632* | 1383.7352 | 2749.4367 | 1375.2220 | 2748.4526 | 1374.7300 | 26 |

| | | | | | | | | | | | | | | |
|---|---|---|---|---|---|---|---|---|---|---|---|---|---|---|
| **18** | 1904.0011 | 952.5042 | 1886.9746 | 943.9909 | 1885.9906 | 943.4989 | **I** | 2679.4312 | 1340.2192 | 2662.4046 | 1331.7060 | 2661.4206 | 1331.2139 | **25** |
| **19** | 2003.0696 | 1002.0384 | 1986.0430 | 993.5251 | 1985.0590 | 993.0331 | **V** | ***2566.3471*** | 1283.6772 | 2549.3206 | 1275.1639 | 2548.3366 | 1274.6719 | **24** |
| **20** | 2074.1067 | 1037.5570 | 2057.0801 | 1029.0437 | 2056.0961 | 1028.5517 | **A** | 2467.2787 | 1234.1430 | 2450.2522 | 1225.6297 | 2449.2681 | 1225.1377 | **23** |
| **21** | 2187.1907 | 1094.0990 | 2170.1642 | 1085.5857 | 2169.1802 | 1085.0937 | **L** | 2396.2416 | 1198.6244 | 2379.2150 | 1190.1112 | 2378.2310 | 1189.6192 | **22** |
| **22** | 2302.2177 | 1151.6125 | 2285.1911 | 1143.0992 | 2284.2071 | 1142.6072 | **N** | 2283.1575 | 1142.0824 | 2266.1310 | 1133.5691 | 2265.1470 | 1133.0771 | **21** |
| **23** | 2359.2391 | 1180.1232 | 2342.2126 | 1171.6099 | 2341.2286 | 1171.1179 | **G** | ***2168.1306*** | 1084.5689 | 2151.1040 | 1076.0557 | 2150.1200 | 1075.5636 | **20** |
| **24** | 2515.3402 | 1258.1738 | 2498.3137 | 1249.6605 | 2497.3297 | 1249.1685 | **R** | 2111.1091 | 1056.0582 | 2094.0826 | 1047.5449 | 2093.0986 | 1047.0529 | **19** |
| **25** | 2644.3828 | 1322.6951 | 2627.3563 | 1314.1818 | 2626.3723 | 1313.6898 | **E** | 1955.0080 | 978.0076 | 1937.9815 | 969.4944 | 1936.9974 | 969.0024 | **18** |
| **26** | 2773.4254 | 1387.2164 | 2756.3989 | 1378.7031 | 2755.4149 | 1378.2111 | **E** | 1825.9654 | 913.4863 | 1808.9389 | 904.9731 | 1807.9549 | 904.4811 | **17** |
| **27** | 2860.4575 | 1430.7324 | 2843.4309 | 1422.2191 | 2842.4469 | 1421.7271 | **S** | 1696.9228 | 848.9651 | 1679.8963 | 840.4518 | 1678.9123 | 839.9598 | **16** |
| **28** | 2957.5102 | 1479.2588 | 2940.4837 | 1470.7455 | 2939.4997 | 1470.2535 | **P** | 1609.8908 | 805.4490 | 1592.8642 | 796.9358 | 1591.8802 | 796.4438 | **15** |
| **29** | 3070.5943 | 1535.8008 | 3053.5677 | 1527.2875 | 3052.5837 | 1526.7955 | **L** | 1512.8380 | 756.9227 | 1495.8115 | 748.4094 | 1494.8275 | 747.9174 | **14** |
| **30** | 3183.6784 | 1592.3428 | 3166.6518 | 1583.8295 | 3165.6678 | 1583.3375 | **L** | 1399.7540 | 700.3806 | 1382.7274 | 691.8673 | 1381.7434 | 691.3753 | **13** |
| **31** | 3296.7624 | 1648.8848 | 3279.7359 | 1640.3716 | 3278.7519 | 1639.8796 | **I** | ***1286.6699*** | 643.8386 | 1269.6434 | 635.3253 | 1268.6593 | 634.8333 | **12** |
| **32** | 3353.7839 | 1677.3956 | 3336.7573 | 1668.8823 | 3335.7733 | 1668.3903 | **G** | ***1173.5858*** | 587.2966 | 1156.5593 | 578.7833 | 1155.5753 | 578.2913 | **11** |
| **33** | 3481.8425 | 1741.4249 | 3464.8159 | 1732.9116 | 3463.8319 | 1732.4196 | **Q** | ***1116.5644*** | 558.7858 | 1099.5378 | 550.2726 | 1098.5538 | 549.7805 | **10** |
| **34** | 3609.9010 | 1805.4542 | 3592.8745 | 1796.9409 | 3591.8905 | 1796.4489 | **Q** | ***988.5058*** | 494.7565 | 971.4793 | 486.2433 | 970.4952 | 485.7513 | **9** |
| **35** | 3696.9331 | 1848.9702 | 3679.9065 | 1840.4569 | 3678.9225 | 1839.9649 | **S** | ***860.4472*** | 430.7272 | 843.4207 | 422.2140 | 842.4367 | 421.7220 | **8** |
| **36** | 3797.9807 | 1899.4940 | 3780.9542 | 1890.9807 | 3779.9702 | 1890.4887 | **T** | ***773.4152*** | 387.2112 | 756.3886 | 378.6980 | 755.4046 | 378.2060 | **7** |
| **37** | 3897.0492 | 1949.0282 | 3880.0226 | 1940.5149 | 3879.0386 | 1940.0229 | **V** | ***672.3675*** | 336.6874 | 655.3410 | 328.1741 | 654.3570 | 327.6821 | **6** |
| **38** | 3984.0812 | 1992.5442 | 3967.0546 | 1984.0310 | 3966.0706 | 1983.5389 | **S** | ***573.2991*** | 287.1532 | 556.2726 | 278.6399 | 555.2885 | 278.1479 | **5** |
| **39** | 4099.1081 | 2050.0577 | 4082.0816 | 2041.5444 | 4081.0976 | 2041.0524 | **D** | ***486.2671*** | 243.6372 | 469.2405 | 235.1239 | 468.2565 | 234.6319 | **4** |
| **40** | 4198.1765 | 2099.5919 | 4181.1500 | 2091.0786 | 4180.1660 | 2090.5866 | **V** | ***371.2401*** | 186.1237 | 354.2136 | 177.6104 | | | **3** |
| **41** | 4295.2293 | 2148.1183 | 4278.2028 | 2139.6050 | 4277.2187 | 2139.1130 | **P** | ***272.1717*** | 136.5895 | 255.1452 | 128.0762 | | | **2** |
| **42** | | | | | | | **R** | ***175.1190*** | 88.0631 | 158.0924 | 79.5498 | | | **1** |

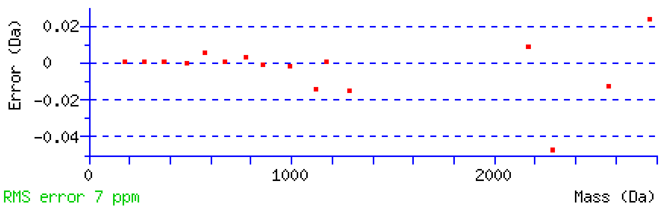


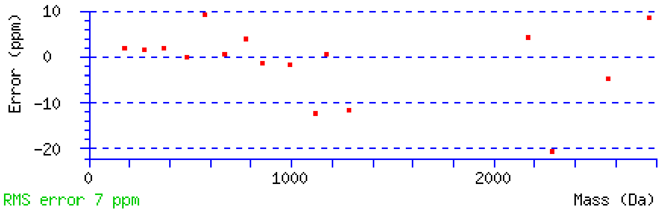


NCBI **BLAST** search of NSITLTNLTPGTEYVVSIVALNGREESPLLIGQQSTVSDVPR
(Parameters: blastp, nr protein database, expect=20000, no filter, PAM30)
Other BLAST web gateways

**All matches to this query**

| Score | Mr(calc) | Delta | Sequence | Site Analysis |
|---|---|---|---|---|
| 110.9 | 4468.3337 | 2.0254 | NSITLTNLTPGTEYVVSIVALNGREESPLLIGQQSTVSDVPR | Deamidated N22 99.72% |
| 82.5 | 4468.3337 | 2.0254 | NSITLTNLTPGTEYVVSIVALNGREESPLLIGQQSTVSDVPR | Deamidated N7 0.14% |
| 82.3 | 4468.3337 | 2.0254 | NSITLTNLTPGTEYVVSIVALNGREESPLLIGQQSTVSDVPR | Deamidated N1 0.14% |
| 57.1 | 4468.3337 | 2.0254 | NSITLTNLTPGTEYVVSIVALNGREESPLLIGQQSTVSDVPR | Deamidated Q33 0.00% |
| 46.1 | 4468.3337 | 2.0254 | NSITLTNLTPGTEYVVSIVALNGREESPLLIGQQSTVSDVPR | Deamidated Q34 0.00% |

**Mascot:** http://www.matrixscience.com/
