## Supplementary figures and images for "Aging-induced isoDGR-modified fibronectin activates monocytic and endothelial cells to promote atherosclerosis"

### Supplementary Data 5

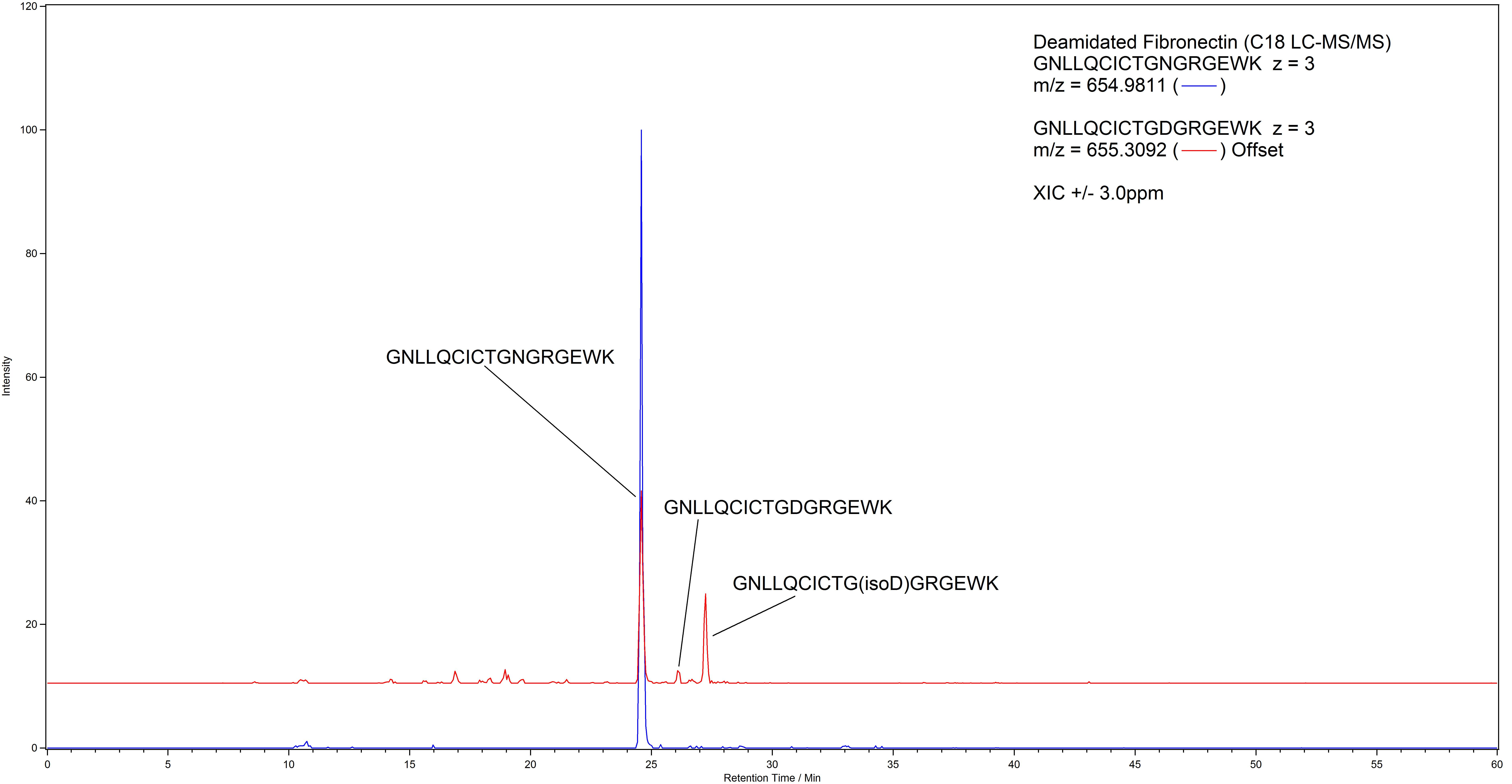

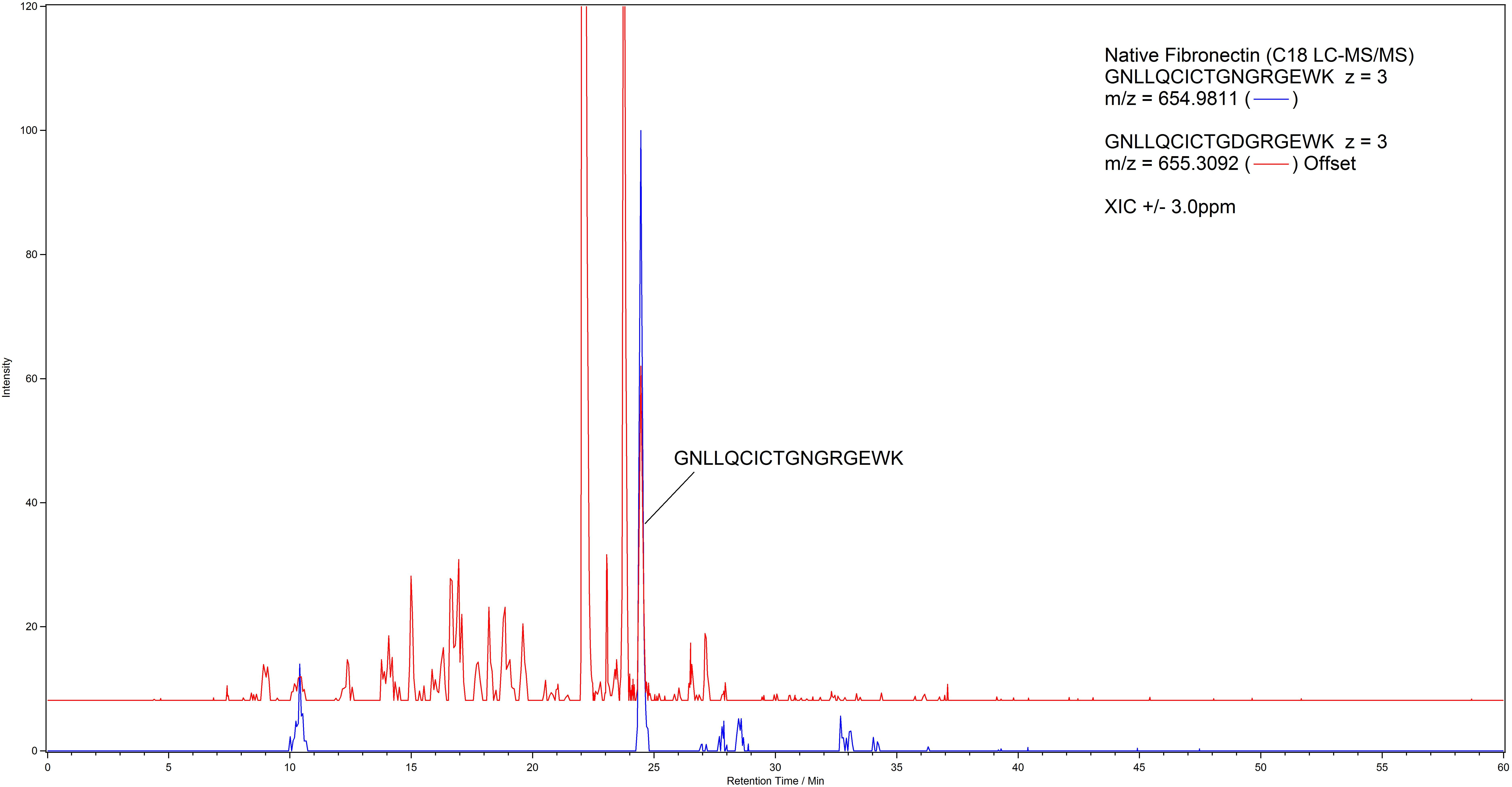

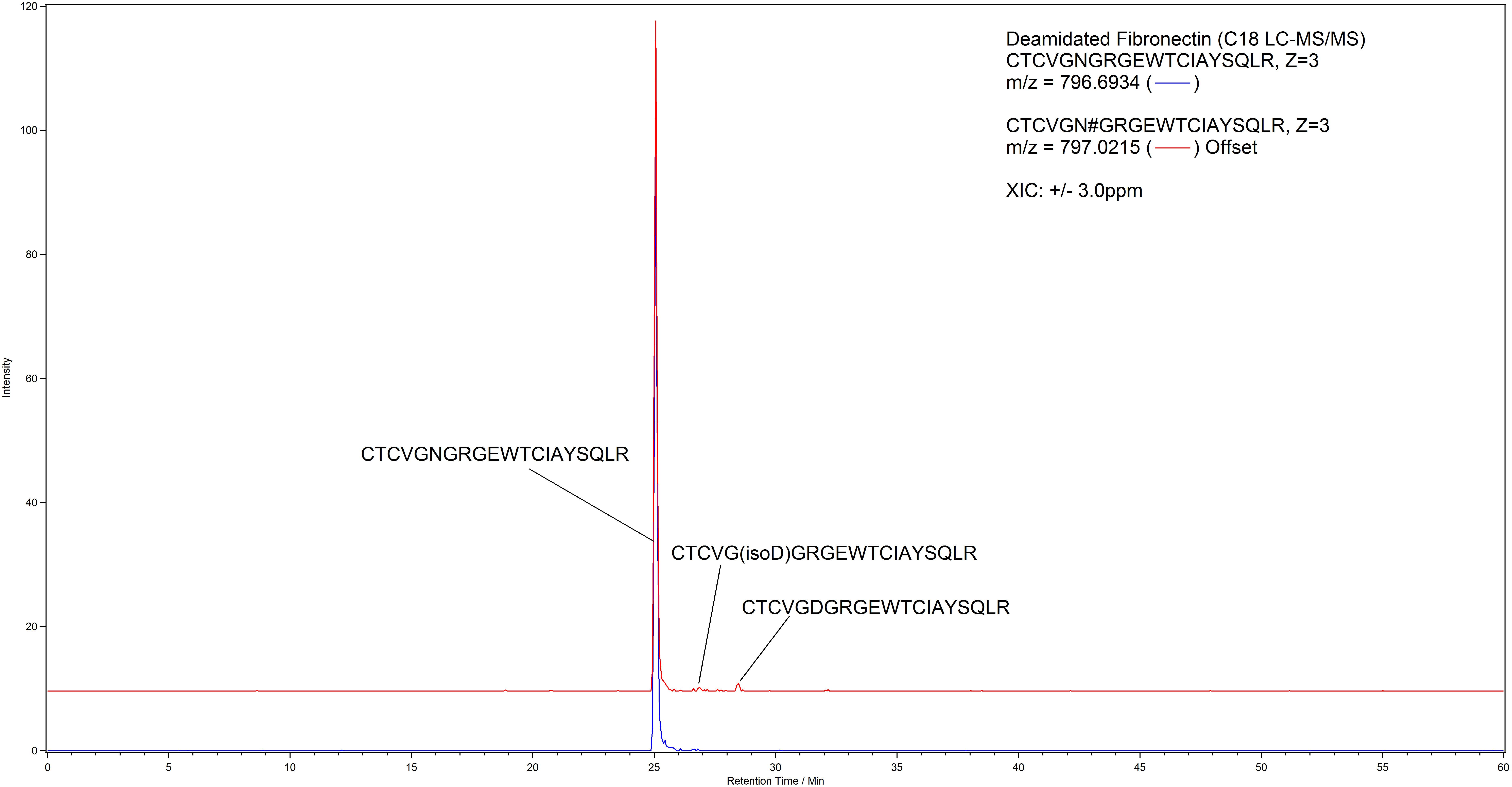

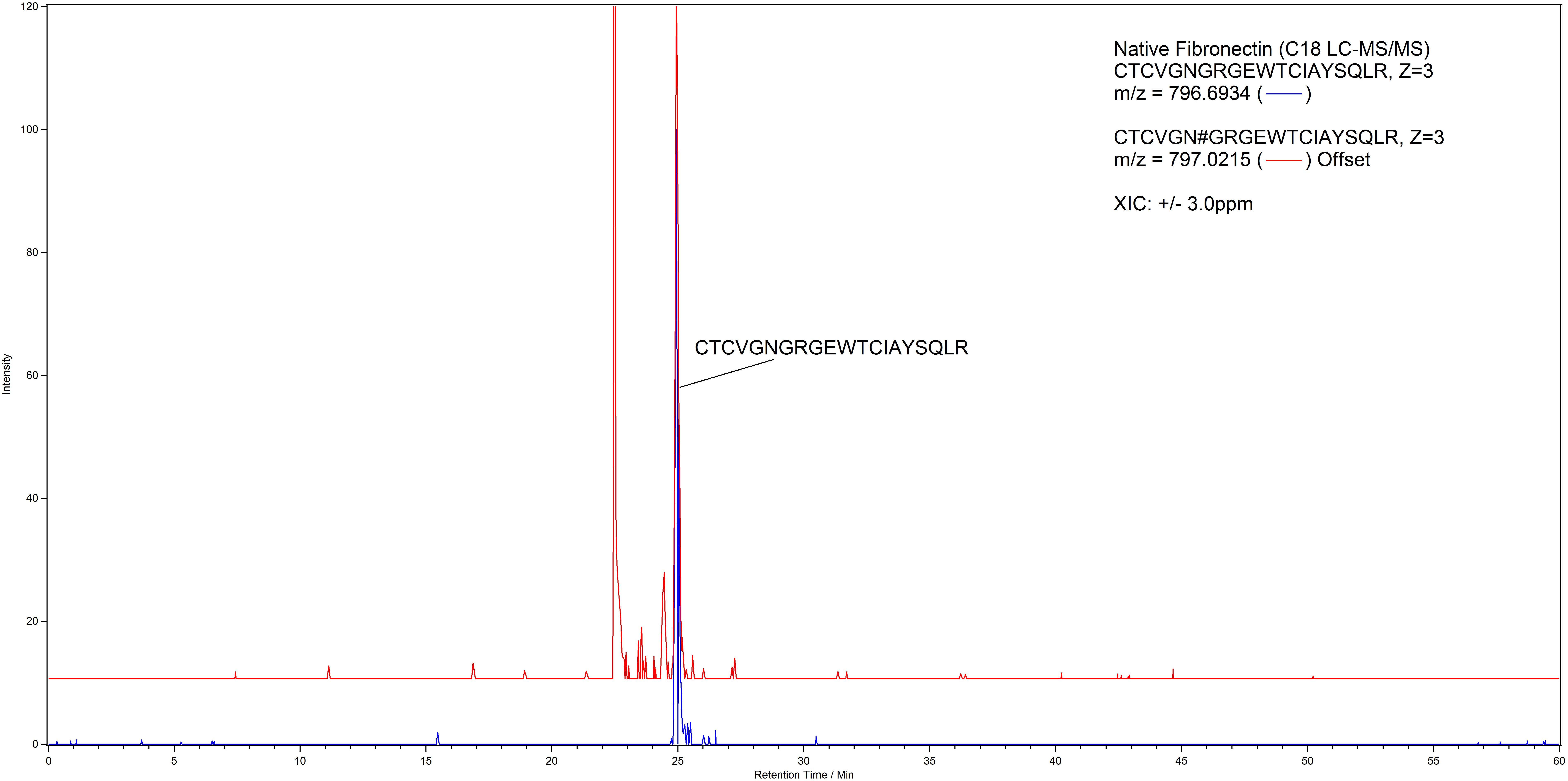

### Supplementary Data 6

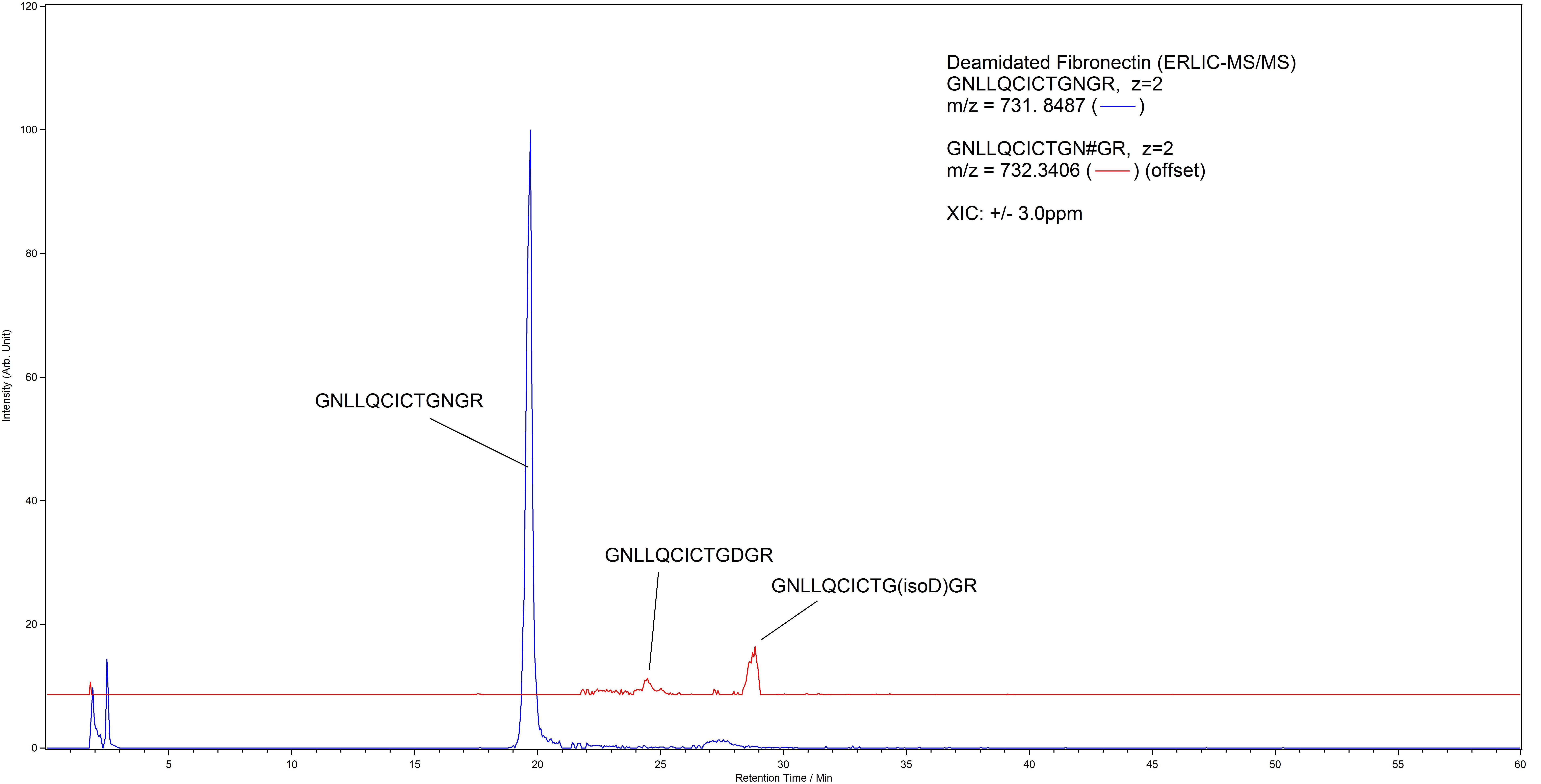

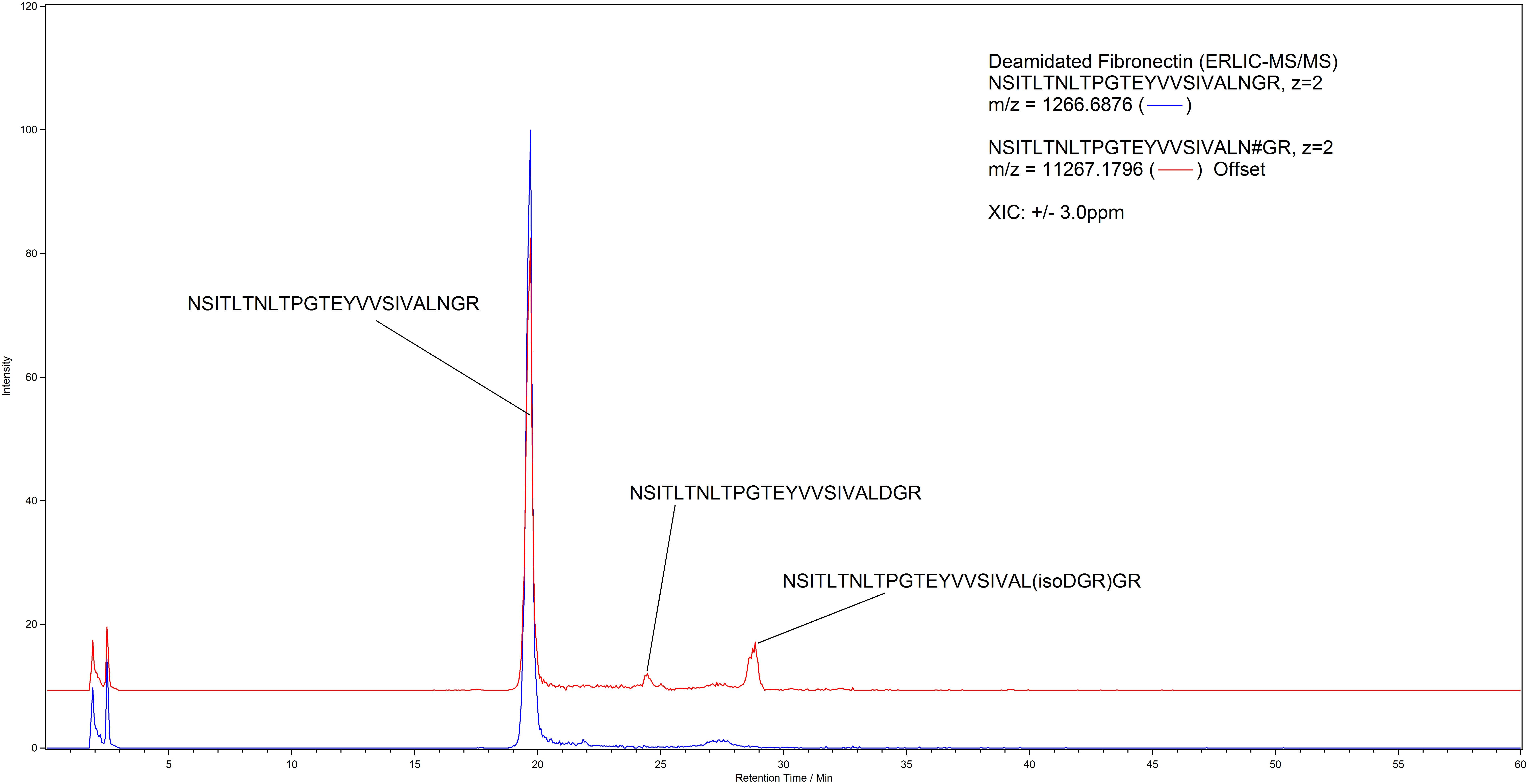

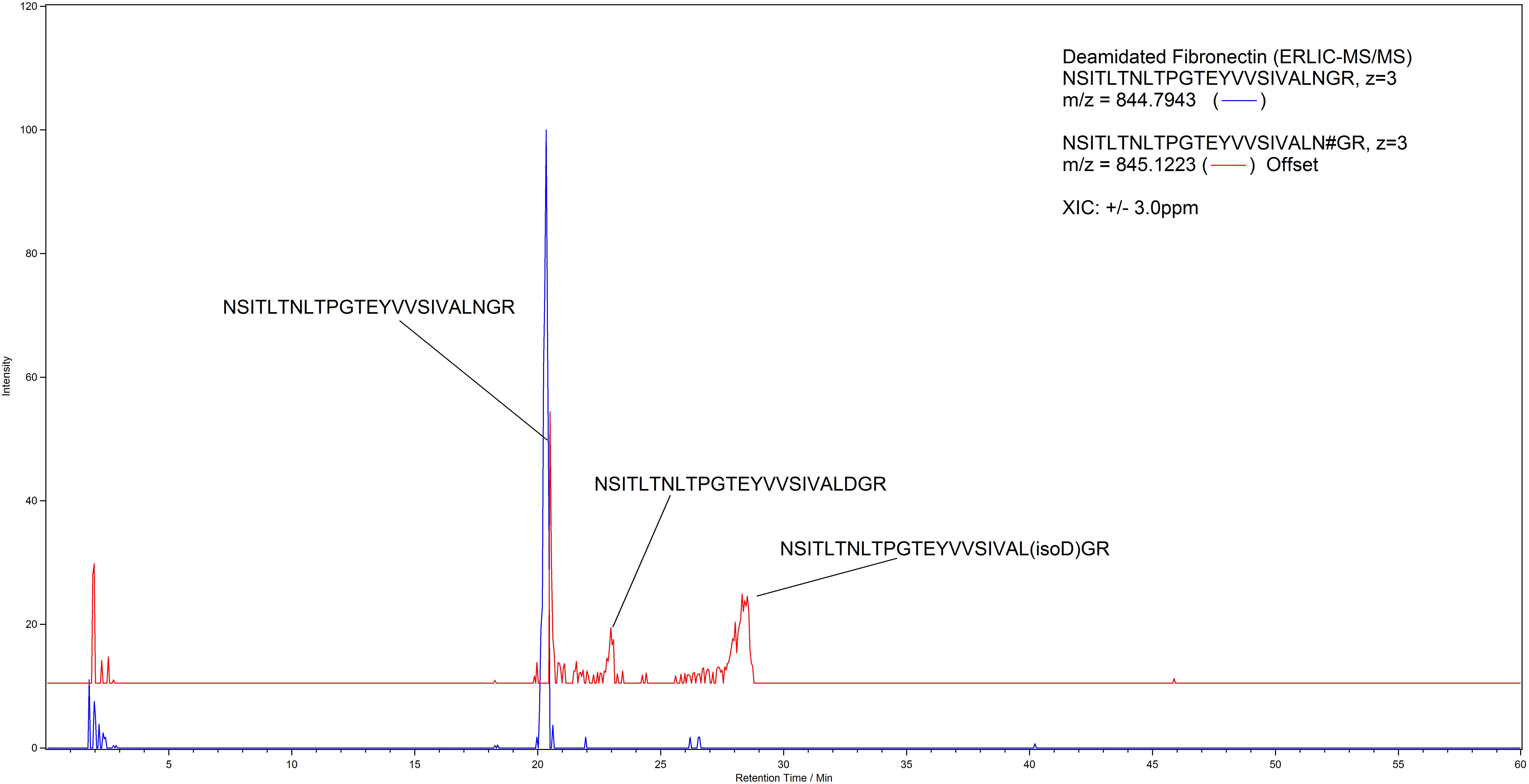

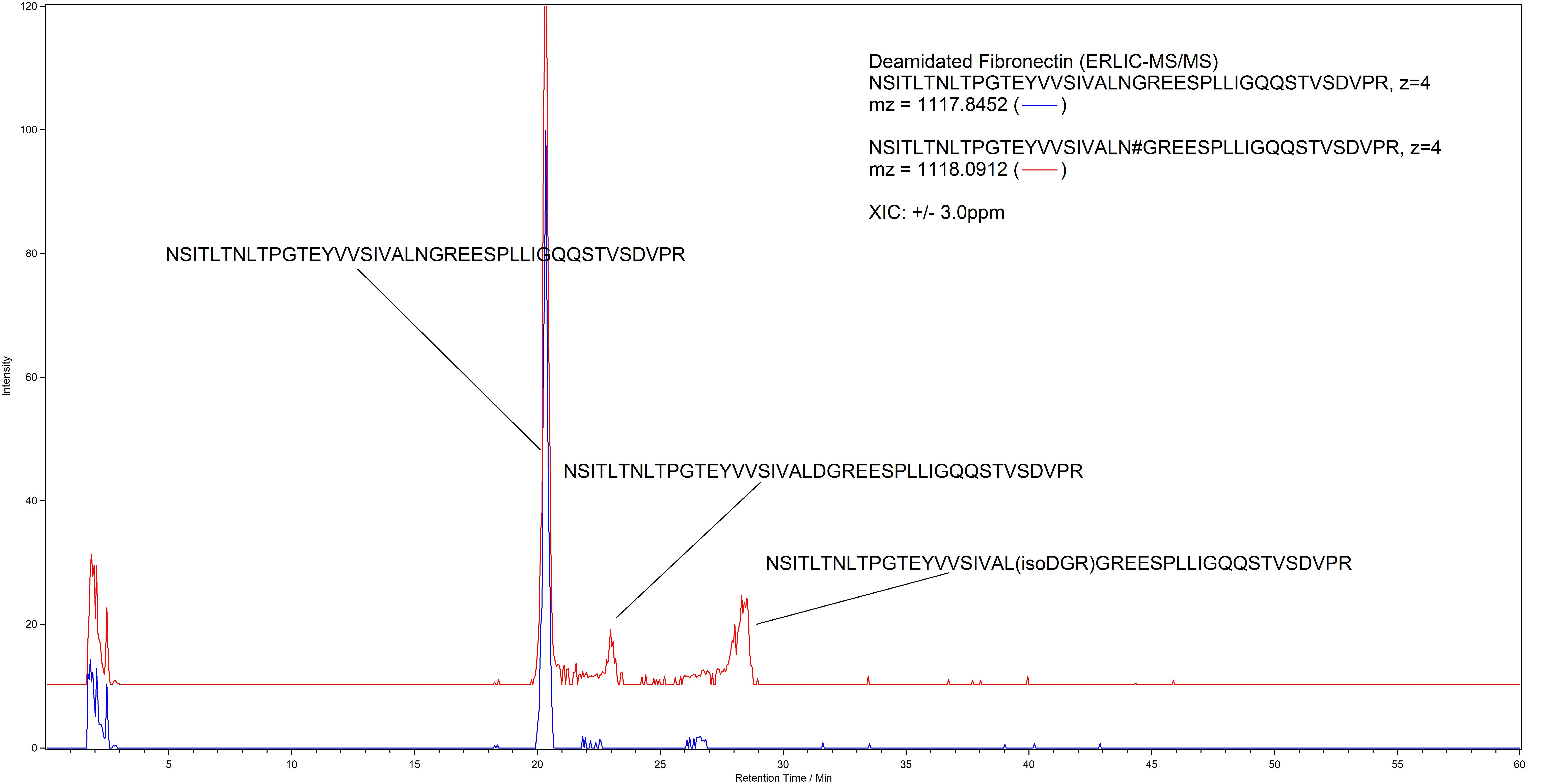
