## Supplementary Data 7 for "Aging-induced isoDGR-modified fibronectin activates monocytic and endothelial cells to promote atherosclerosis"

| S.No | Gene | Sequence 3’-5’ |
| --- | --- | --- |
| 1 | MCP-1 | FP- TGCTACTCATTCACCAGCAA  RP- GTCTGGACCCATTCCTTCTT |
| 2 | TNFα | FP- TCAGCCTCTTCTCATTCCTG  RP- TGGTGGTTTGTGAGTGTGAG |
| 3 | MMP9 | FP- CCATGTCACTTTCCCTTCAC  RP- CTCACTAGGGCAGAAACCAA |
| 4 | GM-CSF | FP- TTGAACATGACAGCCAGCTA  RP- TTTTGCATTCAAAGGGGATA |
| 5 | CCL4 | FP- CCCTCTCTCTCCTCTTGCTC  RP- TCTGTCTGCCTCTTTTGGTC |
| 6 | IFN-β | FP- GAACATTCGGAAATGTCAGG  RP- CTGCATCTTCTCCGTCATCT |
| 7 | β-actin | FP- GCGTGACATCAAAGAGAAGC  RP- CAGGCAGCTCATAGCTCTTC |
| 8 | GAPDH | FP- CTGGAGAAACCTGCCAAGTA  RP- AAGAGTGGGAGTTGCTGTTG |

qPCR Primers
