## Supplementary Data 1 for "Aging-induced isoDGR-modified fibronectin activates monocytic and endothelial cells to promote atherosclerosis"

| **Subject**  **No** | **Disease** | **Treatment** | **Gender** | **Age Range** | **EF** | **DM** | **ESRF** | **Smoking** | **Hypertension** | **Hyperlipidaemia** | **Lipid medication (statin therapy)** | | **PVD** |
| --- | --- | --- | --- | --- | --- | --- | --- | --- | --- | --- | --- | --- | --- |
| 1 | CAD | OPCAB | Male | 61-65 | >45% | Yes | No | Yes | Yes | Yes | | Yes | No |
| 2 | CAD | CABG | Male | 66-70 | >45% | Yes | No | No | Yes | Yes | | Yes | No |
| 3 | CAD | CABG | Male | 66-70 | >45% | Yes | No | Yes | Yes | Yes | | Yes | No |
| 4 | CAD | CABG | Male | 56-60 | 45-35% | Yes | No | No | Yes | Yes | | Yes | No |
| 5 | CAD | Redo CABG & Femoral Cannulation | Male | 61-65 | >45% | No | No |  | No | Yes | | Yes | No |
| 6 | CAD | CABG | Male | 51-55 | <35% | Yes | No | Yes | Yes | Yes | | Yes | No |
| 7 | CAD | CABG | Male | 56-60 | >45% | Yes | No | Yes | Yes | Yes | | Yes | No |
| 8 | CAD | CABG | Male | 71-75 | >45% | Yes | No | Yes | Yes | Yes | | Yes | No |
| 9 | CAD | CABG | Male | 76-80 | 45-35% | Yes | No | No | No | Yes | | Yes | No |
| 10 | CAD | CABG | Female | 66-70 |  | No | No | No | Yes | Yes | | Yes | No |
| 11 | CAD | CABG | Male | 41-45 | 45-35% | No | No | Yes | Yes | Yes | | Yes | No |
| 12 | CAD | CABG | Male | 51-55 | <35% | Yes | No | No | Yes | Yes | | Yes | No |
| 13 | CAD | CABG | Female | 61-65 | >45% | Yes | No | No | Yes | Yes | | Yes | No |
| 14 | CAD | CABG | Male | 61-65 | >45% | No | No | No | Yes | Yes | | Yes | No |
| 15 | CAD | CABG | Female | 66-70 | >45% | Yes | No | No | Yes | Yes | | Yes | No |
| 16 | CAD | CABG | Male | 61-65 | >45% | No | No | Yes | Yes | Yes | | Yes | No |
| 17 | CAD | CABG | Male | 66-70 | >45% | Yes | No | Yes | Yes | Yes | | Yes | No |
| 18 | CAD | CABG | Male | 71-75 |  | Yes | No |  | Yes | Yes | | Yes | No |
| 19 | CAD | CABG | Female | 61-65 | >45% | No | No | No | Yes | Yes | | Yes | No |
| 20 | CAD | Redo-sternotomy CABG | Female | 66-70 | >45% | Yes | No | No | No | Yes | | Yes | No |
| 21 | CAD | CABG | Male | 61-65 | >45% | Yes | No | No | Yes | Yes | | Yes | No |
| 22 | CAD | Minimally Invasive CABG | Male | 51-55 | >45% | No | No | No | No | Yes | | Yes | No |
| 23 | CAD | CABG | Female | 66-70 | >45% | No | No | No | Yes | Yes | | Yes | No |
| 24 | CAD | CABG | Male | 51-55 | <35% | No | No | Yes | No | Yes | | Yes | No |
| 25 | CAD | CABG | Female | 56-60 | >45% | No | No | No | No | Yes | | Yes | No |

**Supplementary Table 1. Clinical characteristics of CAD patients**

**Supplementary Table 2. Clinical characteristics of control subjects**

| **Subject no** | **Group** | **Treatment** | **Gender** | **Age**  **Range** | **EF** | **DM** | **ESRF** | **Smoking** | **Hypertension** | **Hyperlipidaemia** | **Lipid medication (statin therapy)** | **PVD** |
| --- | --- | --- | --- | --- | --- | --- | --- | --- | --- | --- | --- | --- |
| 1 | Non-CAD | Conservative | Male | 31-35 | >45% | No | No | Yes | No | No | No | No |
| 2 | Non-CAD | Conservative | Male | 56-60 | >45% | No | No | No | Yes | No | No | No |
| 3 | Non-CAD | Conservative | Male | 41-45 | >45% | No | No | No | No | Yes | No | No |
| 4 | Non-CAD | Conservative | Female | 66-70 | >45% | No | No | No | No | Yes | Yes | No |
| 5 | Non-CAD | Conservative | Female | 51-55 | >45% | Yes | No | No | Yes | No | No | No |
| 6 | Non-CAD | Conservative | Female | 56-60 | >45% | No | No | No | No | Yes | Yes | No |
| 7 | Non-CAD | Conservative | Male | 46-50 | >45% | No | No | No | No | No | No | No |
| 8 | Non-CAD | Conservative | Male | 56-60 | >45% | No | No | No | No | No | No | No |
| 9 | Non-CAD | Conservative | Male | 46-50 | >45% | No | No | No | No | No | No | No |
| 10 | Non-CAD | Conservative | Male | 56-60 | >45% | No | No | No | No | Yes | Yes | No |
| 11 | Non-CAD | Conservative | Male | 46-50 | >45% | No | No | No | No | No | No | No |
| 12 | Non-CAD | Conservative | Female | 46-50 | >45% | Yes | No | No | No | No | Yes | No |
| 13 | Non-CAD | Conservative | Female | 56-60 | >45% | No | No | No | No | No | No | No |
| 14 | Non-CAD | Conservative | Female | 66-70 | >45% | No | No | No | No | Yes | Yes | No |
| 15 | Non-CAD | Conservative | Male | 46-50 | >45% | No | No | No | No | No | No | No |
| 16 | Non-CAD | Conservative | Male | 51-55 | >45% | No | No | No | Yes | No | No | No |
| 17 | Non-CAD | Conservative | Male | 61-65 | >45% | No | No | No | No | No | Yes | No |
| 18 | Non-CAD | Conservative | Female | 51-55 | >45% | No | No | No | No | Yes | Yes | No |
| 19 | Non-CAD | Conservative | Male | 61-65 | >45% | No | No | No | Yes | No | No | No |
| 20 | Non-CAD | Conservative | Male | 51-55 | >45% | No | No | No | No | No | Yes | No |
| 21 | Non-CAD | Conservative | Female | 56-60 | >45% | No | No | No | No | No | No | No |
| 22 | Non-CAD | Conservative | Female | 61-65 | >45% | No | No | No | Yes | No | Yes | No |
| 23 | Non-CAD | Conservative | Male | 46-50 | >45% | No | No | No | No | Yes | No | No |
| 24 | Non-CAD | Conservative | Female | 61-65 | >45% | No | No | No | No | Yes | Yes | No |
| 25 | Non-CAD | Conservative | Male | 61-65 | >45% | No | No | No | No | No | Yes | No |
